# An atlas of trait associations with resting-state and task-evoked human brain functional architectures in the UK Biobank

**DOI:** 10.1101/2022.02.22.22271371

**Authors:** Bingxin Zhao, Tengfei Li, Yujue Li, Zirui Fan, Di Xiong, Xifeng Wang, Mufeng Gao, Stephen M. Smith, Hongtu Zhu

## Abstract

Functional magnetic resonance imaging (fMRI) has been widely used to identify brain regions linked to critical functions, such as language and vision, and to detect tumors, strokes, brain injuries, and diseases. It is now known that large sample sizes are necessary for fMRI studies to detect small effect sizes and produce reproducible results. Here we report a systematic association analysis of 647 traits with imaging features extracted from resting-state and task-evoked fMRI data of more than 40,000 UK Biobank participants. We used a parcellation-based approach to generate 64,620 functional connectivity measures to reveal fine-grained details about cerebral cortex functional architectures. The difference between functional organizations at rest and during task has been quantified, and we have prioritized important brain regions and networks associated with a variety of human traits and clinical outcomes. For example, depression was most strongly associated with decreased connectivity in the somatomotor network. We have made our results publicly available and developed a browser framework to facilitate exploration of brain function-trait association results (http://165.227.92.206/).

---

Functional magnetic resonance imaging (fMRI) is a noninvasive and comprehensive method of assessing functional architectures of the human brain. By measuring blood oxygen level dependent (BOLD) signal changes, fMRI can map complex brain functions and estimate neural correlations between different brain regions^1^. When the subject is performing a specific task, fMRI can detect brain signals and regions that link to the task^2^, which is known as task-evoked fMRI. As an alternative, resting-state fMRI can observe brain signals during rest and measure intrinsic functional organization without performing any tasks^3^. Both task-evoked and resting-state fMRIs have been widely used in clinical and epidemiological neuroscience research to explore the relationship between inter-individual variations in brain function and human traits. For example, resting-state functional abnormalities are frequently observed in neurological and psychiatric disorders, such as Alzheimer’s disease^4^, attention-deficit/hyperactivity disorder (ADHD)^5^, schizophrenia^6^, and major depressive disorder (MDD)^7^. fMRI has also been used to identify the influence of multi-system diseases and complex traits, such as diabetes^8^, alcohol consumption^9^, and dietary behaviors^10^, on brain functions.

A major limitation of most fMRI association studies has been their small sample size, which is usually less than one hundred or a few hundred. Comparatively to structural magnetic resonance imaging (sMRI) measures, functional connectivity measures are generally noisier and show larger intra-subject variations^11^. Consequently, it may be difficult to replicate fMRI-trait associations found in small studies^12^. This problem can be resolved statistically by increasing the sample size of fMRI studies, which can detect weaker signals and reduce the uncertainty of the results. For example, Marek, et al. ^12^ showed that when the sample size is larger than 2,000, brain-behavioral phenotype associations can become more reproducible. However, the high assessment costs of fMRI may make it difficult to increase sample sizes sufficiently to collect the necessary data in every study. In the last few years, several large-scale fMRI datasets involving over 10,000 subjects have become publicly available, including the Adolescent Brain Cognitive Development^13^ (ABCD), the Chinese Imaging Genetics (CHIMGEN)^14^, and the UK Biobank^15^ (UKB). Particularly, the UKB study collected a rich variety of human traits and disease variables^16^, providing the opportunity to discover and validate fMRI-trait associations in a large-scale cohort.

Based on fMRI data from more than 40,000 subjects in the UKB study, we investigated resting-state and task-evoked functional architectures and their associations with human traits and health outcomes. By processing raw fMRI images from the UKB study, we represented the brain as a functional network containing 360 brain areas in a parcellation^17^ developed using the Human Connectome Project^18^ (HCP) data (referred to as the Glasser360 atlas, **Fig. 1, Fig. S1**, and **Table S1**). The Glasser360 atlas contained 64,620 full correlation measures to represent the functional connections among brain areas, providing fine-grained details of functional architecture over 12 functional networks^19^: the primary visual, secondary visual, auditory, somatomotor, cingulo-opercular, default mode, dorsal attention, frontoparietal, language, posterior multimodal, ventral multimodal, and orbito-affective networks. We performed a systematic analysis with 647 traits and diseases (selected to represent a wide range of traits and health conditions) using a discovery-validation design. Functional brain regions and networks were found to be strongly associated with a range of disorders and complex traits, including depression, risk-taking, cognitive traits, the use of electronic devices, physical activity, and atrial fibrillation. We also explored the differences between resting-state and task-evoked functional architectures, as well as age and sex-related effects. In order to evaluate how the choice of parcellation may impact our results, we additionally applied another parcellation^20^ on the same datasets, which divided the brain into 200 regions, referred to as the Schaefer200 atlas (**Fig. S2** and **Table S2**). We found that the two parcellations can yield similar conclusions and patterns, whereas the Glasser360 atlas can provide more biological insights due to its finer partitioning. The results of our trait-fMRI association studies have been made publicly available, and a browser tool has been developed to facilitate exploring the data (http://165.227.92.206/).

**Fig. 1.**
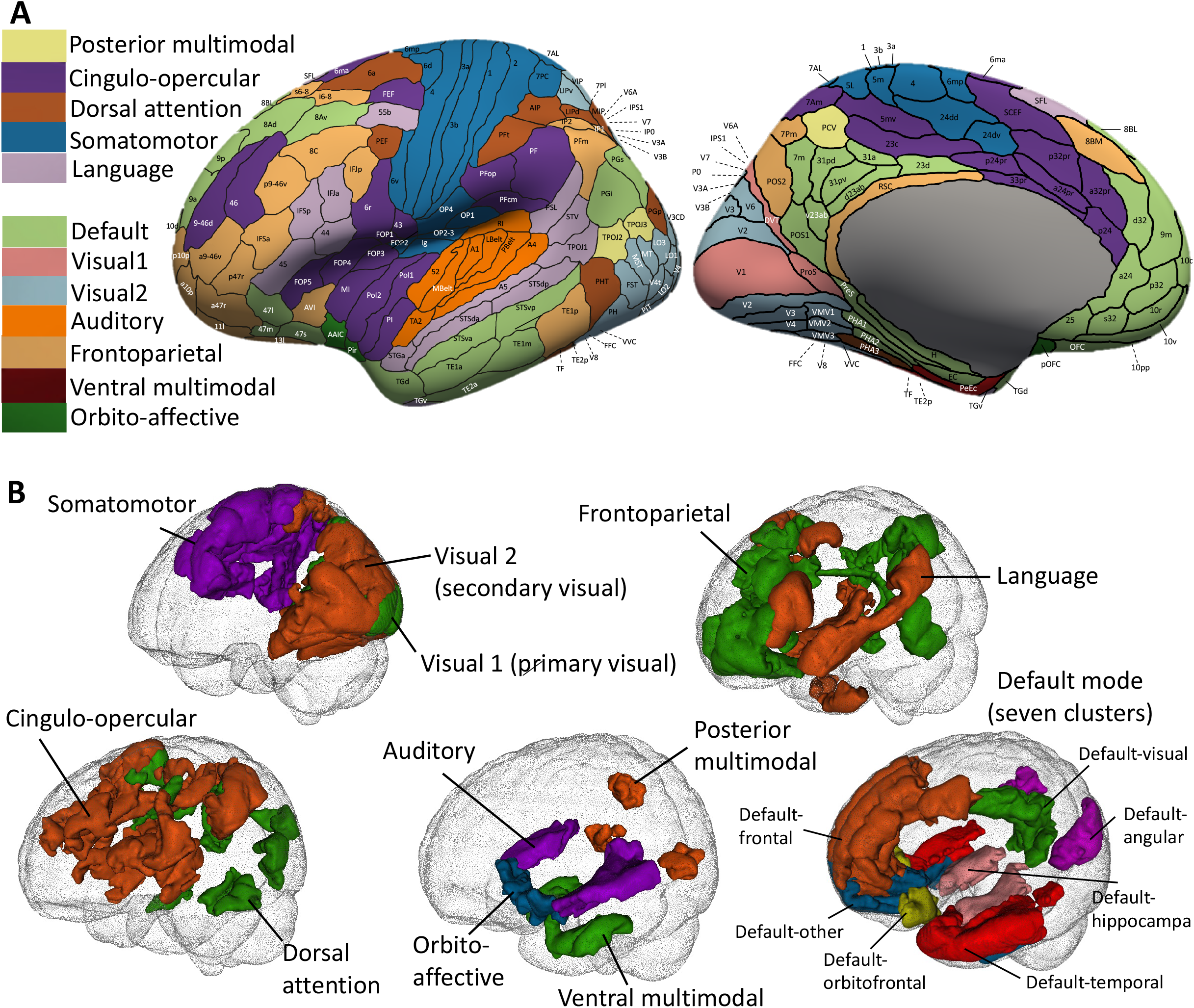
Illustration of functional areas and networks in the Glasser360 atlas. **(A)** Functional areas defined in the Glasser360 atlas (left hemisphere). See Table S1 for information of these areas and Figure S1 for maps of the whole brain (both hemispheres). Visual1, the primary visual network; Visual2, the secondary visual network. **(B)** Annotation of the 12 functional networks in the human brain. The default mode network (bottom right) is further divided into seven clusters, mainly based on their physical locations. See Figure S11 for more information of the seven clusters.

## RESULTS

### Consistency and reproducibility of the cerebral cortex functional organizations

First, we examined the consistency and reproducibility of functional connectivity using annotations from the Glasser360 atlas in the UKB study. As in Glasser, et al. ^17^, we first compared the group means of two independent sets of UKB subjects: the UKB phases 1 and 2 data (imaging data released up through 2018^21^, *n* = 17,374 for resting and 15,891 for task) and the UKB phase 3 data (data released in early 2020, *n* = 16,852 for resting and 13,232 for task, removing the relatives of subjects in early released data). **Figures S3-S4** illustrate the consistent spatial patterns of functional connectivity across the two independent groups. Similar to previous studies of other datasets^13,17,22^, the group mean maps in the two independent datasets of the UKB study were highly similar, with the correlation across the 64,620 (360 × 359/2) functional connectivity being 0.996 in resting fMRI and 0.994 in task fMRI. These results may suggest that the HCP-trained Glasser360 atlas can provide a set of well-defined and biologically meaningful brain functional traits that are generalizable across datasets.

Next, we evaluated the intra-subject reproducibility of the Glasser360 atlas using the repeat scans from the UKB repeat imaging visit (*n* = 2,771 for resting and 2,014 for task, average time between visits = 2 years). We performed two analyses. The first analysis is to compare the group mean maps of the original imaging visit to those of the repeat visit. Group means were highly consistent between the two visits, with correlation of 0.997 and 0.994 for resting and task fMRIs, respectively (ranges across different networks were [0.995, 0.999] for resting and [0.987, 0.998] for task, **Figs. S5-S6**). The second analysis quantified individual-level differences between the two visits. Specifically, we evaluated the reproducibility of each functional connectivity by calculating the correlation between two observations from all revisited individuals. Overall, the average reproducibility was 0.37 (standard error = 0.11) for resting fMRI and 0.31 (standard error = 0.08) for task fMRI (**Figs. 2A-B**). The reproducibility of within-network connectivity was generally high in resting fMRI (**Fig. 2C**, mean = 0.46). During task fMRI, the overall reproducibility was decreased (mean = 0.32) and the secondary visual and posterior multimodal networks exhibited higher functional connectivity on average than others. In addition, the connectivity within activated functional areas (defined by group-level *Z*-statistic maps, **Supplementary Note**) showed higher reproducibility than that within nonactivated areas (**Fig. 2D** and **Fig. S7A**, mean = 0.40 vs. 0.30, *P* < 2.2 × 10^−16^). The majority of the above-defined activations occurred in the secondary visual, dorsal attention, and somatomotor networks (**Fig. S8**). Furthermore, we examined the reproducibility of amplitude measures of fMRI^23,24^, which quantified the functional activity within each of the 360 brain areas. The average amplitude reproducibility was 0.60 (standard error = 0.08) for resting fMRI and 0.45 (standard error = 0.07) for task fMRI (**Fig. 2E**). In accordance with the findings in functional connectivity, the reproducibility of amplitude measurements of activated areas in task fMRI was higher than that of nonactivated areas (**Fig. 2F**, mean = 0.49 vs. 0.43, *P* = 1.1 × 10^−12^).

**Fig. 2.**
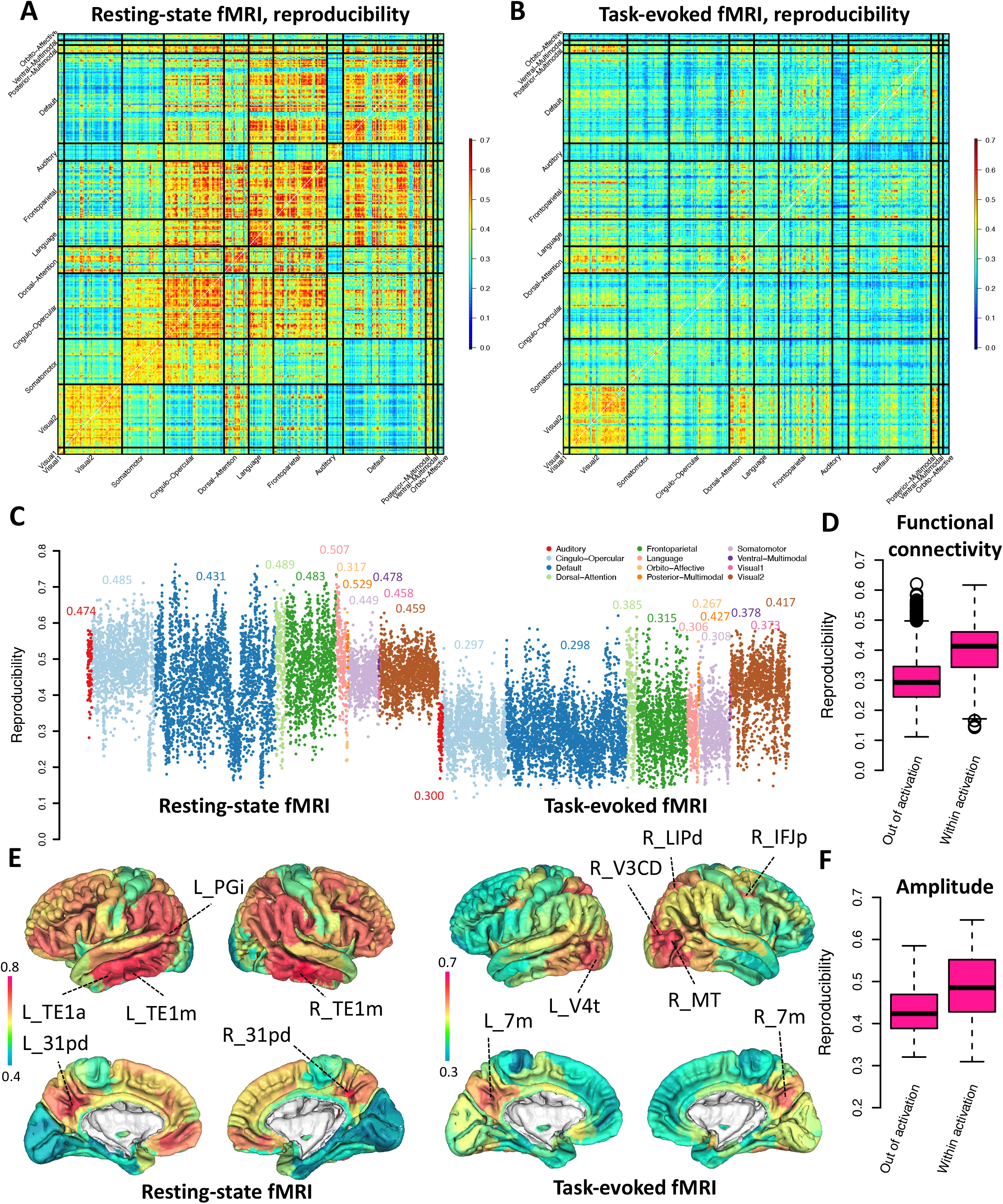
Distribution of reproducibility across brain functional areas and networks. We illustrate the spatial maps of reproducibility of functional connectivity for resting fMRI in **(A)** and task fMRI in **(B). (C)** Comparison of reproducibility of functional connectivity across 12 brain functional networks in resting (left panel) and task (right panel) fMRI. **(D)** Comparison of reproducibility of functional connectivity between the activated areas (within activation) and the nonactivated areas (out of activation) in task fMRI. **(E)** Comparison of reproducibility of amplitude measures in resting (left panel) and task (right panel) fMRI. See Table S1 for information of the labeled brain areas. **(F)** Comparison of reproducibility of amplitude measures between the activated areas (within activation) and the nonactivated areas (out of activation) in task fMRI. The activation map can be found in Figure S8.

Finally, we compared the spatial patterns of UKB and HCP studies. The correlation between UKB and HCP was 0.90 for resting fMRI and 0.78 for task fMRI in the group mean analysis (**Fig. S9**). These results demonstrate a substantial level of overall consistency between the typical subjects in a healthy young adult cohort and those of middle age and older age. Next, we examined the reproducibility of functional connectivity in the Glasser360 atlas using the repeated scans in HCP study (*n* = 1075, average time between two scans = 1 day). The average reproducibility was 0.40 (standard error = 0.09) for resting fMRI and 0.22 (standard error = 0.11) for task fMRI (the emotion task) (**Fig. S7B**). These results show that the two studies have similar reproducibility, suggesting that the quality of fMRI traits in the biobank-scale UKB study is comparable to that of the HCP project. Similar to the UKB study, the connectivity among activated functional areas (defined by group-level *Z*-statistic maps, **Supplementary Note**) had higher reproducibility than the nonactivated connectivity in HCP task fMRI (**Fig. S7C**, mean = 0.382 vs. 0.225, *P* < 2.2 × 10^−16^). In general, the excellent group mean map consistency, as well as the similar reproducibility between the UKB and the HCP studies, provides confidence that the Glasser360 atlas will be able to consistently annotate the functional organization of typical subjects in a healthy population. On the other hand, the relatively low intra-subject reproducibility of fMRI matches previous findings^11^, may suggest that a large sample size is needed to produce reproducible association results in downstream analyses^12^.

### Comparison of resting-state and task-evoked functional architectures

Understanding how the brain changes its functionality in response to tasks/stimuli is of great interest and has a wide range of clinical applications^25^. For example, fMRI studies with an emotional task consistently showed abnormalities in the prefrontal cortex-limbic area in patients with anxiety disorders, who tend to overreact to emotional stimuli^26^. Based on relatively small sample sizes, previous literature has found that intrinsic and extrinsic functional architectures are highly similar, with small but consistent differences across a range of tasks^27-33^. Using parcellation-based data from the large-scale UKB study, we uncover more details about resting-task functional connectivity differences.

The correlation between resting fMRI and task fMRI group mean maps was 0.754 in the UKB study and 0.782 in the HCP study, indicating the high degree of similarity between intrinsic and extrinsic functional architectures (**Fig. S9**). We found that the auditory and default mode networks exhibited the greatest resting-task differences. In the auditory network, task fMRI revealed stronger intra-hemispheric connections than resting fMRI, while the inter-hemispheric connections in task fMRI generally weakened or remained unchanged (**Fig. S10**). Task-related changes were more complex in the default mode network. To summarize the patterns, we grouped the 77 areas in the default mode network into seven clusters, mainly based on their physical locations (**Fig. S11**). We found that functional connectivity within the frontal, visual, and hippocampal clusters was stronger in task fMRI than in resting fMRI, while the connectivity between the frontal and the other two clusters decreased (**Fig. S12**). Moreover, the frontal cluster of default mode network can be further divided into two subclusters, the first subcluster consisted of left/right 9a, 9m, 9p, 8BL, 8Ad, and 8Av areas, mainly in the dorsolateral superior frontal gyrus (referred to as the dorsolateral superior subcluster); and the second one included left/right 10v, 10r, p32, a24, and 10d areas in the medial orbital superior frontal gyrus and pregenual anterior cingulate cortex (referred to as the medial orbital superior subcluster). The dorsolateral superior subcluster had decreased connectivity with the areas in other clusters of the default mode network in task fMRI, especially those in the temporal cluster. On the other hand, the medial orbital superior subcluster had a greater level of connectivity with a few other areas of the default mode network when performing the task, especially with the orbitofrontal complex (OFC) cluster and the neighboring 10pp area. Furthermore, the visual cluster maintained strong intra-cluster connectivity during the task, whereas its connectivity with the angular, frontal, and temporal clusters decreased. Although the default mode network has been originally recognized as brain areas with greater connectivity in resting fMRI than task fMRI^34^, recent studies have found that the default mode network also had positive functional contributions to tasks, which may result in increased activity in task fMRI^35^. Our results provided further insight into the complicated task-positive and task-negative functional connectivity change patterns in this network.

Several areas of the secondary visual network were less connected to other visual areas when the task was performed, including the left/right V6A (in the superior occipital), V6 (in the cuneus), VMV1 (in the lingual gyrus), and VMV2 (in the lingual and fusiform gyrus) (**Fig. S13**). Interestingly, some of these visual areas, such as the left/right V6, had increased functional connectivity with the default mode network (**Fig. S14**). There was also an increase in connections between the default mode network and other major cognitive networks, such as the cingulo-opercular and frontoparietal (**Fig. S15**). For the somatomotor network, the insula-related areas (including left/right Ig, FOP2, OP2-3, and right RI) had reduced connections with other somatomotor areas in task fMRI (**Fig. S16**). Similar to the auditory network, the inter-hemispheric connectivity in the cingulo-opercular network decreased in task fMRI (**Fig. S17**). Additionally, we found that the dorsal attention, frontoparietal, and language networks had similar functional connectivity patterns in resting and task fMRI (**Figs. S18-S20**). In summary, our results confirm the similarity of functional structures between resting and task fMRI, while also identifying specific patterns of differences.

### Age effects and sex differences in functional architectures

By using the large-scale fMRI data, we quantified the age and sex effect patterns on resting and task functional organizations (Methods). Several studies have examined the effects of age and differences between males and females on brain structures and functions, but the locations and patterns of the reported differences may vary across studies^36,37^. We used unrelated white British subjects in UKB phases 1-3 data release (until early 2020) as our discovery sample (*n* = 33,795 for resting and 28, 907 for task) and validated the results in an independent hold-out dataset, which included non-British subjects in UKB phases 1-3 data release and all subjects in UKB phase 4 data release (early 2021 release, removed the relatives of our discovery sample, *n* = 5, 961 for resting and 4, 884 for task). The full list of the adjusted covariates can be found in the Methods section. We reported the results passing the Bonferroni significance level (7.73 × 10^−7^ = 0.05/64,620) in the discovery dataset and being significant at the nominal significance level (0.05) in the validation dataset.

There were widespread age effects on functional connectivity of resting and task fMRI, and network and area-specific details were revealed (**Figs. 3A-B**). For example, as age increased, the connections within the auditory, secondary visual, somatomotor, language, and cingulo-opercular networks were generally weaker (**Figs. S21A-E**). Some areas had particularly large age-effects, such as the left/right PoI2 (the posterior insular area 2) areas in the cingulo-opercular network. However, both positive and negative age effects were observed in the frontoparietal and default mode networks (**Figs. S21F** and **S22**). Some areas had a greater degree of aging effects, such as the left/right POS2 (the parieto-occipital sulcus area 2) areas in the frontoparietal network and left/right POS1 (the parieto-occipital sulcus area 1) areas in the default mode network. Negative age effects in the default mode network were strongest in the hippocampal cluster, such as the left/right PHA1 (the parahippocampal area 1) areas.

**Fig. 3.**
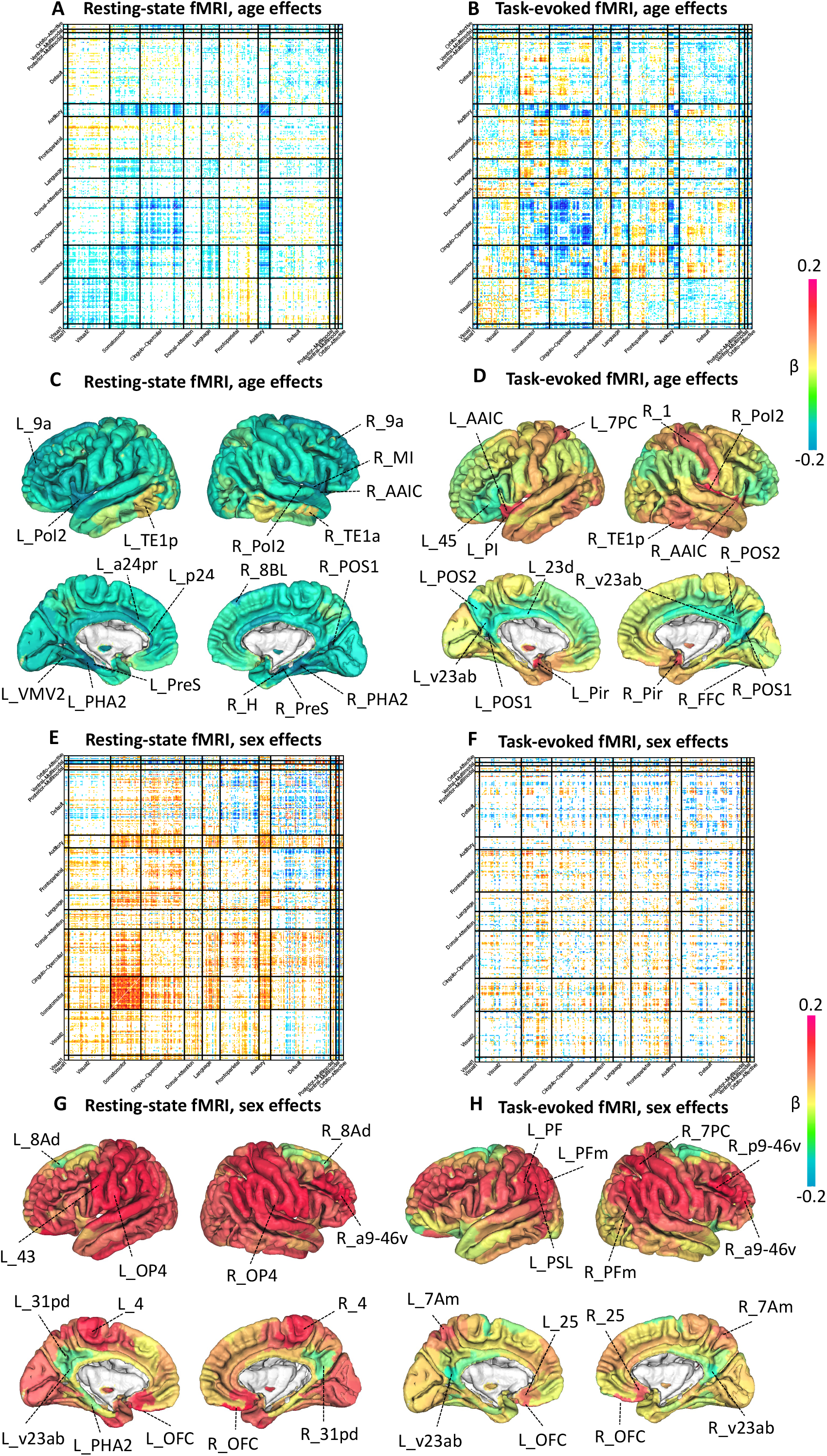
Spatial pattern of age and sex effects on brain functional organizations. We illustrate the spatial pattern of age effects (after adjusting for covariates) on functional connectivity for resting fMRI in **(A)** and for task fMRI in **(B). (C)** and **(D)** display the spatial pattern of age effects on amplitude measures of resting and task fMRI, respectively. See Table S1 for information of the labeled brain areas. We illustrate the spatial pattern of sex effects (after adjusting for covariates) on functional connectivity for resting fMRI in **(E)** and for task fMRI in **(F). (G)** and **(H)** display the spatial pattern of sex effects on amplitude measures of resting and task fMRI, respectively. We labeled the brain areas with the strongest age and sex effects in amplitude measures. For functional connectivity, we illustrated the effects passing the Bonferroni significance level (7.73 × 10^−7^, 0.05/64,620) in the discovery dataset (*n* = 33,795 for resting and 28, 907 for task) and being significant at the nominal significance level (0.05) in the validation dataset (*n* = 5, 961 for resting and 4, 884 for task).

In task fMRI, age effects were different from those in resting fMRI. We highlighted a few patterns. First, the age effects in the auditory network were mainly on the inter-hemispheric connections, where the connectivity between the left and right hemispheres decreased with aging (**Fig. S23A**). Similarly, the inter-hemispheric connectivity between the auditory and cingulo-opercular networks declined as we aged. The age effects on intra-hemispheric connections were much weaker. Except for a few areas (such as the right 8Ad and right PEF), most areas in the cingulo-opercular and default mode networks had reduced functional connectivity with aging (**Fig. S23B** and **S24**). On the other hand, most of the functional connectivity in the secondary visual network increased with aging, especially the left/right V3A and V6A areas in the superior occipital gyrus (**Fig. S23C**). There were both positive and negative effects of aging on other networks, such as the somatomotor, frontoparietal, and dorsal attention (**Figs. S23D-F**). Overall, these results describe the detailed age effect pattern for functional organizations at rest and during task performance.

We also examined the age effects on amplitude measures. In resting fMRI, age-related decreases in brain activity were observed in most brain areas, with the strongest effects in left and right PreS areas (the presubiculum, a subarea of the parahippocampal region, *β* < -0.222, *P* < 5.01 × 10^−193^, **Fig. 3C**). In task fMRI, however, both strong positive and negative effects on brain activity were widely observed (**Fig. 3D**). Because widespread age effects were detected on both functional connectivity and amplitude traits, we examined the conditional age effects on functional connectivity traits after additionally including amplitude traits as covariates. After adjusting for amplitude traits, most of the age effects on functional connectivity traits became much smaller and were not significant at the Bonferroni significance level, especially in the resting fMRI (**Fig. S25**). For example, although a few of the strongest amplitude-adjusted age effects remained significant, most of the other moderate amplitude-adjusted age effects failed to pass the Bonferroni significance level in the default mode network (**Fig. S26**). Overall, these results for amplitude traits indicate that age has a significant effect on the variation of amplitude traits across subjects, which may also be carried over to functional connectivity traits^23^.

Functional connectivity patterns differed between males and females. We found widespread sex differences across different resting fMRI networks, with the strongest differences occurring in the somatomotor network (**Fig. 3E**). Males had stronger functional connectivity in the somatomotor and auditory networks as well as a few specific areas, including the left/right VIP (in the superior parietal gyrus), LIPv (in the superior parietal gyrus), PH (in the inferior temporal gyrus), and V6A (in the superior occipital gyrus) of the secondary visual network, the left/right PFcm (in the superior temporal gyrus) and 43 (in the rolandic operculum) of the cingulo-opercular network, the left/right a9-46v and p9-46v (both in the middle frontal gyrus) of the frontoparietal network, and the left/right PGp (in the middle occipital gyrus) of the dorsal attention network (**Figs. S27A-F**). In the default mode network, the sex difference had a complicated pattern. Specifically, males had stronger connectivity in the hippocampal and OFC clusters, especially in the left 47m area of the posterior orbital gyrus. On the other hand, females had stronger connectivity in many other areas of the default mode network (**Fig. S28**).

The sex differences in task fMRI were more specific to particular brain areas, including the right V6A (in the superior occipital gyrus) and left VMV2 (in the lingual and fusiform gyrus) of the secondary visual network, left/right PHA3 (in the fusiform gyrus) of the dorsal attention network, and left/right RSC (in the middle cingulate cortex) of the frontoparietal network (**Fig. 3F** and **S29A-C**). Males had stronger functional connectivity than females in most areas of the language, auditory, and somatomotor networks (**Figs. S29D-F**). Additionally, males had stronger connectivity in the hippocampal and frontal areas of the default mode network, whereas females had stronger connectivity between the visual cluster and the frontal cluster (**Fig. S30**). As for the amplitude measures, females had stronger brain activity in many areas of the default mode network, whereas males had stronger brain activity in most other networks in resting fMRI (**Fig. 3G**). Sex differences were generally reduced in task fMRI amplitude measurements (**Fig. 3H**). Lastly, we estimated the amplitude-adjusted sex effects on functional connectivity traits by additionally controlling for the amplitude traits as covariates. Similar to the findings of the age effects, the majority of amplitude-adjusted sex effects on functional connectivity traits can be explained by amplitude traits, such as in the somatomotor and default mode networks (**Fig. S31-S32**). In summary, as the fMRI traits of the brain is strongly associated with cognitive impairment and functional abnormalities, our area- and network-specific sex effect maps can be useful for understanding sex differences in brain disorders, such as Alzheimer’s Disease^38^ and depression^39^.

### An atlas of trait associations with cerebral cortex functional areas

In this section, a total of 647 phenotypes (selected to cover a wide range of traits and diseases) were examined for their associations with resting and task-functional organizations (Methods). Similar to the age and sex analyses, we used unrelated white British subjects in UKB phases 1-3 data release as the discovery sample (*n* = 33,795 for resting and 28, 907 for task) and validated the results in an independent hold-out dataset (*n* = 5, 961 for resting and 4, 884 for task). Detailed information on the adjusted covariates can be found in the Methods section. We prioritized significant associations that survived at the false discovery rate (FDR) level of 5% (by the Benjamini-Hochberg procedure) in the discovery sample and remained significant at the nominal significance level (0.05) in the validation sample. Among the 647 traits, 120 had at least one significant association with resting fMRI functional connectivity measures, among which 82 further survived the Bonferroni significance level (7.73 × 10^−7^, 0.05/64,620) (**Table S3**). We highlighted below the association patterns with mental health, cognitive function, physical activity, lifestyle, biomarkers, and disease status.

We observed strong associations between resting fMRI and multiple mental health traits, including risk-taking, depression, MDD, and neuroticism. Enrichments in specific networks and brain areas were observed. For example, risk-taking (Data field 2040) was strongly positively associated with the somatomotor network and the connections between the somatomotor and visual networks (**Fig. 4A**). Risk-taking was also negatively associated with the functional connections of the default mode network. Functional connectivity of sensory/motor areas was recently found to be positively associated with risk-taking^40^ and our findings were consistent with the “sensory-motor-cognitive” mode of brain functional amplitude changes related to aging^41^. In addition, depression was mostly associated with reduced connectivity in the somatomotor and cingulo-opercular networks (curated disease phenotype based on ICD-10 codes, **Fig. 4B**). Consistent patterns were also observed in MDD (ICD-10 code F329, **Fig. S33A**), nervous feelings (Data field 1970, **Fig. S33B**), seen doctor for nerves anxiety tension or depression (Data field 2090, **Fig. S33C**), neuroticism score (Data field 20127, **Fig. S33D**), and suffer from nerves (Data field 2010, **Fig. S33E**). Depression and depressive mood disorders have been linked to the abnormal brain connectivity in various intrinsic networks^42-44^, our results highlighted the specific patterns of the decreased resting functional connectivity, particularly in the somatomotor network.

**Fig. 4.**
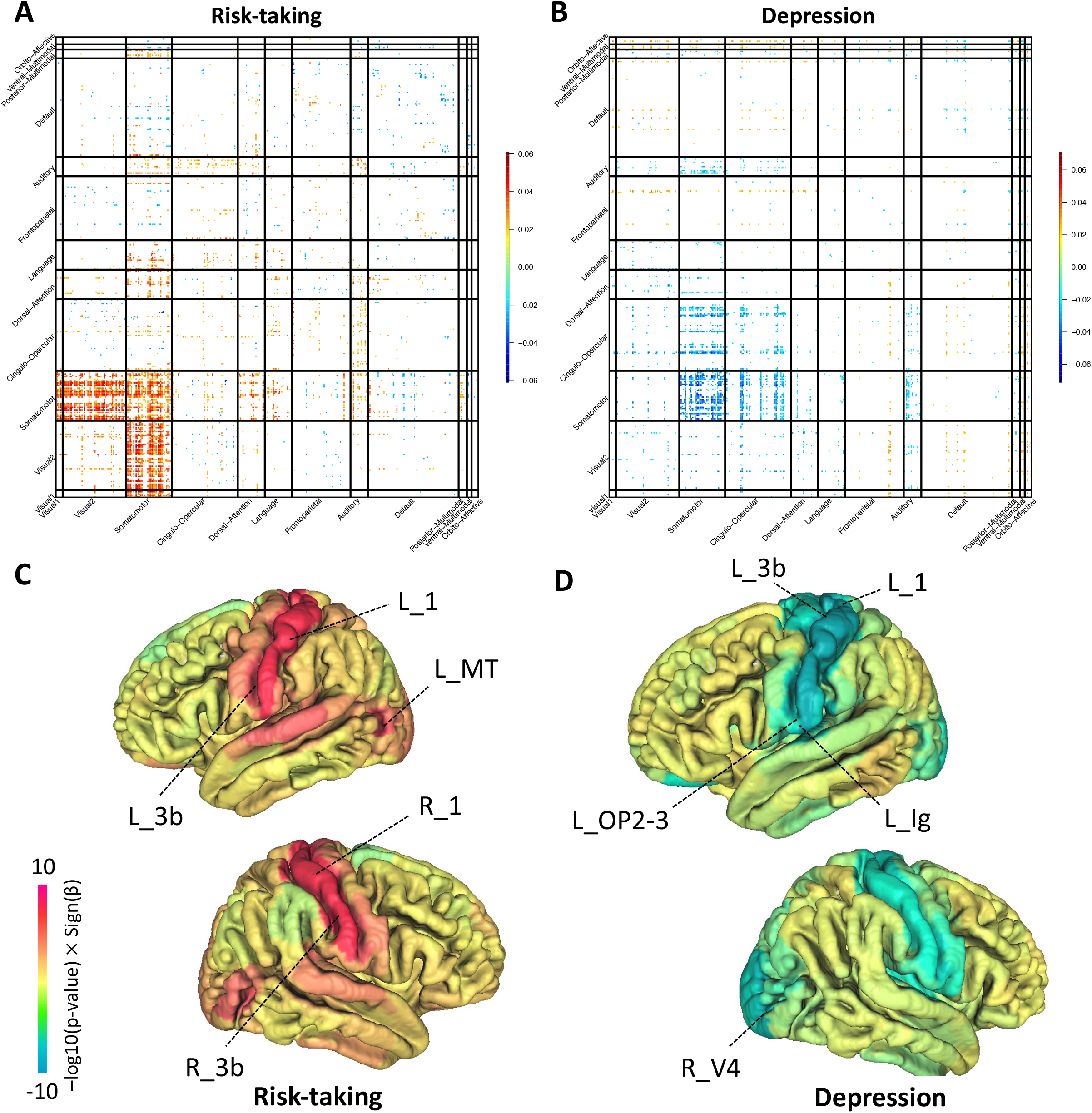
Selected complex traits that were associated with brain functional organizations. **(A)** Associations between risk-taking (Data field 2040) and functional connectivity of resting fMRI. This figure and the top-ranked brain areas can be viewed in an interactive version at http://165.227.92.206/trait/trait85.html. **(B)** Associations between depression (curated disease phenotype) and functional connectivity of resting fMRI. This figure and the top-ranked brain areas can be viewed in an interactive version at http://165.227.92.206/trait/trait230.html. We illustrated the estimated correlation coefficients that were significant at FDR 5% level in the discovery sample (*n* = 33,795) and were also significant at the nominal significance level (0.05) in the validation dataset (*n* = 5, 961). **(C)** and **(D)** display the spatial pattern of associations with amplitude measures of resting fMRI for risk-taking and depression, respectively. Brain areas with the strongest associations were labeled. See Table S1 for information of these areas.

A wide range of cognitive traits were associated with functional connectivity in fMRI, such as the fluid intelligence (Data field 20016), the number of puzzles correctly solved (Data field 6373), duration to complete alphanumeric path (Data field 6350), and maximum digits remembered correctly (Data field 4282). These cognitive traits showed different association patterns. Fluid intelligence, for example, was associated with functional connectivity in the auditory, language, cingulo-opercular, dorsal attention, and default mode networks, most of the associations were positive (**Fig. 5A**). The duration to complete alphanumeric path was mainly negatively associated with functional connectivity in the secondary visual network (**Fig. S34A**), the number of puzzles correctly solved was mostly related to the functional connectivity within the default mode, somatomotor, and secondary visual networks (**Fig. S34B**), and the maximum digits remembered correctly was positively related to the auditory and language networks (**Fig. S34C**). We also uncovered the association pattern for other brain-related complex traits, such as the strong connections between handedness (Data field 1707) and the cingulo-opercular network (**Fig. S34D**).

**Fig. 5.**
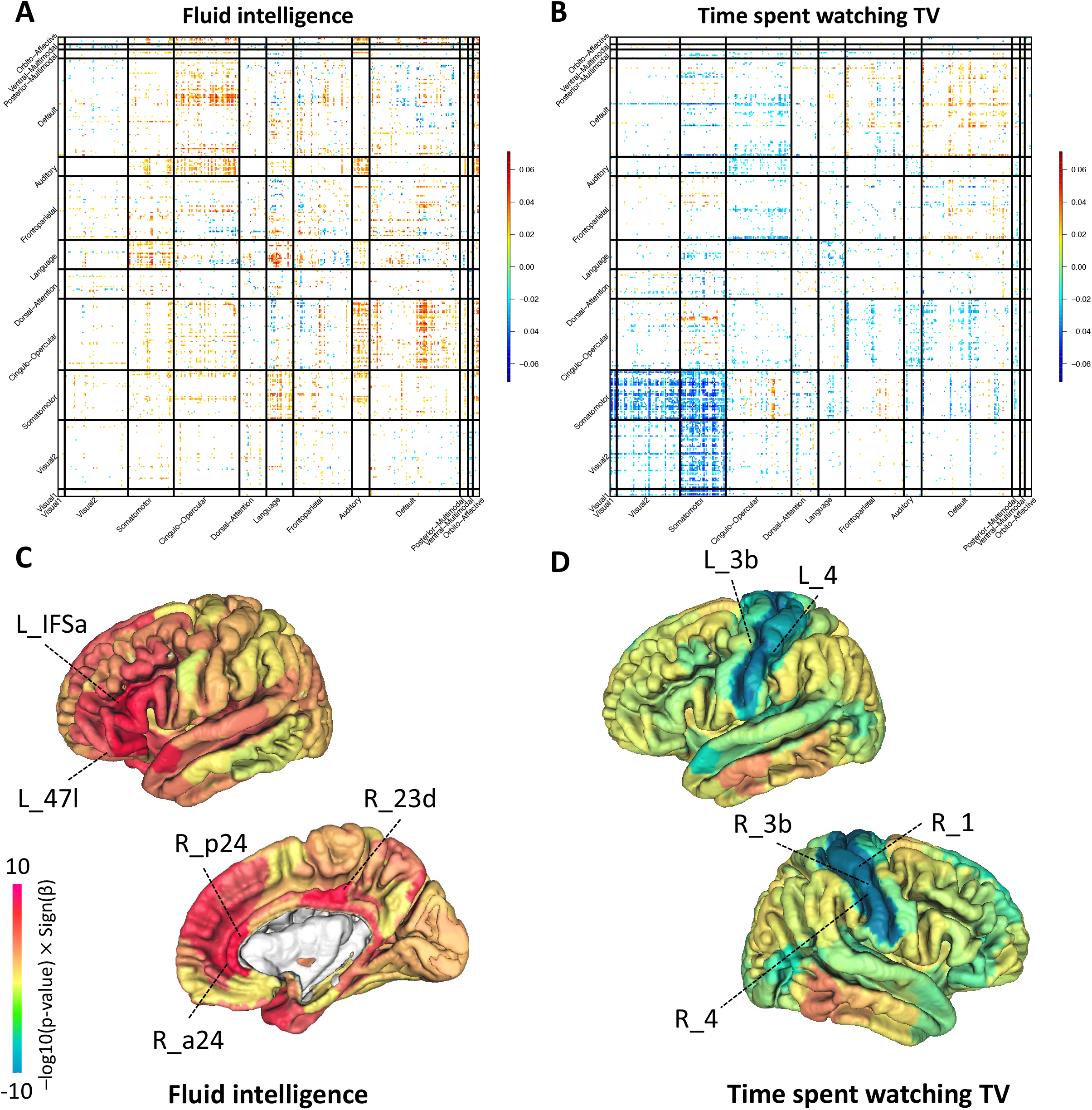
Selected complex traits that were associated with brain functional organizations. **(A)** Associations between fluid intelligence (Data field 20016) and functional connectivity of resting fMRI. This figure and the top-ranked brain areas can be viewed in an interactive version at http://165.227.92.206/trait/trait158.html. **(B)** Associations between time spent watching TV (Data field 1070) and functional connectivity of resting fMRI. This figure and the top-ranked brain areas can be viewed in an interactive version at http://165.227.92.206/trait/trait101.html. We illustrated the estimated correlation coefficients that were significant at FDR 5% level in the discovery sample (*n* = 33,795) and were also significant at the nominal significance level (0.05) in the validation dataset (*n* = 5, 961). **(C)** and **(D)** display the spatial pattern of associations with amplitude measures of resting fMRI for fluid intelligence and time spent watching TV, respectively. Brain areas with the strongest associations were labeled. See Table S1 for information of these areas.

Resting functional connectivity was widely associated with lifestyle and environmental traits, including physical activity, electronic device use, smoking, diet, alcohol, and sun exposure. Similar to risk-taking, mobile phone usage-related traits (Data fields 1120, 1140, and 1110) were consistently positively associated with the somatomotor network and connections between the somatomotor and visual networks (**Figs. S35A-C**). Watching television (TV) for longer periods of time (Data field 1070) may weaken functional connectivity in the somatomotor and visual networks as well as strengthen functional connectivity in the default mode network (**Fig. 5B**). TV viewing has been found to be associated with brain structural variations in visual cortex and sensorimotor areas^45^. Moreover, longer time spent outdoors in summer (Data field 1050) was associated with increased functional connectivity in the default mode network (**Fig. S35D**). These results may indicate that the default mode network is related to outdoor exploration and sunlight exposure.

We found associations between resting fMRI and multiple biomarkers, such as the basal metabolic rate (Data field 23105), albumin (Data field 30600), total protein (Data field 30860), and vitamin D (Data field 30890). For example, the basal metabolic rate was associated with increased functional connectivity in the somatomotor network and reduced functional connectivity in the default mode network (**Fig. S36A**). Higher levels of albumin and total protein were mainly associated with reduced functional connectivity in the somatomotor and visual networks (**Figs. S36B-C**). Human albumin is the most abundant protein, and low serum albumin may increase the risk of Alzheimer’s disease^46^. In addition, vitamin D was associated with increased functional connectivity, especially in the cingulo-opercular and somatomotor networks (**Fig. S36D**). Vitamin D is important for maintaining brain health, and vitamin D deficiency has been associated with the development of dementia, depression, and other mental illnesses^47^.

Strong associations between increased functional connectivity and cardiovascular diseases were identified, including the atrial fibrillation (curated disease phenotype and ICD-10 code I48), vascular/heart problems diagnosed by doctor (Data field 6150), and hypertension (curated disease phenotype and ICD-10 code I10). Atrial fibrillation is the most common clinically significant arrhythmia, and increasing evidence suggests it is associated with cognitive decline and dementia^48^. We found that atrial fibrillation was widely associated with functional connectivity across different networks (**Figs. S37A-B**). Hypertension and vascular/heart problems were associated with reduced functional connectivity in the auditory, somatomotor, secondary visual, and cingulo-opercular networks (**Figs. S37C-D**). Hypertension is a major risk factor of vascular dementia and Alzheimer’s Disease and altered functional connections may reflect the early effects of vascular risk factors on brain functions^49^.

In task fMRI, 96 traits had at least one significant association at the FDR 5% level (and significant at the nominal level in the validation dataset), and 59 further survived the Bonferroni significance level (7.73 × 10^−7^ = 0.05/64,620) (**Table S3**). Of the 96 traits, 69 were also significantly associated with resting fMRI at the 5% FDR level. The association patterns in task and resting fMRI were very similar for a few traits, such as the atrial fibrillation (**Fig. S38**). For many traits, however, we observed different patterns in resting and task fMRI, including fluid intelligence (**Figs. S39A-B**), the number of puzzles correctly solved (**Figs. S39C-D**), time spent outdoors in summer (**Figs. S40A-B**), time spent watching TV (**Figs. S40C-D**), and basal metabolic rate (**Figs. S41A-B**). For example, both fluid intelligence and the number of solved puzzles were positively associated with intra-hemispheric connections of the auditory network in task fMRI, whereas no or negative associations were observed with inter-hemispheric connections. There were similar intra- and inter-hemispheric connection differences in the cingulo-opercular network. Overall, the results indicate differences between resting and task-related functional associations with complex traits, especially for cognitive functions.

Task fMRI also revealed new insights into the brain function associations with more traits, such as early life factors. Specifically, we found strong associations between task fMRI and the place of birth in UK (the north co-ordinate and east co-ordinate, Data fields 129 and 130) in the auditory, somatomotor, and cingulo-opercular networks (**Fig. S42A-B**). These results may shed light on the impact of the environment on brain development related to the emotion processing task. Additionally, we observed stronger associations with multiple biomarkers than in resting fMRI, such as the triglycerides (Data field 30870, **Fig. S42C**) and urate (Data field 30880, **Fig. S42D**). In contrast, task fMRI was not associated with a few traits that were strongly related to resting fMRI, including mental health traits (such as risk-taking and depression) and electronic device use (such as usage of mobile phone).

We also quantified the association patterns with amplitude traits and prioritized brain areas whose functional activity was related to traits and diseases. We observed similar patterns to the functional connectivity results. For example, risk-taking has the strongest associations with brain activity of the postcentral gyrus in the somatomotor network, especially the primary somatosensory cortex^40^ (**Fig. 4C**, *β* > 0.033, *P* < 8.14 × 10^−6^). The postcentral gyrus, insula, and Rolandic operculum areas of the somatomotor network were most negatively related to depression (**Fig. 4D**, *β* < -0.036, *P* < 7.10 × 10^−7^). All significant associations with fluid intelligence were positive, with the top three areas being the middle cingulate, anterior cingulate, and orbital part of the inferior frontal gyrus (IFG pars orbitalis) in the default mode network (**Fig. 5C**, *β* > 0.053, *P* < 1.31 × 10^−12^). Time spent watching TV was strongly negatively associated with the postcentral gyrus, precentral gyrus, paracentral lobule, and the supplementary motor area in the somatomotor network (**Fig. 5D**, *β* < -0.050, *P* < 2.03 × 10^−12^). In summary, this section analyzes fMRI data with a variety of complex traits in a discovery-validation design. We provide new insights into the association maps with human brain resting and task functional organizations, which could assist in building better disease prediction models and selecting clinically useful neuroimaging biomarkers. The full set of results can be browsed at http://165.227.92.206/traitList.html.

### Alternative analyses using the Schaefer200 atlas

The brain parcellation may play a crucial role in the definition of the brain functional network and affect the results of downstream analysis^50^. To explore the impact of parcellation choice on the large-scale UKB study, we additionally applied another parcellation (the Schaefer200 atlas^20^) and repeated our analysis of on the same set of subjects. Briefly, the Schaefer200 atlas partitioned the brain into 200 regions, resulting in 19,900 pairwise functional full correlation measures (200 × 199/2). We mapped the 200 regions onto the same 12 networks used in the Glasser360 atlas (**Table S2**, Methods).

The average reproducibility in the Schaefer200 atlas was 0.387 (standard error = 0.10) for resting fMRI and 0.312 (standard error = 0.07) for task fMRI, which was in the same range as the Glasser360 atlas. **Figure S43** compares the reproducibility of the two parcellations. Glasser360 and Schaefer200 atlases showed similar patterns across a variety of networks, with the largest differences being observed in the secondary visual network, where the Glasser360 atlas was more reproducible. In addition, consistent spatial patterns of functional connectivity were observed in the two parcellations, although the strength of connectivity was slightly higher in the Schaefer200 atlas, which may partly be explained by the smaller number of brain areas (**Fig. S44**). These results demonstrate the good generalizability of functional organizations modeled by the Glasser360 atlas.

We evaluated the age and sex effects in the Schaefer200 atlas. **Figure S45** compares the age effect patterns in the Schaefer200 and Glasser360 atlases. In both atlases, decreasing resting functional connectivity was consistently associated with aging, especially in the auditory, cingulo-opercular, and somatomotor networks. The main difference was in the secondary visual network, where the age effects in the Glasser360 atlas were stronger than those in the Schaefer200 atlas (**Fig. S45A**). This finding may be attributed to the lower reproducibility of the Schaefer200 atlas in the secondary visual network, suggesting that the Glasser360 atlas may be more suitable for studying the brain connectivity of the visual cortex. In addition, consistent intra- and inter-hemispheric association differences in task fMRI were observed (**Fig. S45B**). The Schaefer200 and Glasser360 atlases also showed similar sex effect patterns, in which the strongest effects were both detected in the somatomotor and auditory networks (**Fig. S46**).

Next, we repeated the association analysis with the 647 traits. In resting fMRI, 131 traits had at least one significant association at the FDR 5% level and 83 further passed the Bonferroni significance level (2.51 × 10^−6^ = 0.05/19,900, also passing the nominal significance level (0.05) in the independent validation dataset, **Table S3**). Of the 120 traits with significant associations in the Glasser360 atlas analysis, 109 (90.83%) were also significant in the Schaefer200 atlas analysis. Additionally, the association maps were largely consistent in the two atlases. For example, time spent watching TV was consistently associated with decreased functional connections of the somatomotor and visual networks, as well as increased functional connectivity in the default mode network (**Fig. S47A**). Moreover, fluid intelligence was consistently linked to increased functional connectivity, particularly in the language and auditory networks (**Fig. S47B**). In both atlases, depression was associated with reduced functional connectivity in the somatomotor and cingulo-opercular networks (**Fig. S48**). At the FDR 5% level, 90 traits showed significant associations with task fMRI, including 76 of the 96 (79.2%) traits that were significant in the Glasser360 atlas analysis (**Table S3**). All these results are available on our website. In summary, the Schaefer200 atlas results agree well with those of the Glasser360 atlas, indicating that the patterns observed in our Glasser360 analysis are not parcellation-specific.

Finally, we examined the trait associations with 1,701 functional connectivity traits based on the whole brain spatial independent component analysis (ICA)^24,51,52^ approach in resting fMRI. These ICA functional connectivity traits were available from the UK Biobank data release (https://www.fmrib.ox.ac.uk/ukbiobank/index.html, Data fields 25752 and 25753), which were partial correlations and the timeseries were estimated from group ICA maps via the dual-regression^24^. Of the 647 traits, 76 demonstrated at least one significant association at the FDR 5% level and 58 remained significant at the Bonferroni significance level (2.94 × 10^−5^ = 0.05/1,701, also passing the nominal significance level in the independent validation dataset, **Table S3**). Among the 76 ICA-significant traits, 65 (85.53%) were also significant in the above Glasser360 atlas analysis. Compared to the ICA-derived traits, parcellation-based traits from the Glasser360 atlas (which identified significant associations with 120 complex traits at the FDR 5% level and 82 at the Bonferroni significance level) were able to detect associations with more traits.

In addition, we ranked the 58 ICA-significant complex traits (at the Bonferroni significance level) by the number of their significant associations with ICA-derived traits. Then we compared the association strengths of the top ten traits with ICA-derived traits and those with Glasser360 traits. On these ten traits, ICA-derived traits and Glasser360 traits showed similar levels of association strength (**Fig. S49**). For example, many ICA-derived and Glasser360 traits were found to be significantly associated with systolic blood pressure (Data field 4080), and most of these associations were in a similar range of effect size and *P* value (**Figs. S50-51**). Furthermore, the results of Glasser360 traits indicate that the auditory and somatomotor networks may be more strongly associated with systolic blood pressure than other networks. In summary, parcellation-based traits may reveal more network and area-level details with comparable association strength to ICA-derived traits.

### Fluid intelligence prediction by integrating multiple data types

Our association analyses demonstrate the potential value of large-scale fMRI data for a variety of complex traits and disorders in clinical and epidemiological research. For example, it is of great interest to construct prediction models by integrating fMRI data and other data types^53-55^. Fluid intelligence is a key indicator of cognitive ability and is associated with multiple neurological and neuropsychiatric disorders^56^. In this section, we performed prediction for fluid intelligence using neuroimaging traits from multiple modalities, including resting fMRI, task fMRI, diffusion MRI (dMRI)^21^, and structural MRI (sMRI)^57^. We further integrated these neuroimaging data with a wide range of other data types, including common genetic variants, biomarkers, local environments, early life factors, diet, and behavioral traits. The relative contributions and joint performance of these data types were assessed in a training, validation, and testing design (Methods). All model parameters were tuned using the validation data and we evaluated the prediction performance on the independent testing data by calculating the correlation between the predicted values and the observed intelligence, while adjusting for the covariates listed in the Methods section.

The prediction performance of multi-modality neuroimaging traits was summarized in **Figure 6A**. The prediction correlation of resting fMRI was 0.272 (standard error = 0.012), suggesting that about 7.4% variation in fluid intelligence can be predicted by resting fMRI connectivity. The prediction correlation was similar in task fMRI (correlation = 0.279) and was improved to 0.333 by jointly using resting and task fMRI, which suggests that resting and task fMRI had unique contributions to intelligence prediction. This improvement matched our association results where both resting and task fMRI showed strong associations with fluid intelligence with different spatial patterns. In addition, the dMRI and sMRI traits had much lower prediction accuracy than fMRI traits. Specifically, the prediction correlation was 0.09 for diffusion tensor imaging (DTI) parameters of dMRI and 0.08 for regional brain volumes of sMRI. Moreover, adding these structural traits in addition to fMRI traits did not substantially improve the prediction performance (correlation = 0.342), indicating the prediction power of brain structural traits for intelligence can be largely captured by the functional traits.

**Fig. 6.**
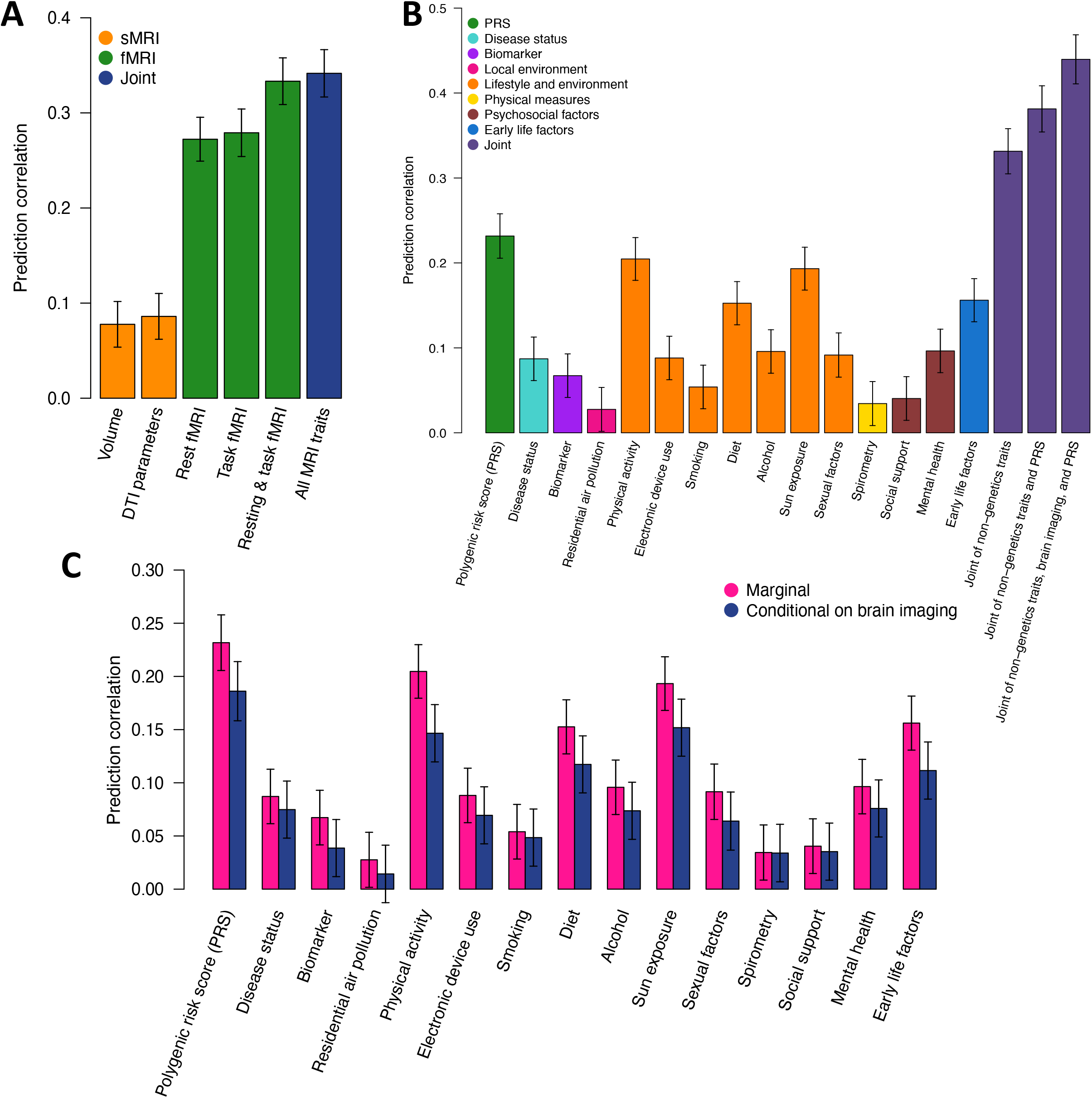
Integrative prediction model for fluid intelligence. **(A)** Prediction accuracy of neuroimaging traits for fluid intelligence. Volume, region brain volumes from brain structural MRI (sMRI); DTI parameters, diffusion tensor imaging parameters to measure brain white matter microstructures; All MRI traits, including brain volume, DTI parameters, resting fMRI, and task fMRI. **(B)** Prediction accuracy of non-neuroimaging traits from different trait categories and their joint performance. PRS, polygenic risk scores of genetic variants. **(C)** Comparison of predictive power of non-neuroimaging traits before (“marginal”) and after controlling for the neuroimaging traits (“conditional on brain imaging”).

Next, we examined the prediction performance of non-neuroimaging data types (**Fig. 6B**). The prediction correlation of intelligence genetic polygenic risk score (PRS) was 0.232 (standard error = 0.013), which was slightly lower than the performance of resting fMRI. Several categories of lifestyle and environmental traits had strong predictive power, including physical activity (correlation = 0.205), sun exposure (correlation = 0.193), and diet (correlation = 0.153). Moreover, biomarkers, disease records, and early life factors all had significant predictive performance, with prediction correlations being 0.067, 0.087, and 0.156, respectively. By combining all these non-neuroimaging data types, the prediction correlation increased to 0.381. The performance was further improved to 0.440 by including neuroimaging data, which was much higher than when using only one type of data.

To explore whether the predictive power of non-neuroimaging traits (such as physical activity) is mediated by brain structure and function, we evaluated their conditional predictive performance on fluid intelligence after controlling for neuroimaging traits. There was a reduction of performance on multiple categories of non-neuroimaging predictors, suggesting their effects on intelligence may be indirect and partially mediated by brain structure and function (**Fig. 6C** and **Table S4**). For example, the prediction performance of genetic PRS decreased from 0.232 to 0.186, indicating that 19.8% of the genetic predictive power on intelligence can be captured by brain structural and functional variations measured by brain MRI. The proportion was 28.3% for physical activity, 23.1% for diet, and 28.6% for early life factors. Overall, these results illustrate the neuroimaging traits, especially the ones from resting and task fMRI, are powerful predictors of cognitive function. Future studies can integrate genetic, biomarker, behavioral/environmental factors, and multi-modality MRI data for better prediction of brain-related complex traits and disorders.

## DISCUSSION

Inter-individual variations in brain function and their relationship to human health and behavior are of great interest. The intra-individual reproducibility of brain fMRI traits is generally lower than that of structural MRI traits, although the group-level consistency is high^11,13,22,58^. Then it has been suggested that a large sample size is needed for fMRI studies to detect trait associations with small effect sizes^59,60^. The UKB study provided an extensive biobank-scale data resource for quantifying fMRI associations with many phenotypes. The present study conducted a systematic analysis of intrinsic and extrinsic functional architectures with a parcellation-based approach using fMRI data collected from over 40,000 individuals. We measured the differences between resting and task fMRI, investigated age and sex effects on brain function, and examined the cross-parcellation variability of our findings. We evaluated the fMRI associations with 647 traits chosen from a variety of trait domains. In comparison to the prior literature^15^, which applied data-driven spatial independent component analysis^24,51,52^ to about 5000 subjects, the parcellation-based approach and much larger sample size allowed us to quantify functional organizations in fine-grained details. We found distinct brain functional areas and networks that were strongly related to traits from various categories, such as mental health, physical activity, cognitive performance, and biomarkers. We developed integrative prediction models for fluid intelligence, suggesting that integrating fMRI traits with multiple data types can improve prediction performance for brain-related complex traits and diseases.

We found that the strongest sex difference in resting fMRI was in the somatomotor network, where females had weaker functional connectivity than males (**Fig. 3E**). Additionally, depression was strongly associated with decreased connectivity in the somatomotor network (**Fig. 4B)**. Considering the fact that depression is two times more prevalent in females than in males, our results may help understand the brain function-related sex differences in depression^39^. In addition, we found that a wide variety of complex traits were strongly associated with the functional connectivity between the visual and somatomotor networks, including risk-taking, time spent watching TV, usage of mobile phone, albumin, and total protein (**Figs. 4A, 5B, S31A, S32B**, and **S32C**). Future studies could investigate the biological mechanisms underlying these functional connectivity alterations as well as causal medication pathways among lifestyle, biomarker, brain function, and mental health^61^.

Our results confirm that group-level intrinsic and extrinsic functional spatial patterns are largely similar (correlation = 0.754), as observed in previous fMRI datasets with smaller sample sizes^27-32^. The large-scale UKB data also revealed that resting and task fMRI may have different associations with complex traits, such as mental health and cognitive abilities. For example, depression was strongly associated with resting fMRI, but not with task fMRI. Moreover, in resting and task fMRI, the associations with fluid intelligence had different spatial distributions. Our prediction analysis further suggests that task fMRI has additional predictive power on intelligence on top of resting fMRI. These results demonstrate the differences between resting and task-evoked brain functions in terms of their connections with brain health and cognition.

The UKB task fMRI data used in this study were from a single emotion processing task^62,63^. Previous studies have shown that the functional architectures of different tasks were highly similar^27,29,30^. Hence, our findings from this specific task might be generalizable to other tasks. More insights might be revealed in future studies by integrating multiple neuroimaging data resources. For example, joint analysis with other large-scale neuroimaging studies, such as the ABCD study, may help understand the age-related interaction with complex traits across the lifespan. In addition, further investigations are needed to examine the effects of topographical misalignments on trait-fMRI associations and sex differences. There has been an observation in the HCP study that the cross-subject variability can be explained by the misalignment in topography between individual subjects’ true connectivity topography and group-average ICA maps used by the ICA dual regression^64,65^. This residual functional misalignment can mean that between-subject spatial variability appears as variability in network connectivity; the extent of this problem of misinterpretation may vary across different analysis methods (e.g., group-ICA with dual-regression vs hard parcellation). It would be interesting to quantify the effects of spatial misalignment on both parcellation-based and whole-brain ICA-based fMRI traits in the large-scale UKB dataset. Finally, our main analyses were based on parcellation-based full correlations. Although the FMRIB’s ICA-based X-noiseifier (FIX) has been applied to the UKB dataset to remove scanner artifacts and motion effects, full correlation measures can be more sensitive to the remaining global artifacts and noises than partial correlations^66,67^. It is possible to further remove global artifacts by measuring the partial functional connectivity between paired brain regions after removing the dependency of other brain regions^68^. Future studies need to explore parcellation-based partial correlation traits for a large number of parcels (such as the 360 regions in the Glasser360 atlas) with a limited number of time points in the UKB study.

## METHODS

Methods are available in the ***Methods*** section.

*Note: One supplementary information pdf file, one supplementary figure pdf file, and one supplementary table zip file are available*.

## Data Availability

Our results and summary-level data can be downloaded and browsed at http://165.227.92.206/. The individual-level UK Biobank data can be obtained from https://www.ukbiobank.ac.uk/.

## ACKNOWLEDGEMENTS

This research was partially supported by U.S. NIH grants MH086633 (H.Z.) and MH116527 (TF.L.). We thank the individuals represented in the UKB and HCP studies for their participation and the research teams for their work in collecting, processing and disseminating these datasets for analysis. We would like to thank the University of North Carolina at Chapel Hill and Purdue University and their Research Computing groups for providing computational resources and support that have contributed to these research results. This research has been conducted using the UK Biobank resource (application number 22783), subject to a data transfer agreement.

## AUTHOR CONTRIBUTIONS

B.Z., H.Z., and S.M.S designed the study. B.Z., T.L., Z.F., D.X., X.W., and M.G. processed and analyzed the data. Y.L. and B.Z. designed the website and developed online resources. B.Z. wrote the manuscript with feedback from all authors.

## CORRESPINDENCE AND REQUESTS FOR MATERIALS

should be addressed to H.Z.

## COMPETETING FINANCIAL INTERESTS

The authors declare no competing financial interests.

## METHODS

### Brain imaging data

We generated functional connectivity measures from the raw resting and task fMRI data downloaded from the UKB data category 111 and 106, respectively. Details of image acquisition and preprocessing procedures were summarized in the **Supplementary Note**. We mapped the preprocessed images onto the Glasser360 atlas^17^, which projected the fMRI data onto a brain parcellation with 360 areas, resulting in a 360 × 360 functional full correlation matrix for each subject (full correlation). The Glasser360 atlas was originally a surface-based parcellation^69^, and has been converted into a volumetric atlas that is compatible with UKB data (**Supplementary Note**). The 360 brain functional areas were grouped into 12 functional networks^19^, including the primary visual, secondary visual, auditory, somatomotor, cingulo-opercular, default mode, dorsal attention, frontoparietal, language, posterior multimodal, ventral multimodal, and orbito-affective (**Table S1**). The 64,620 (360 × 359/2) functional connectivity measures were studied in our main analyses. These high-resolution fMRI traits provided fine details on cerebral cortex functional organization and allowed us to compare the resting and task-evoked functional architectures. To investigate the potential cross-parcellation variability, we also projected the fMRI data onto the Schaefer200 atlas^20^ and obtained the 200 × 200 functional connectivity matrices (full correlation, **Table S2**). The resting and task fMRI data from the HCP study were also used in our analysis (**Supplementary Note**). In addition to functional connectivity measures, we generated amplitude measures for the brain functional areas in the Glasser360 atlas, which quantified the brain functional activity^23,24^. The UKB study has obtained ethics approval from the North West Multi-Centre Research Ethics Committee (MREC, approval number: 11/NW/0382), and obtained written informed consent from all participants prior to the study. All experimental procedures in the HCP study were approved by the institutional review boards at Washington University (approval number: 201204036).

### Age effects and sex differences analysis

Between 2006 and 2010, approximately half a million participants aged 40 to 69 were recruited for the UKB study. The UKB imaging study is an ongoing project to re-invite 100,000 UKB participants to collect multi-modal brain and body imaging data^70^. We used the UKB phases 1 to 4 data (released up through early 2021, *n* = 40,880 for resting fMRI and 34,671 for task fMRI) in our analysis. The age (at imaging) range of subjects was 44 to 82 (mean age = 64.15, standard error = 7.74) and the proportion of female was 51.6%. In the age and sex analysis, we fitted the following model for each fMRI trait: *y* = *xβ*_1_ + *zβ*_2_ + *xzα* + *wη* + *ϵ*, where *y* is the standardized fMRI trait, *x* is the standardized age, *z* is the sex factor (0 for female and 1 for male), *w* is the set of adjusted covariates, *β*_1_ is the main effect of *x* on *y, β*_2_ is the main effect of *z* on *y, α* is the effect of age-sex interaction term *xz* on *y, η* represents effects of covariates, and *∈* is random error variable. We adjusted the following covariates: imaging site, head motion, head motion-squared, brain position, brain position-squared, volumetric scaling, height, weight, body mass index, heel bone mineral density, the top 10 genetic principal components. For each continuous trait or covariate variable, we removed values greater than five times the median absolute deviation from the median. These removed values will be treated as missing entries in the dataset. We performed the analysis in a discovery-validation design and only reported the results that were significant in both discovery and validation datasets (at different significance levels). Specifically, we used the UKB white British subjects in phases 1 to 3 data (*n* = 33,795 for resting and 28, 907 for task) as our discovery sample. The assignment of ancestry in UKB was based on self-reported ethnicity and has been verified in Bycroft, et al. ^16^. The UKB non-British subjects in phases 1 to 3 data and the individuals in newly released UKB phase 4 data (*n* = 5,961 for resting and 4,884 for task, removed relatives of the discovery sample) were treated as the validation sample. We reported *P* values from the two-sided t test and focused on the results that were significant at Bonferroni significance level (7.73 × 10^−7^, 0.05/64,620 for the Glasser360 atlas; and 2.51 × 10^−6^, 0.05/19,900 for the Schaefer200 atlas) in the discovery dataset and were also significant at nominal significance level (0.05) in the validation dataset.

### Trait-fMRI association analysis

For each fMRI trait, we performed linear regression with 647 phenotypes, which were selected to reflect a variety of traits and diseases across different domains (**Table S3**). Specifically, there were 24 mental health traits (Category 100060), 10 cognitive traits (Category 100026), 12 physical activity traits (Category 100054), 6 electronic device use traits (Category 100053), 8 sun exposure traits (Category 100055), 3 sexual factor traits (Category 100056), 3 social support traits (Category 100061), 12 family history of diseases (Category 100034), 21 diet traits (Category 100052), 9 alcohol drinking traits (Category 100051), 6 smoking traits (Category 100058), 34 blood biochemistry biomarkers (Category 17518), 3 blood pressure traits (Category 100011), 3 spirometry traits (Category 100020), 20 early life factors (Categories 135, 100033, 100034, and 100072), 9 greenspace and coastal proximity (Category 151), 2 hand grip strength (Category 100019), 13 residential air pollution traits (Category 114), 5 residential noise pollution traits (Category 115), 2 body composition traits by impedance (Category 100009), 4 health and medical history traits (Category 100036), 3 female specific factors (Category 100069), 1 education trait (Category 100063), 48 curated disease phenotypes based on Dey, et al. ^71^, and 386 disease diagnosis coded according to International Classification of Diseases (ICD-10, Category 2002). We selected all diseases in Category 2002 that had at least 100 patients in our resting fMRI imaging cohort.

For all traits, we adjusted for the effects of age (at imaging), age-squared, sex, age-sex interaction, age-squared-sex interaction, imaging site, head motion, head motion-squared, brain position, brain position-squared, volumetric scaling, height, weight, body mass index, heel bone mineral density, and the top 10 genetic principal components. Similar to the age and sex analysis, we used the UKB white British subjects in phases 1 to 3 data (*n* = 33,795 for resting and 28, 907 for task) as our discovery sample and validated our results in the hold-out independent validation dataset (*n* = 5,961 for resting and 4,884 for task, removed relatives of the discovery sample). We reported *P* values from the two-sided t test and prioritized on the results that were significant at FDR 5% level in the discovery dataset and were also significant at nominal significance level (0.05) in the validation dataset.

### Prediction models with multiple data types

We built prediction models for fluid intelligence using multi-modality neuroimaging traits, including 64,620 resting fMRI traits, 64,620 task fMRI traits, 215 DTI parameters from dMRI^21^, and 101 regional brain volumes from sMRI^57^. After removing relatives according to Bycroft, et al. ^16^, we randomly partitioned the white British imaging subjects into three independent datasets: training (*n* = 20,270), validation (*n* = 6,764), and testing (*n* = 6,761). The effect sizes of imaging predictors were estimated from the training data (*n* = 20,270). We removed the effects of age, age-squared, sex, age-sex interaction, age-squared-sex interaction, imaging site, head motion, head motion-squared, brain position, brain position-squared, volumetric scaling, height, weight, body mass index, heel bone mineral density, and the top 10 genetic principal components.

We also integrated other data types into our prediction model, including genetic variants and several categories of traits studied in our trait-fMRI association analysis (**Table S4**). For non-neuroimaging traits, the effect sizes were estimated from all UKB white British subjects except for the ones in validation and testing data (after removing relatives). We adjusted for all the covariates listed above for neuroimaging traits, except for the imaging-specific variables including imaging site, head motion, volumetric scaling, and brain position. The genetic effects were estimated by fastGWA^72^ and were aggregated using polygenic risk scores via lassosum^73^. We downloaded imputed genotyping data (Category 100319) and performed the following quality controls^57^: 1) excluded subjects with more than 10% missing genotypes; 2) excluded variants with minor allele frequency less than 0.01; 3) excluded variants with missing genotype rate larger than 10%; 4) excluded variants that failed the Hardy-Weinberg test at 1 × 10^−7^ level; and 5) removed variants with imputation INFO score less than 0.8. All non-genetic predictors (including neuroimaging traits) were modeled using ridge regression via glmnet^74^ (R version 3.6.0). All model parameters were tuned in the validation dataset, and we evaluated the prediction performance on the testing data by calculating the correlation between the predicted values and the observed ones.

## Code availability

We made use of publicly available software and tools. The codes used to generate fMRI traits are publicly available on Zenodo (https://doi.org/10.5281/zenodo.5784010).

## Notes

### Competing Interest Statement

The authors have declared no competing interest.

### Author Declarations

The individual-level UK Biobank data can be obtained from https://www.ukbiobank.ac.uk/.

### Summary of Updates

The previous issue has been successfully solved, and we would like to upload the new version.

