## Supplementary material for "An atlas of trait associations with resting-state and task-evoked human brain functional architectures in the UK Biobank": supp_figures

**This PDF file includes:**

Supplementary Figures Figs. S1-S51.

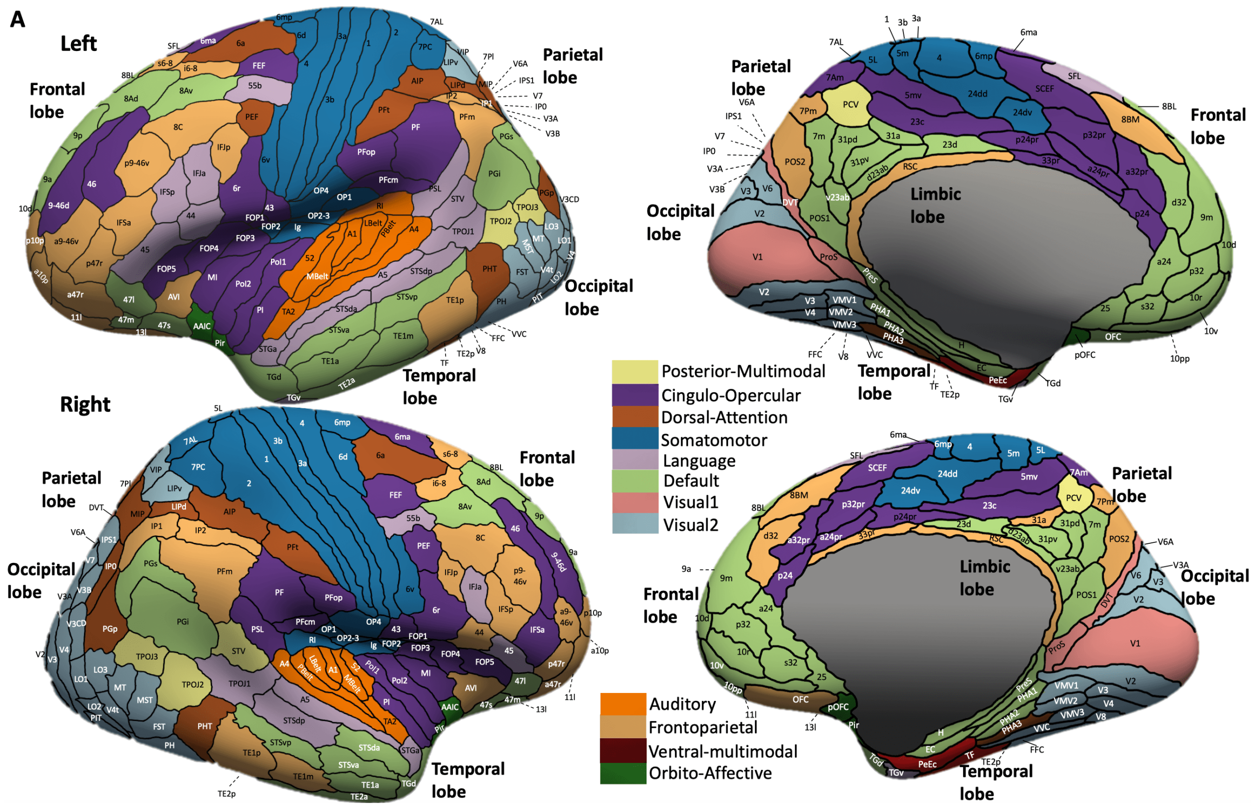


**Fig. S1 Location of the 360 functional areas defined in the Glasser360 atlas.**

See Table S1 for more information of the areas. Visual1, primary visual network; Visual2, secondary visual network; Default, default mode network.

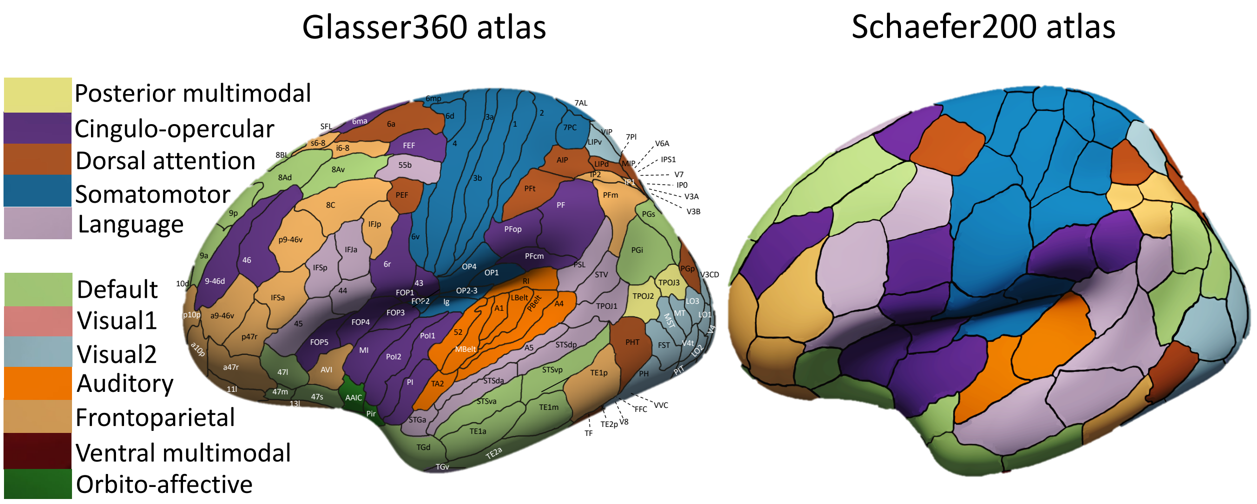


**Fig. S2 Comparison of the functional areas defined in the Glasser 360 and Schaefer200 atlases.**

See Table S2 for more information of the areas defined in the Schaefer200 atlas. Visual1, primary visual network; Visual2, secondary visual network; Default, default mode network.

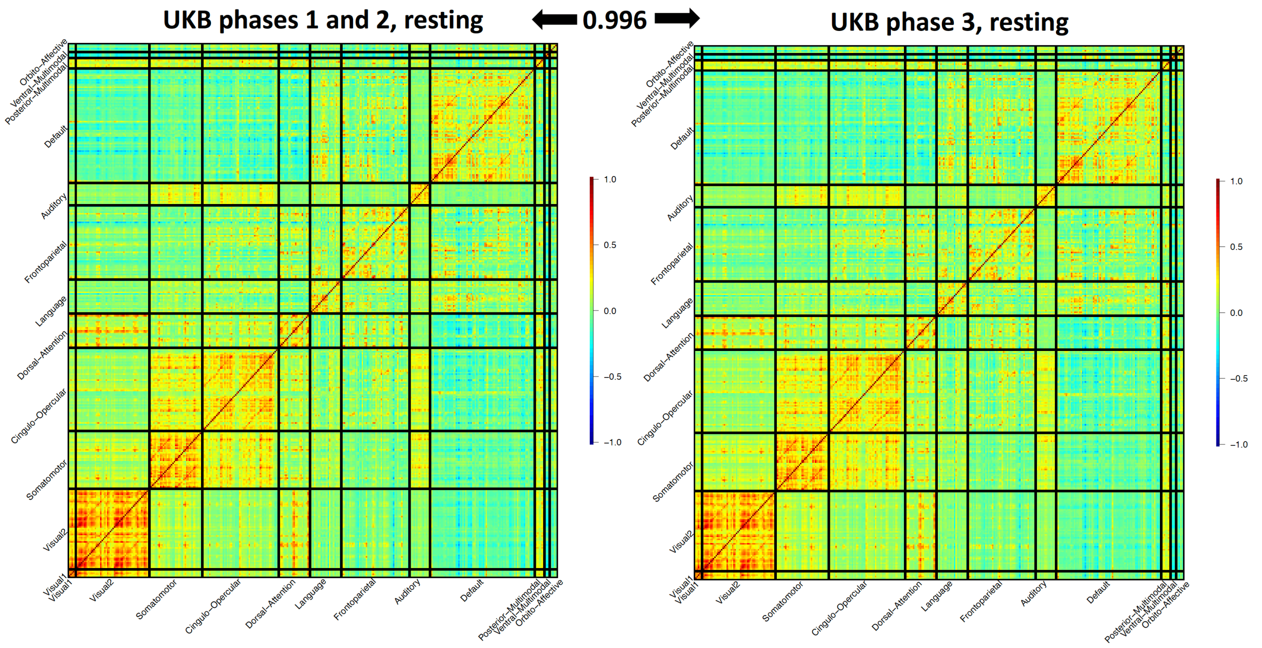


**Fig. S3 Group mean maps of resting-state fMRI in UKB phases 1 and 2 dataset (left) and UKB phase 3 dataset (right).**

The sample size in UKB phases 1 and 2 dataset was 17,374 subjects, and the sample size in UKB phase 3 dataset was 16,852 (removing the relatives of subjects in early released dataset). We calculated the group average of functional connectivity for all the 64,620 (360 × 359 / 2) functional connectivity measures across all subjects within each of the two groups. The correlation of group means across all the 64620 functional connectivity measures was 0.996. Visual1, primary visual network; Visual2, secondary visual network; Default, default mode network.

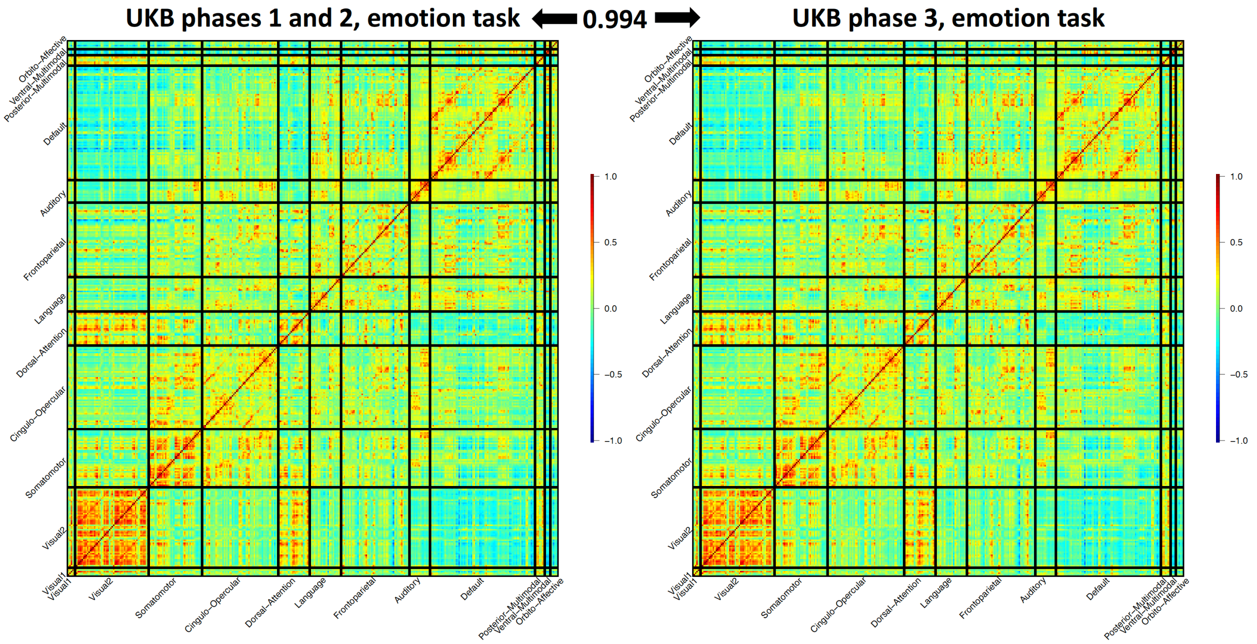


**Fig. S4 Group mean maps of task-evoked fMRI in UKB phases 1 and 2 dataset (left) and UKB phase 3 dataset (right).**

The sample size in UKB phases 1 and 2 dataset was 15,891 subjects, and the sample size in UKB phase 3 dataset was 13,232 (removing the relatives of subjects in early released dataset). We calculated the group average of functional connectivity for all the 64,620 (360 × 359 / 2) functional connectivity measures across all subjects within each of the two groups. The correlation of group means across all the 64,620 functional connectivity measures was 0.994. Visual1, primary visual network; Visual2, secondary visual network; Default, default mode network.

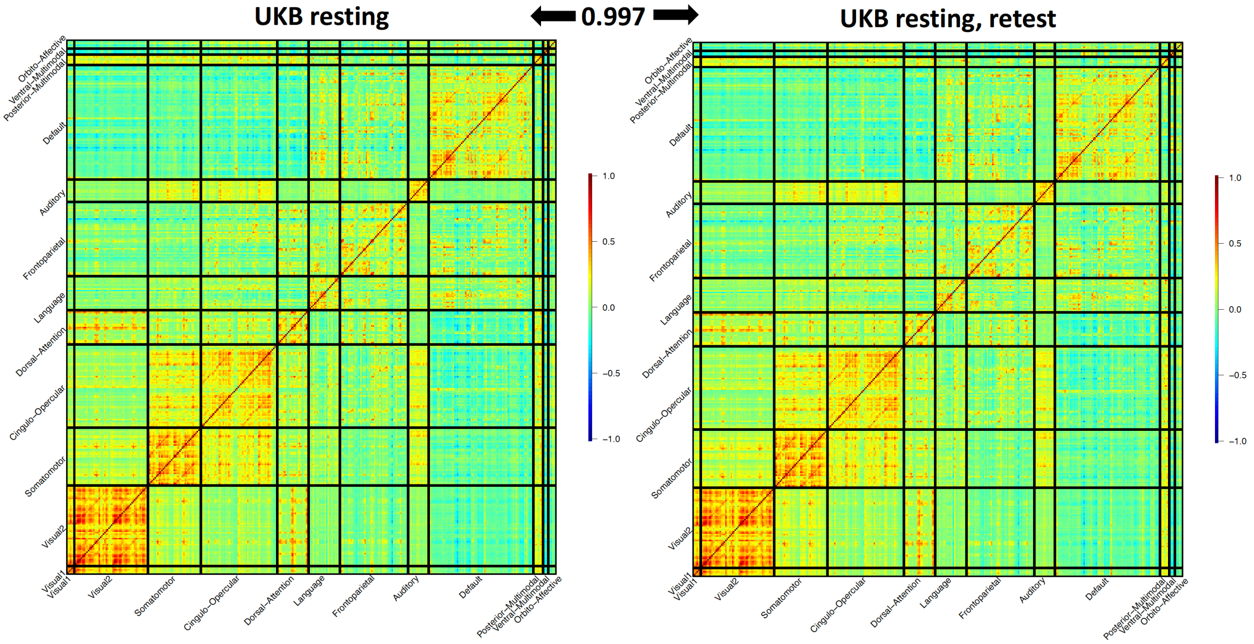


**Fig. S5 Group mean maps of resting-state fMRI in the UKB original imaging visit (left) and the UKB repeat imaging visit (right).**

The sample size of the UKB original imaging visit (UKB phases 1 to 3) dataset was 37,794 subjects, and the sample size of the UKB repeat imaging visit dataset was 2,771. We calculated the group average of functional connectivity for each functional connectivity across all subjects within each of the two datasets. The correlation of group means across all the 64620 functional connectivity measures is 0.997. Visual1, primary visual network; Visual2, secondary visual network; Default, default mode network.

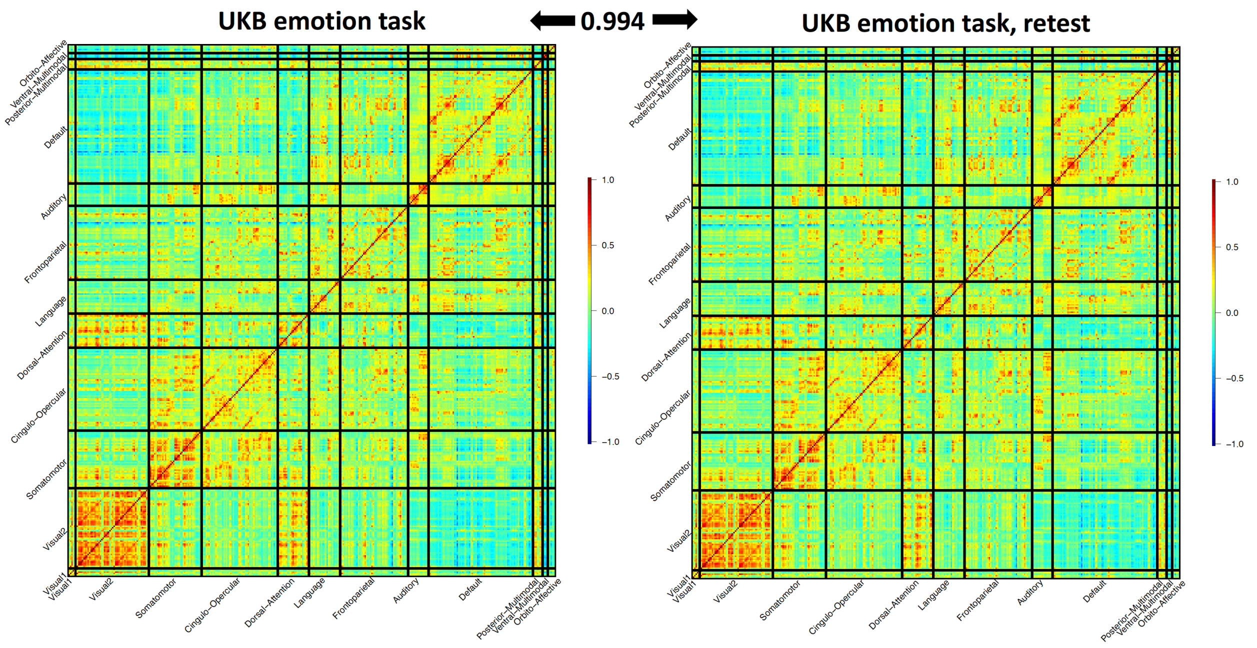


**Fig. S6 Group mean maps of task-evoked fMRI in the UKB original imaging visit (left) and the UKB repeat imaging visit (right).**

The sample size of the UKB original imaging visit (UKB phases 1 to 3) dataset was 32,144 subjects, and the sample size of the UKB repeat imaging visit dataset was 2,014. We calculated the group average of functional connectivity for each functional connectivity across all subjects within each of the two datasets. The correlation of group means across all the 64620 functional connectivity measures was 0.994. Visual1, primary visual network; Visual2, secondary visual network; Default, default mode network.

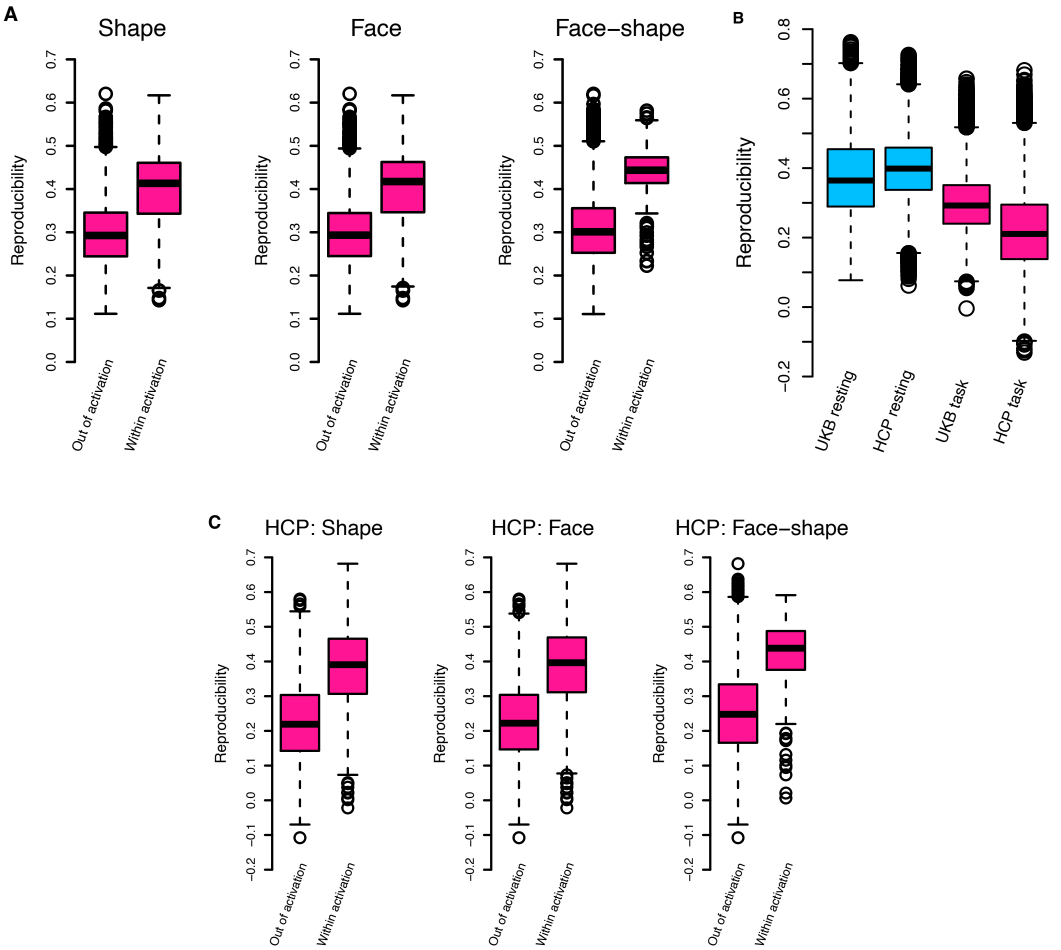


**Fig. S7 Reproducibility of fMRI connectivity in UK Biobank and HCP.**

**A:** Comparison of the reproductivity between connectivity within the activated functional areas (within activation) in UKB task-evoked fMRI and connectivity within nonactivated areas (out of activation). Shape, shape activation contrast, Face, face activation contrast, Face-shape, face-shape activation contrast. See https://biobank.ctsu.ox.ac.uk/crystal/crystal/docs/brain_mri.pdf for details. **B:** Comparison of the reproductivity between UKB and HCP for the resting-state fMRI (blue) and task-evoked fMRI (red). **C:** Comparison of the reproductivity between connectivity within the activated functional areas of HCP task-evoked fMRI and connectivity within nonactivated areas.

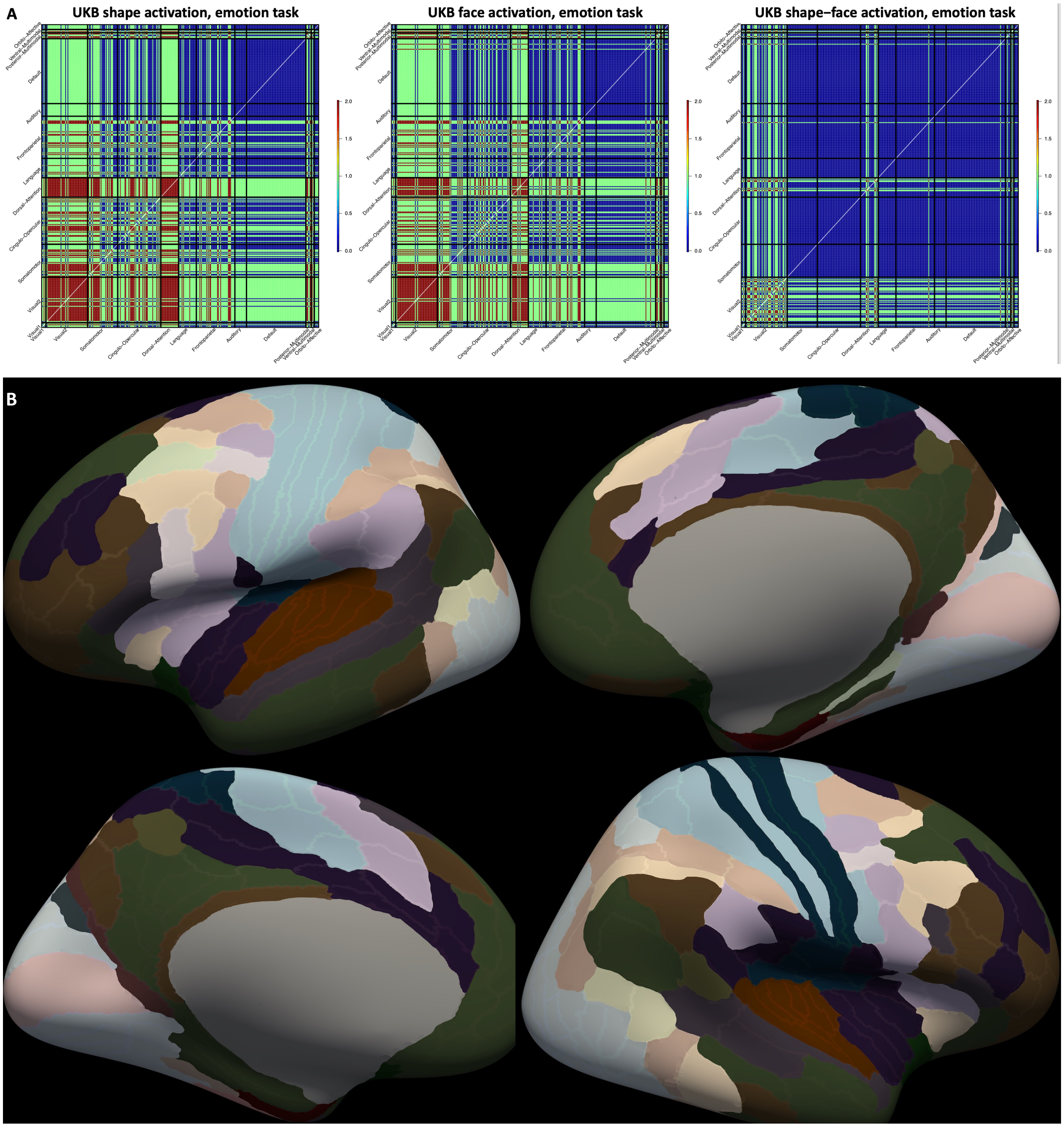


**Fig. S8 Illustration of the activated functional areas and networks in the emotion processing task.**

**A.** We illustrated the functional connectivity that were between the activated functional areas (in red), between the nonactivated areas (in blue), and between one activated area and one nonactivated area (in green). **B.** The activated functional areas (in the shape activation) are highlighted.

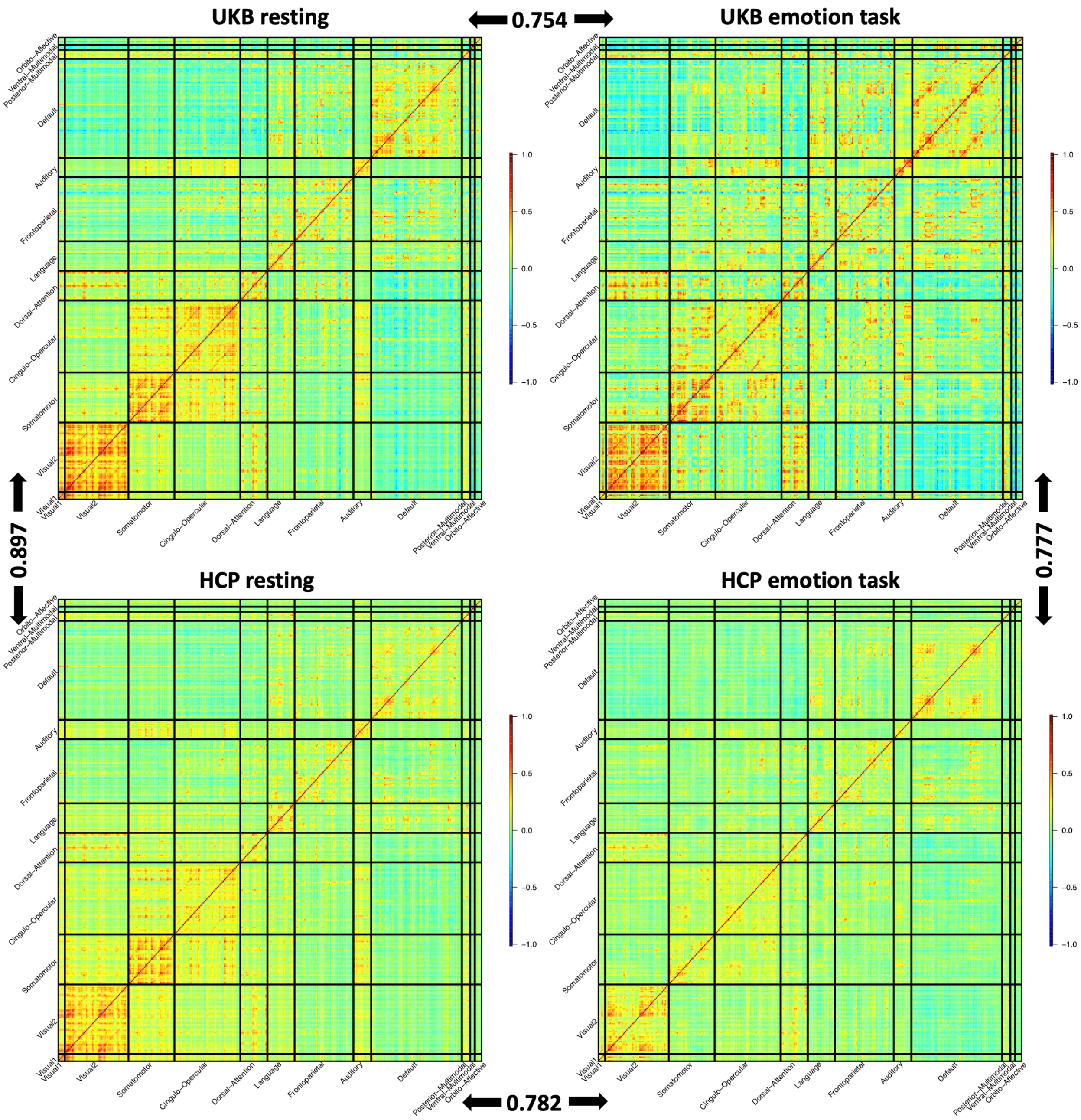


**Fig. S9 Comparation of the group mean spatial patterns in UKB and HCP studies.**

The correlation between UKB and HCP group mean maps was 0.897 for resting-state fMRI and 0.777 for task-evoked fMRI.

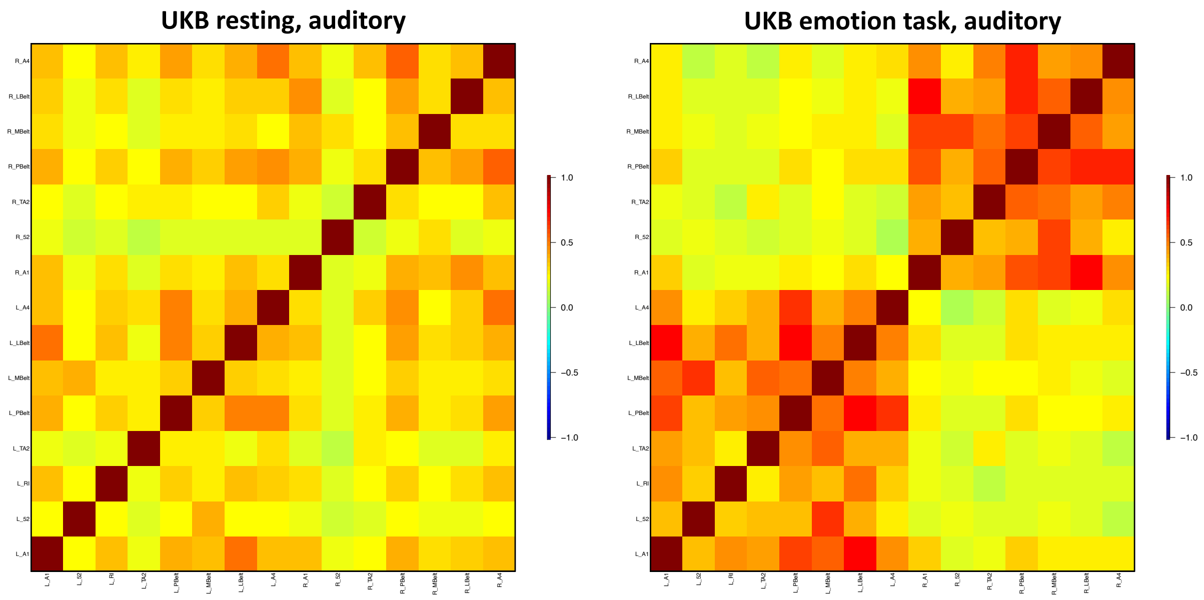


**Fig. S10 Comparation of the auditory network spatial patterns in resting and task fMRI.**

The sample size was 37,794 subjects for resting fMRI and 32,144 subjects for task fMRI. We calculated the group average for each functional connectivity across all subjects.

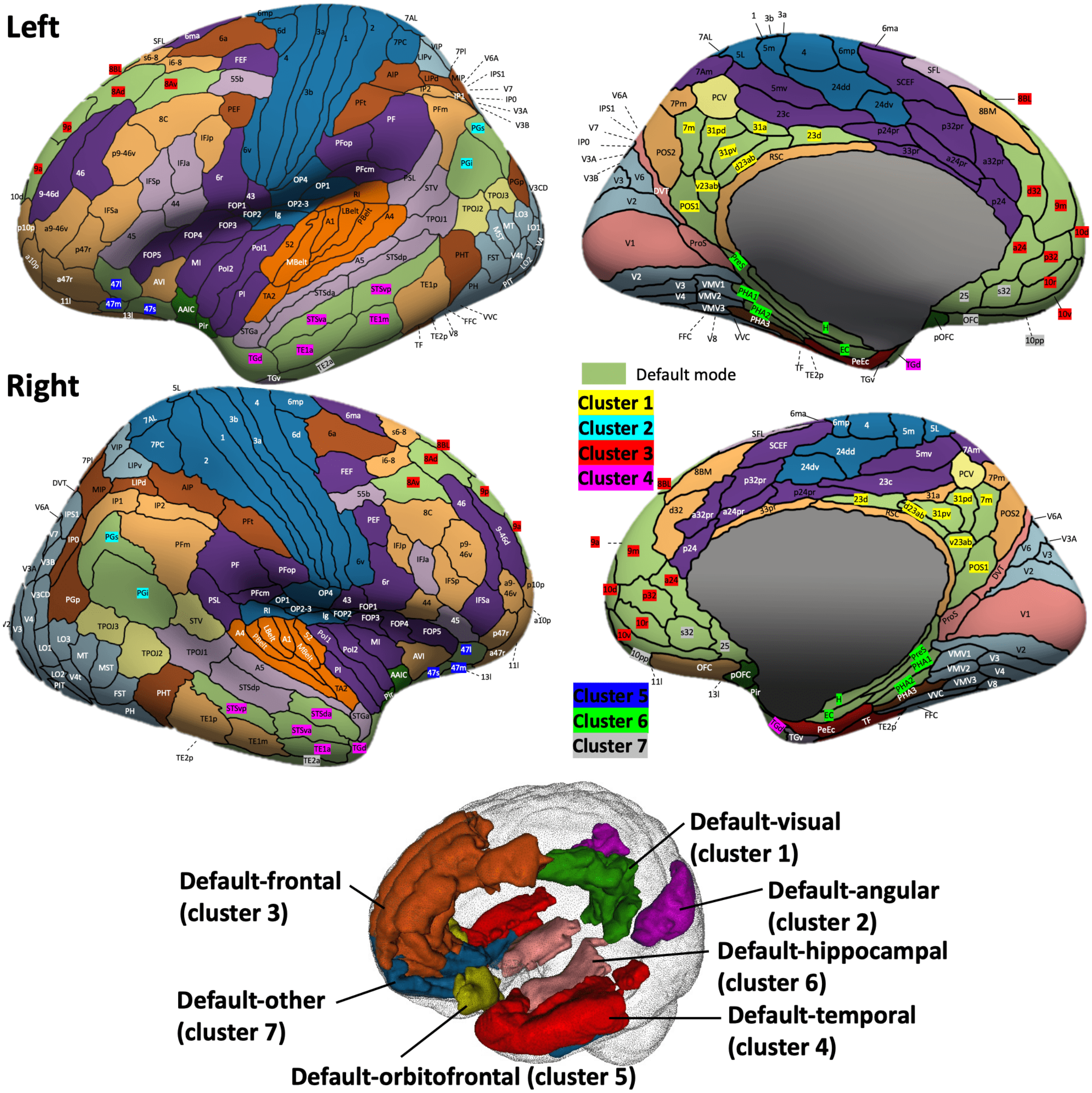


**Fig. S11 Location of the 7 clusters in the default mode network.**

We divided the functional areas in the default mode network into 7 clusters, mainly based on their locations in the brain. The cluster 1 was mainly in the visual cortex (precuneus, calcarine, and cingulate), the cluster 2 was mainly in the angular, the cluster 3 was mainly in the frontal, the cluster 4 was mainly in the temporal, the cluster 5 was mainly in the orbitofrontal, the cluster 6 was mainly in the hippocampus, and the cluster 7 included other areas.

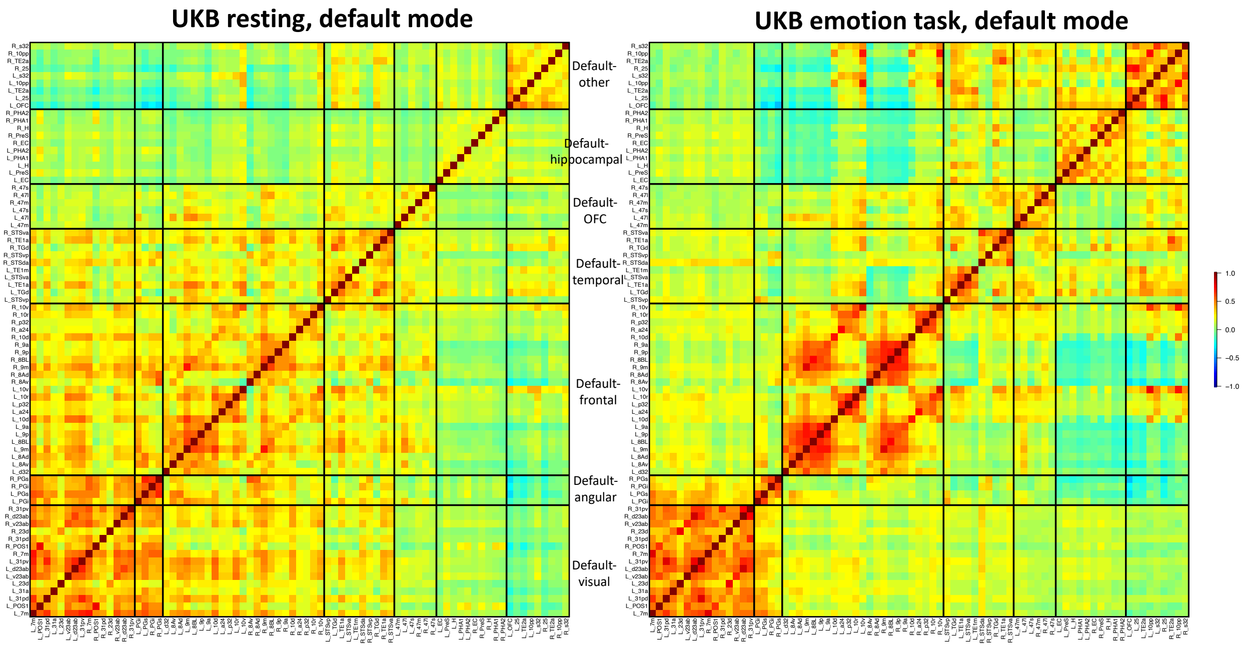


**Fig. S12 Comparation of the default mode network spatial patterns in resting and task fMRI.**

The sample size was 37,794 subjects for resting fMRI and 32,144 subjects for task fMRI. We calculated the group average for each functional connectivity across all subjects. We grouped all functional areas in the default mode network into seven clusters. These areas were mainly organized by their physical locations (Fig. S11). OFC, orbitofrontal complex.

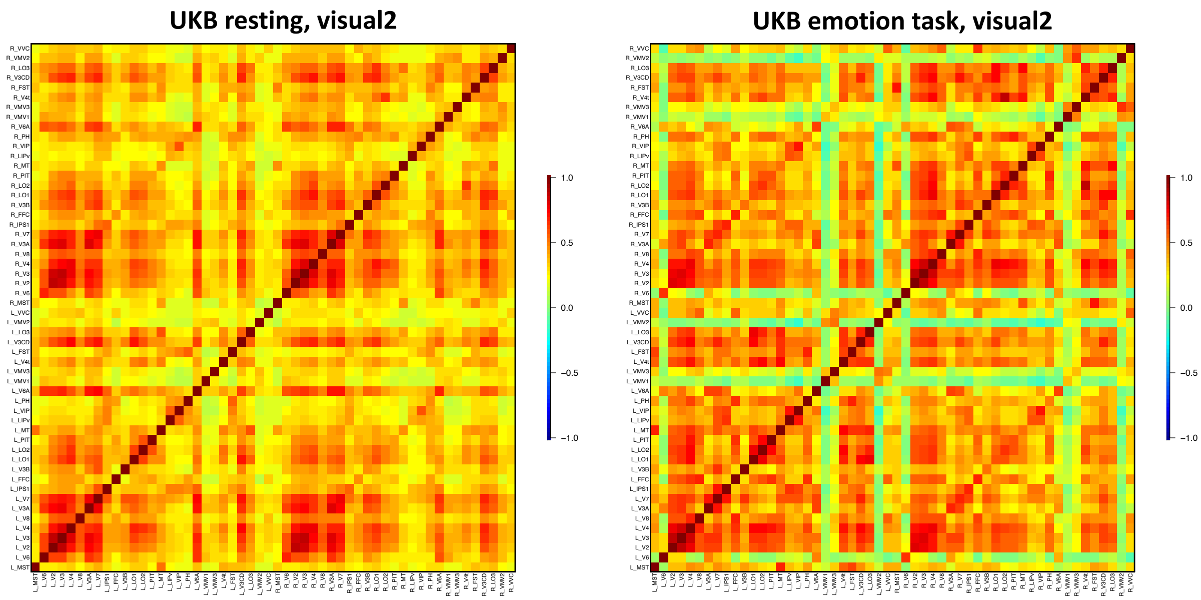


**Fig. S13 Comparation of the secondary visual network spatial patterns in resting and task fMRI.**

The sample size was 37,794 subjects for resting fMRI and 32,144 subjects for task fMRI. We calculated the group average for each functional connectivity across all subjects.

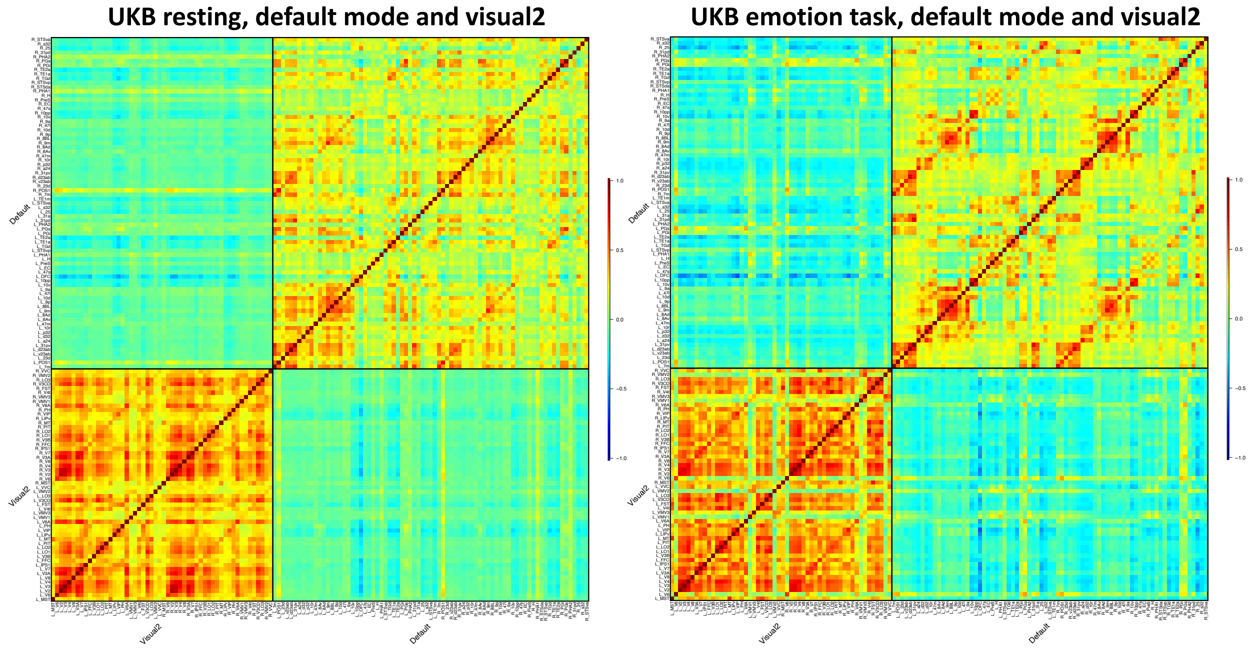


**Fig. S14 Comparation of the secondary visual network spatial patterns in resting and task fMRI.**

The sample size was 37,794 subjects for resting fMRI and 32,144 subjects for task fMRI. We calculated the group average for each functional connectivity across all subjects.

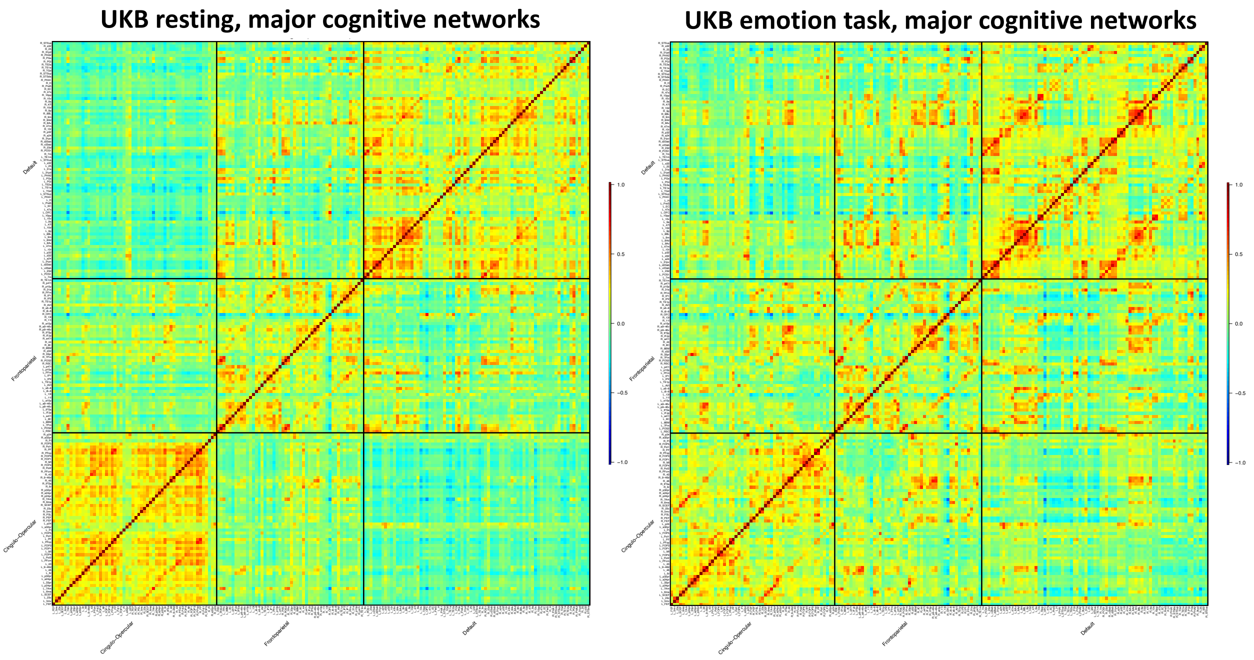


**Fig. S15 Comparation of spatial patterns of major cognitive networks in resting and task fMRI.**

The sample size was 37,794 subjects for resting fMRI and 32,144 subjects for task fMRI. We calculated the group average for each functional connectivity across all subjects. The three major cognitive networks included the default mode, cingulo-opercular, and frontoparietal networks.

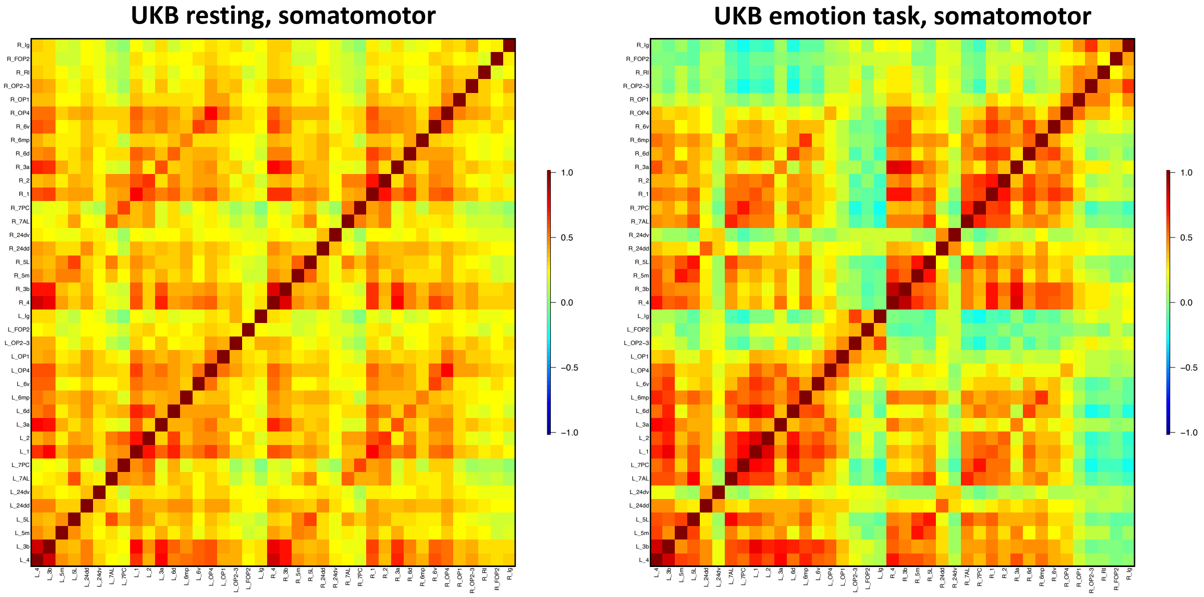


**Fig. S16 Comparation of the somatomotor network spatial patterns in resting and task fMRI.**

The sample size was 37,794 subjects for resting fMRI and 32,144 subjects for task fMRI. We calculated the group average for each functional connectivity across all subjects.

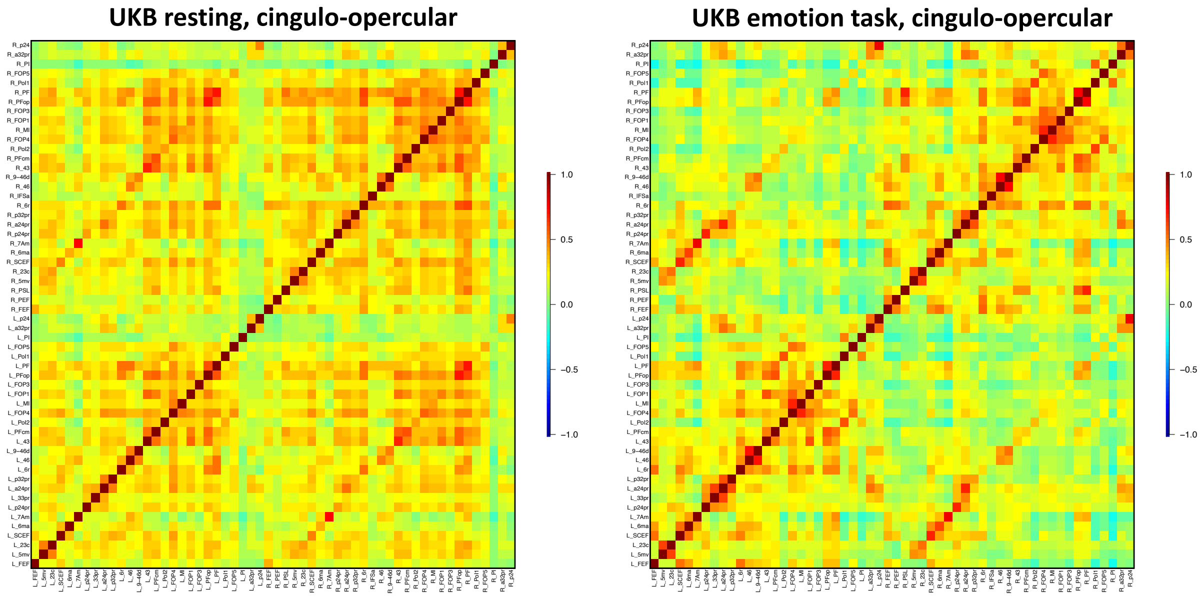


**Fig. S17 Comparation of the cingulo-opercular network spatial patterns in resting and task fMRI.**

The sample size was 37,794 subjects for resting fMRI and 32,144 subjects for task fMRI. We calculated the group average for each functional connectivity across all subjects.

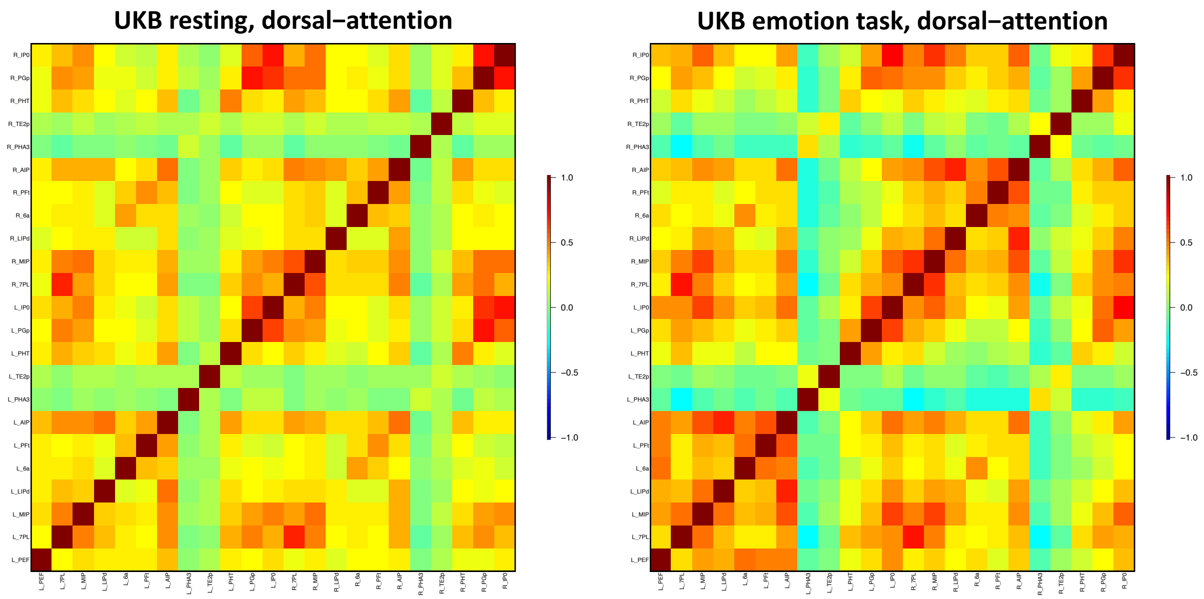


**Fig. S18 Comparation of the dorsal attention network spatial patterns in resting and task fMRI.**

The sample size was 37,794 subjects for resting fMRI and 32,144 subjects for task fMRI. We calculated the group average for each functional connectivity across all subjects.

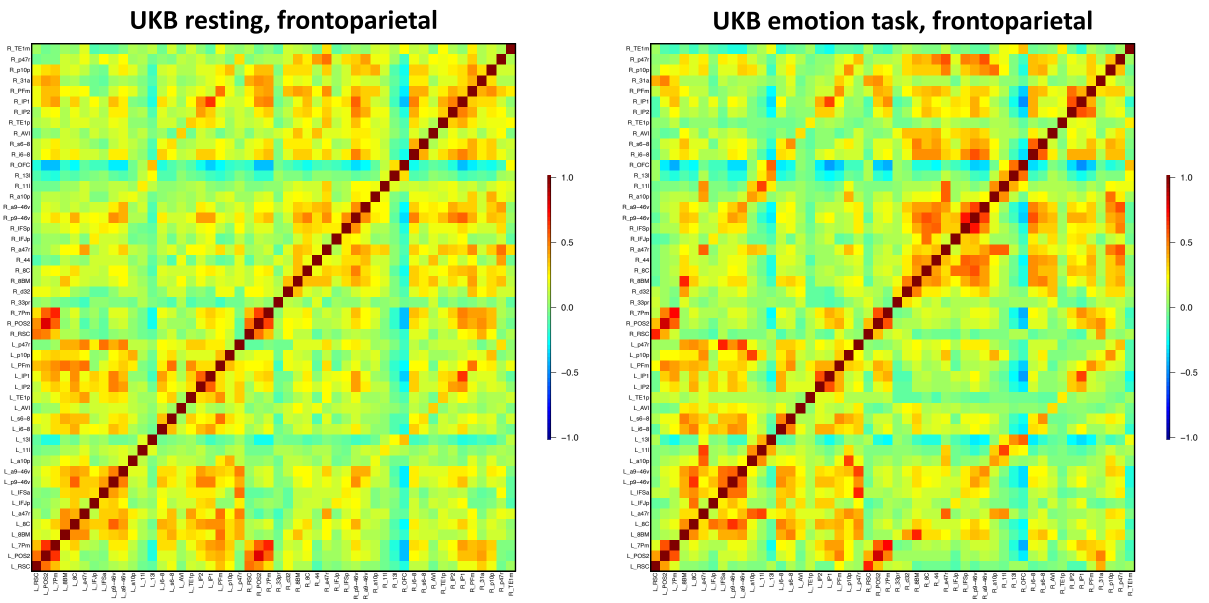


**Fig. S19 Comparation of the frontoparietal network spatial patterns in resting and task fMRI.**

The sample size was 37,794 subjects for resting fMRI and 32,144 subjects for task fMRI. We calculated the group average for each functional connectivity across all subjects.

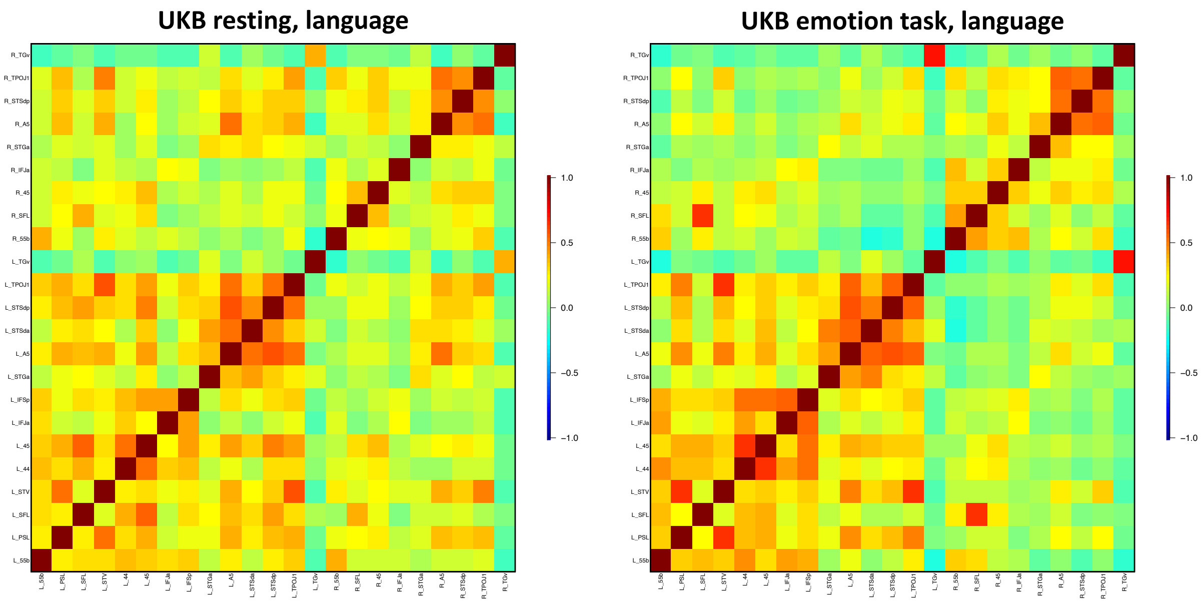


**Fig. S20 Comparation of the language network spatial patterns in resting and task fMRI.**

The sample size was 37,794 subjects for resting fMRI and 32,144 subjects for task fMRI. We calculated the group average for each functional connectivity across all subjects.

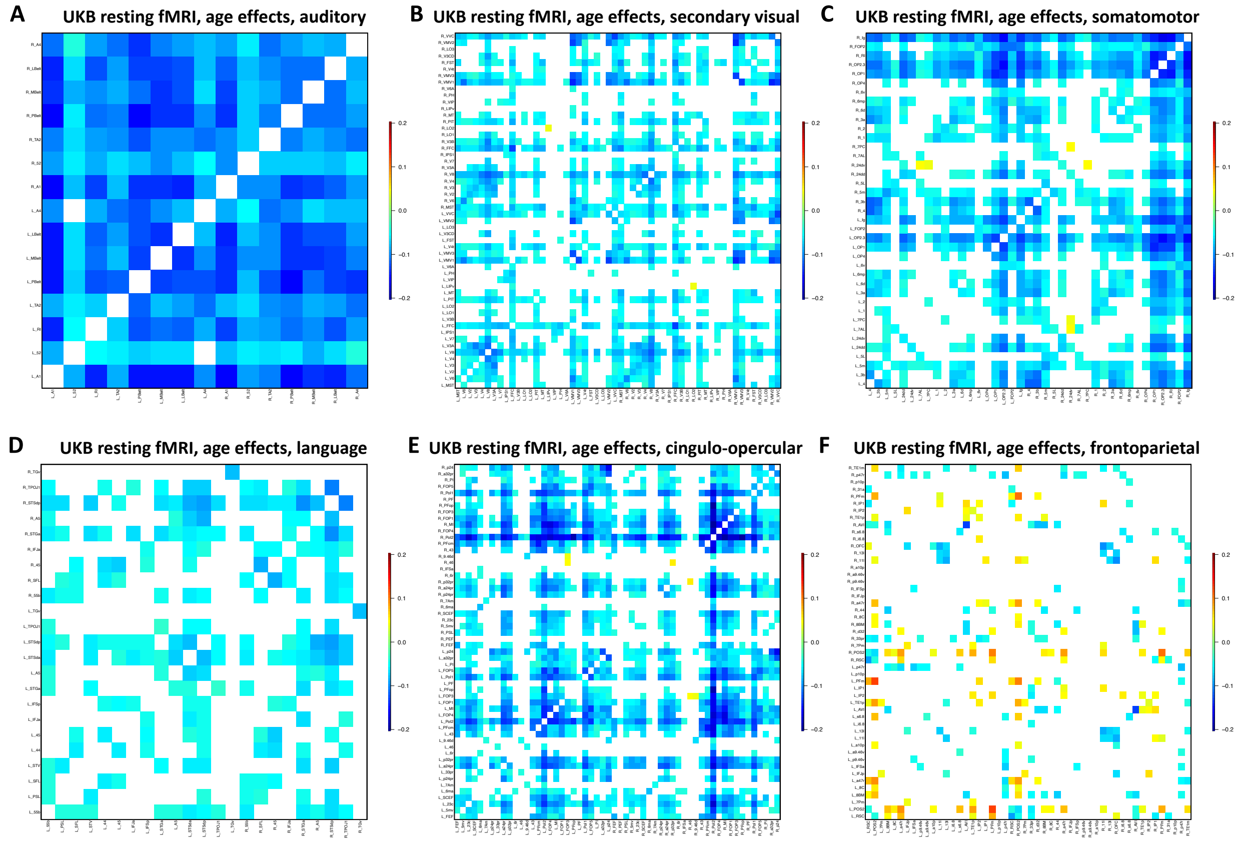


**Fig. S21 Age effect patterns in different functional networks of resting fMRI.**

We illustrated the effects passing the Bonferroni significance level ($7.73 \times 10^{-7}$, 0.05/64,620) in the discovery dataset (*n* = 33,795) and also being significant at the nominal significance level (0.05) in the validation dataset (*n* = 5, 961).

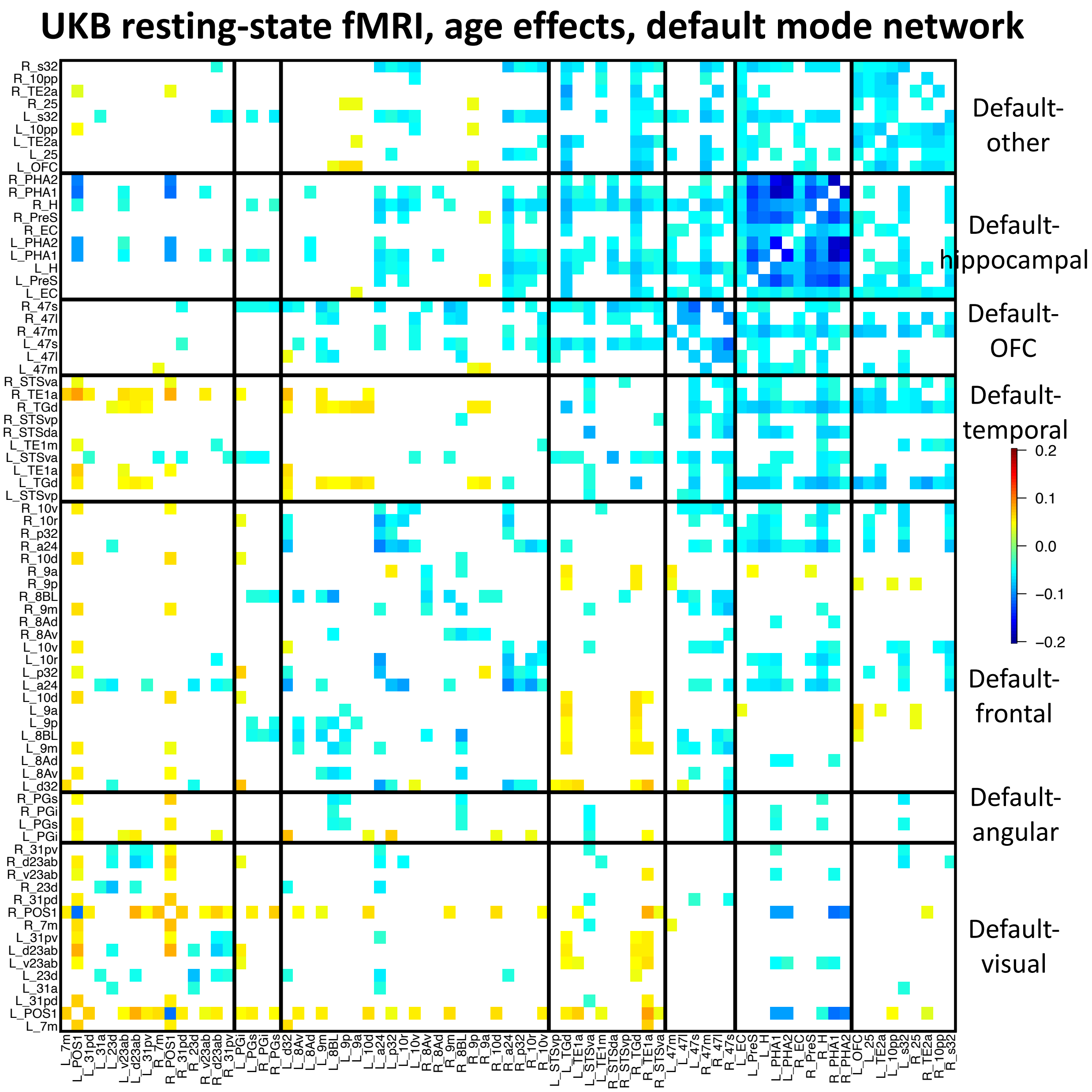


**Fig. S22 Age effect pattern in the default mode network of resting fMRI.**

We illustrated the effects passing the Bonferroni significance level ($7.73 \times 10^{-7}$, 0.05/64,620) in the discovery dataset ($n$ = 33,795) and also being significant at the nominal significance level (0.05) in the validation dataset ($n$ = 5, 961).

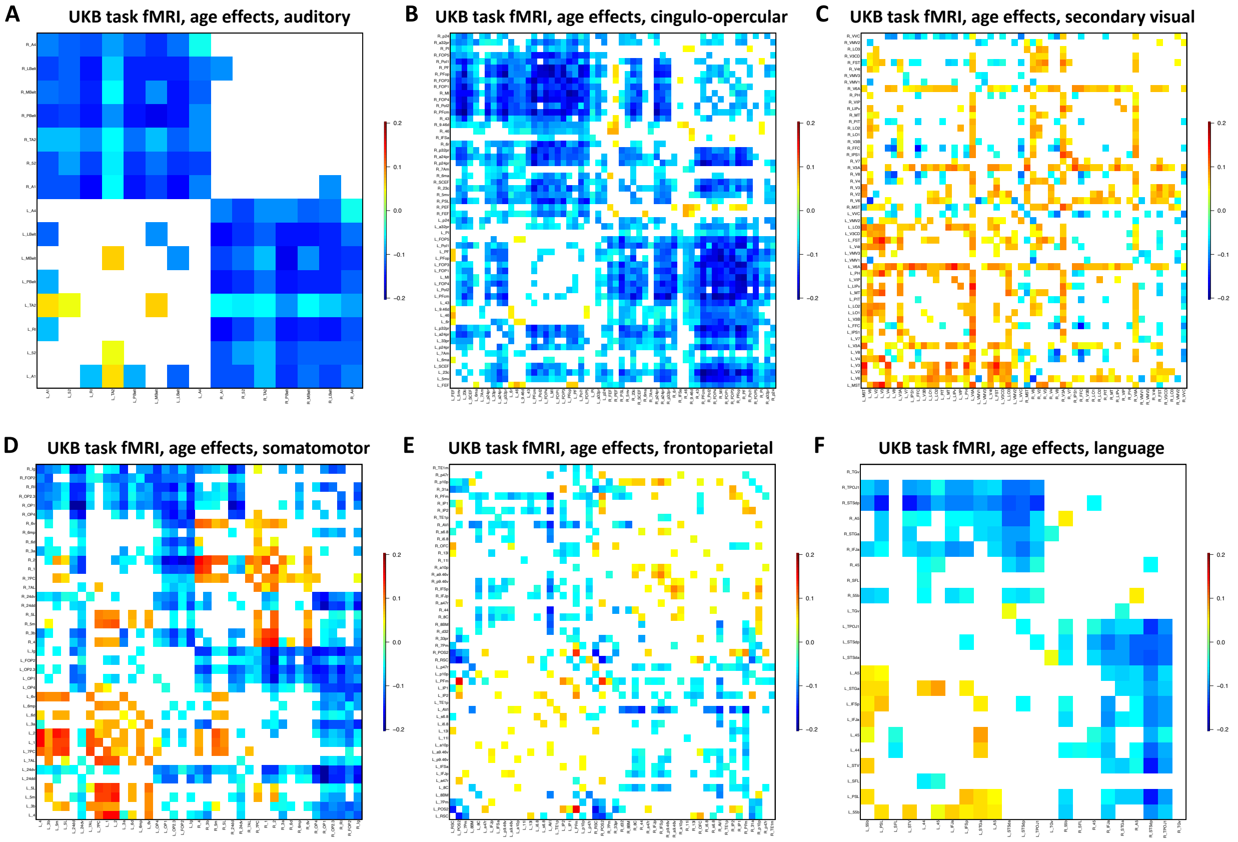


**Fig. S23 Age effect patterns in selected functional networks of task fMRI.**

We illustrated the effects passing the Bonferroni significance level ($7.73 \times 10^{-7}$, 0.05/64,620) in the discovery dataset ($n$ = 28, 907) and also being significant at the nominal significance level (0.05) in the validation dataset ($n$ = 4, 884).

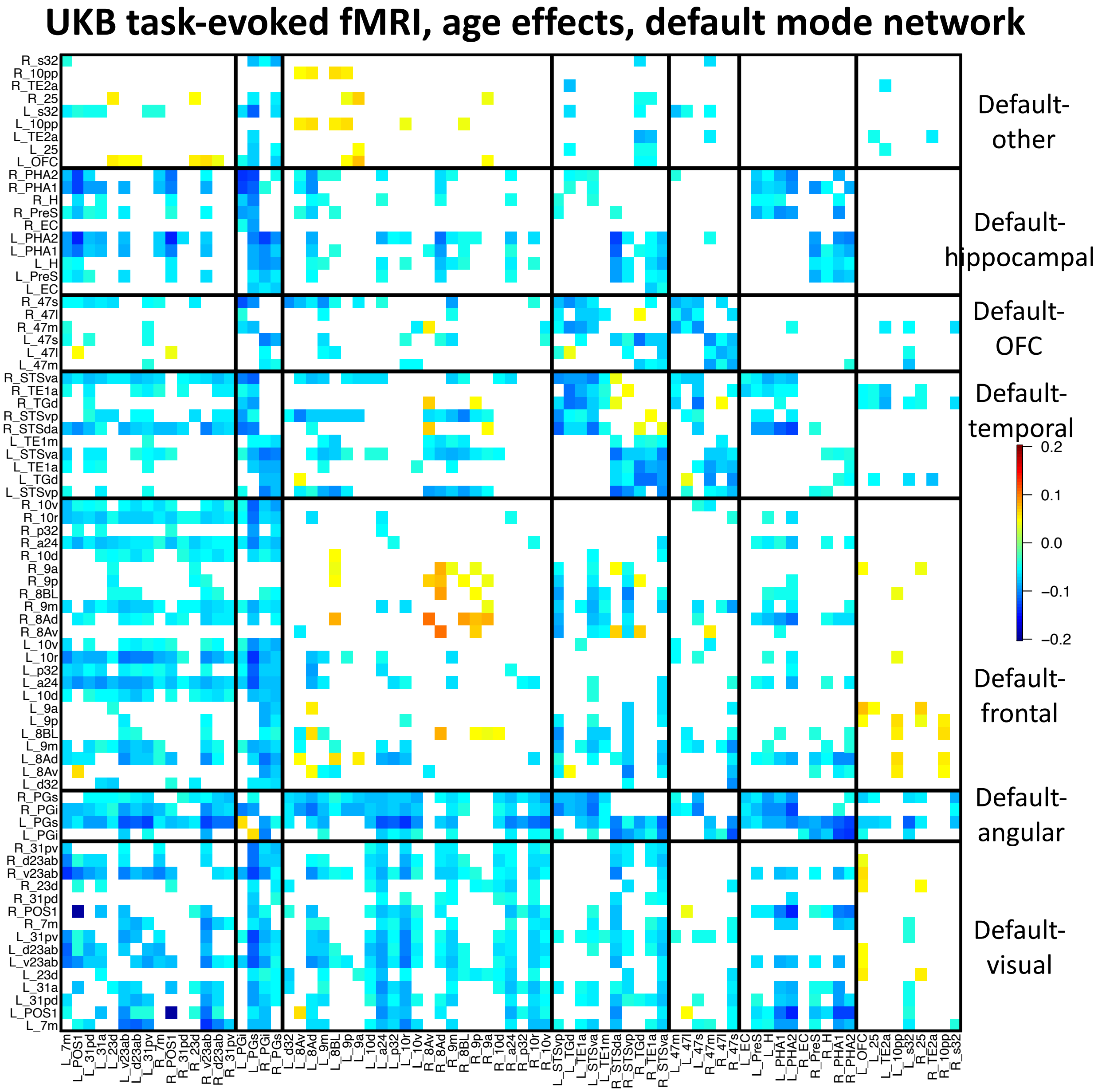


**Fig. S24 Age effect pattern in the default mode network of task fMRI.**

We illustrated the effects passing the Bonferroni significance level ($7.73 \times 10^{-7}$, 0.05/64,620) in the discovery dataset ($n$ = 28, 907) and also being significant at the nominal significance level (0.05) in the validation dataset ($n$ = 4, 884).

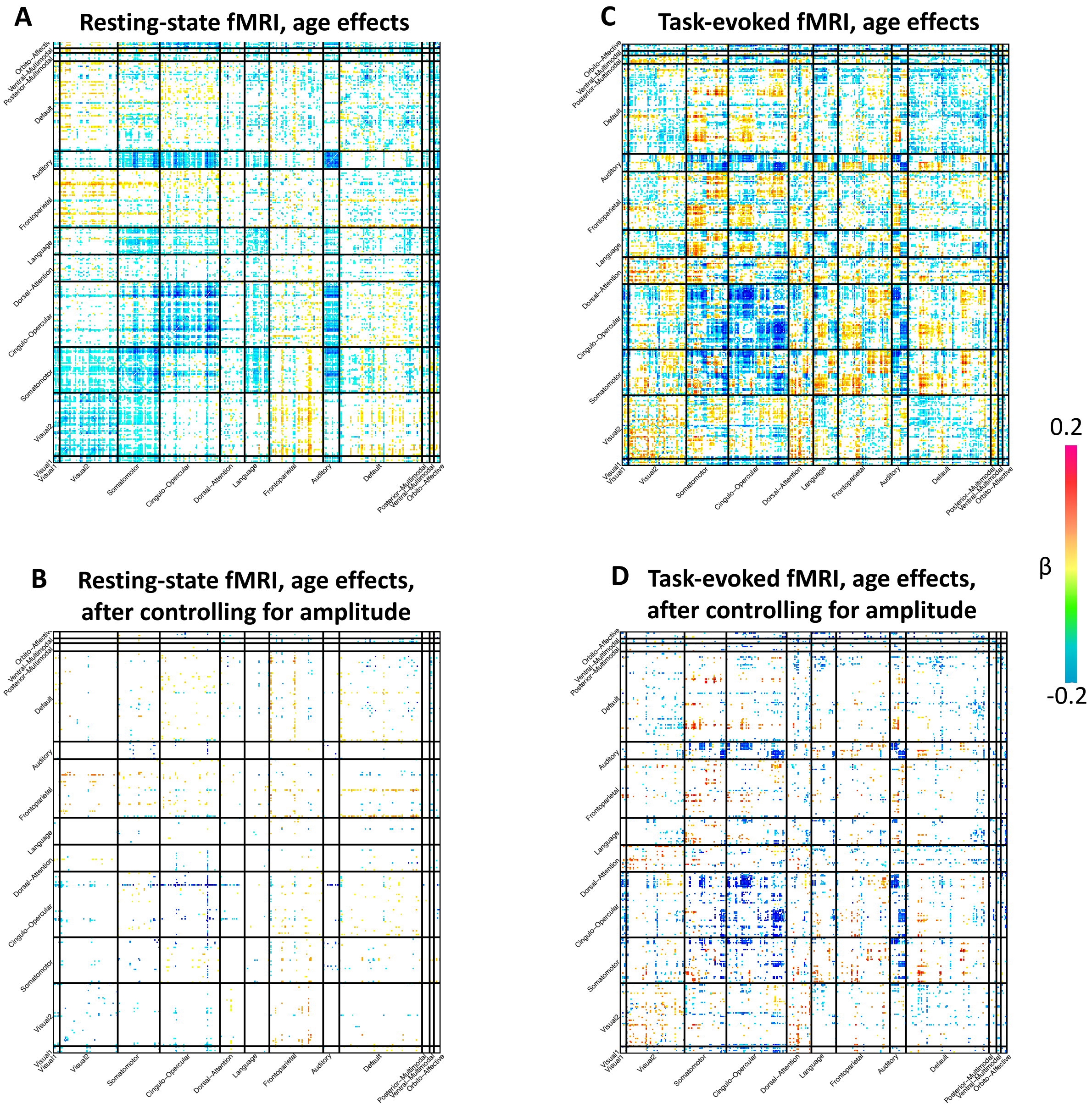


**Fig. S25 Age effect patterns before and after controlling for amplitude traits.**

In A, we illustrated the age effects in resting fMRI passing the Bonferroni significance level (7.73 × $10^{-7}$, 0.05/64,620) in the discovery dataset ($n$ = 33,795) and also being significant at the nominal significance level (0.05) in the validation dataset ($n$ = 5, 961). In B, we illustrated the remaining significant age effects in resting fMRI after additionally controlling for amplitude traits. Similarly, the panel C illustrated the significant age effects in task fMRI and D illustrated the remaining significant age effects in task fMRI after additionally controlling for amplitude traits.

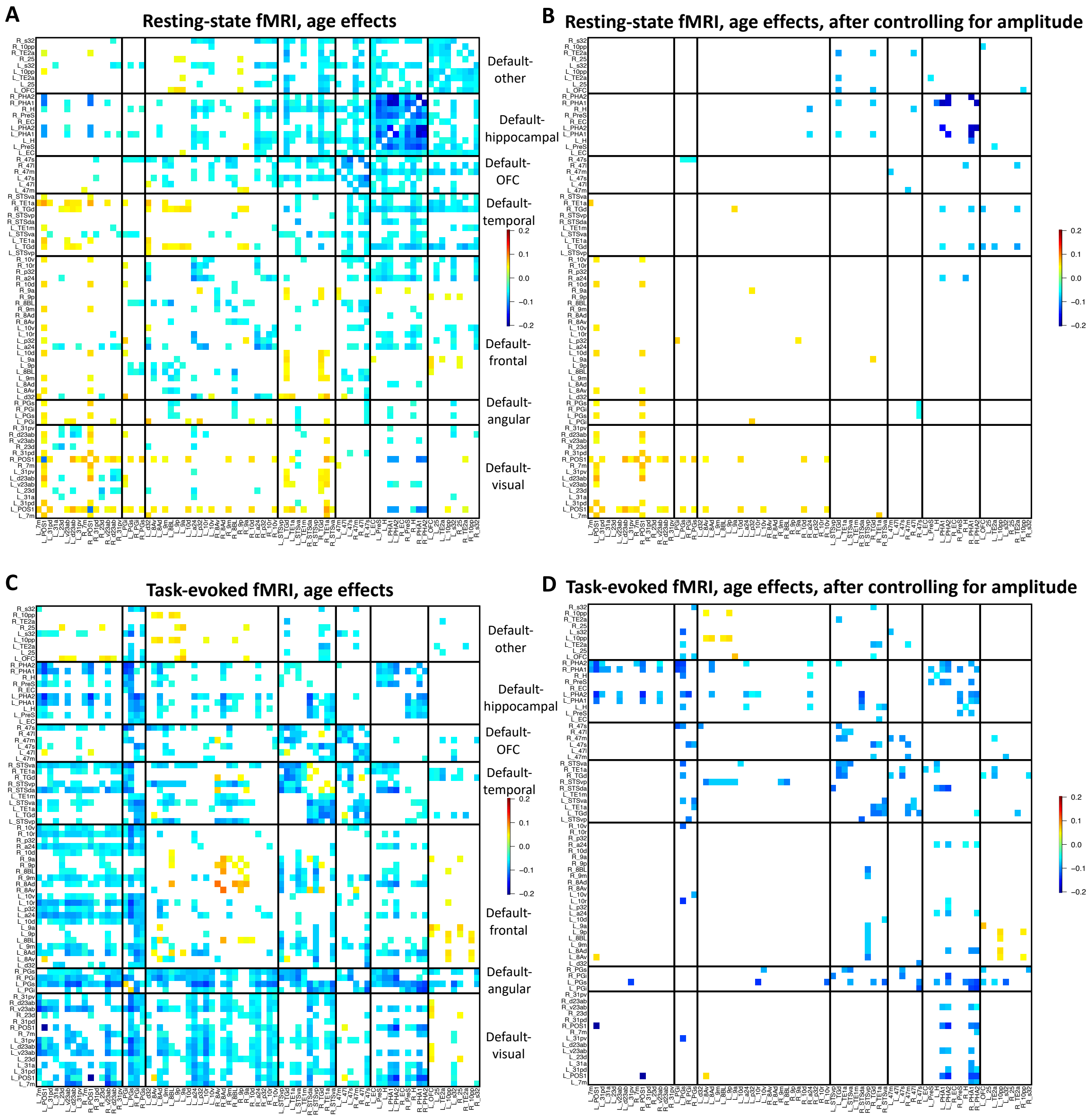


**Fig. S26 Age effect pattern in the default mode network before and after controlling for amplitude traits.**

In A, we illustrated the age effects in the default mode network of resting fMRI passing the Bonferroni significance level ($7.73 \times 10^{-7}$, 0.05/64,620) in the discovery dataset ($n$ = 33,795) and also being significant at the nominal significance level (0.05) in the validation dataset ($n$ = 5, 961). In B, we illustrated the remaining significant age effects in the default mode network of resting fMRI after additionally controlling for amplitude traits. Similarly, the panel C illustrated the significant age effects in the default mode network of task fMRI and D illustrated the remaining significant age effects in the default mode network of task fMRI after additionally controlling for amplitude traits.

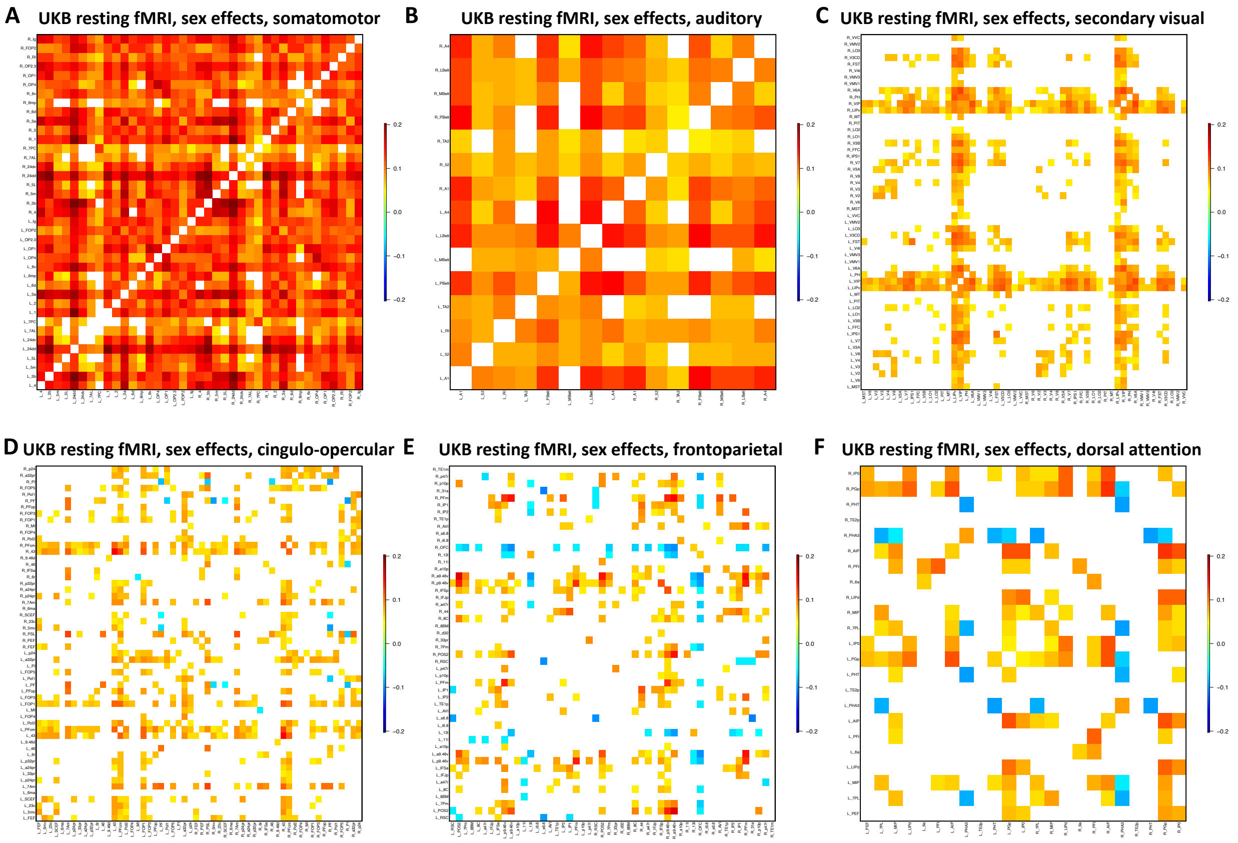


**Fig. S27 Sex effect patterns in selected functional networks of resting fMRI.**

We illustrated the effects passing the Bonferroni significance level ($7.73 \times 10^{-7}$, 0.05/64,620) in the discovery dataset ($n$ = 33,795) and also being significant at the nominal significance level (0.05) in the validation dataset ($n$ = 5, 961).

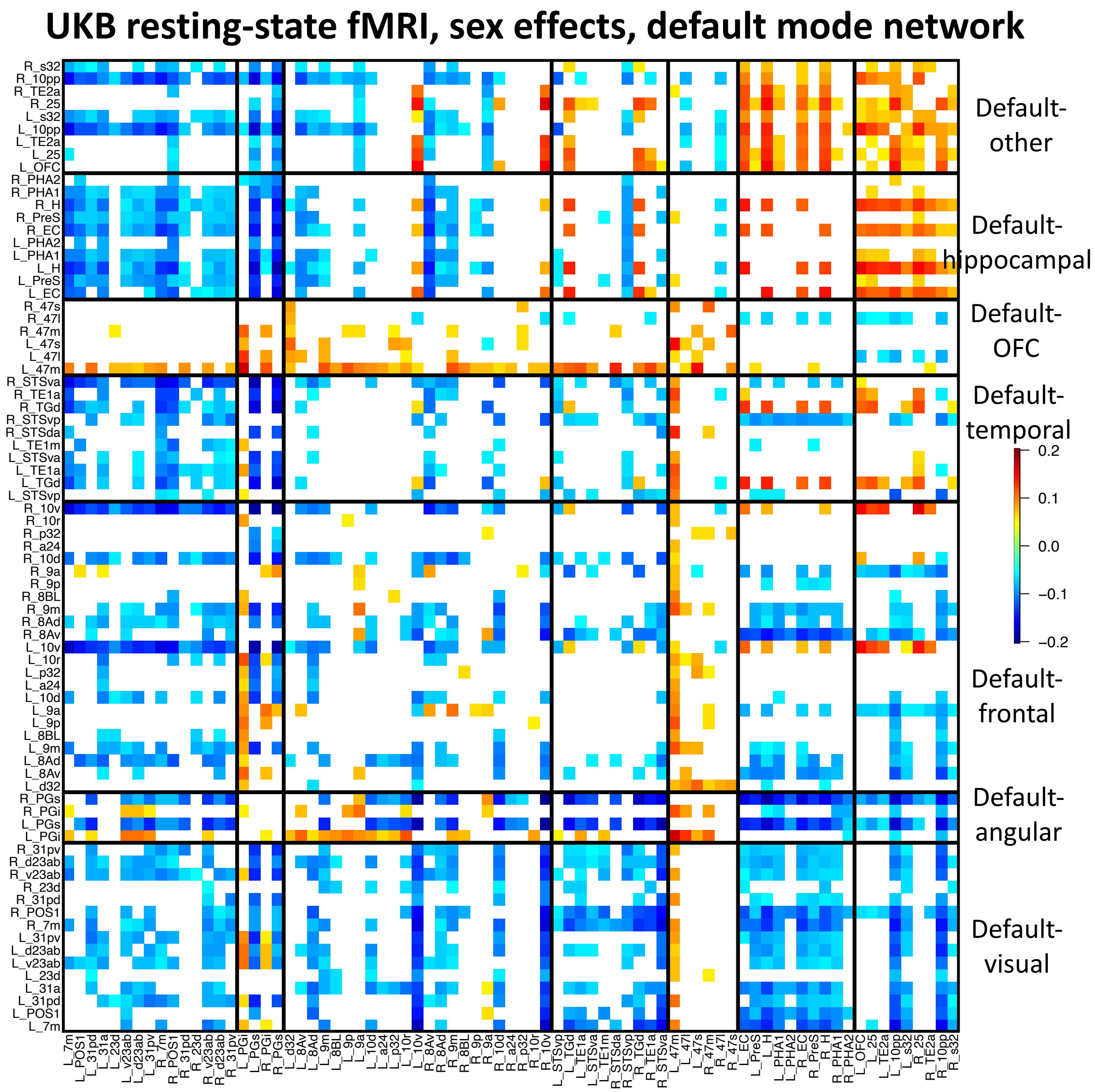


**Fig. S28 Sex effect pattern in the default mode network of resting fMRI.**

We illustrated the effects passing the Bonferroni significance level ($7.73 \times 10^{-7}$, 0.05/64,620) in the discovery dataset ($n$ = 33,795) and also being significant at the nominal significance level (0.05) in the validation dataset ($n$ = 5, 961).

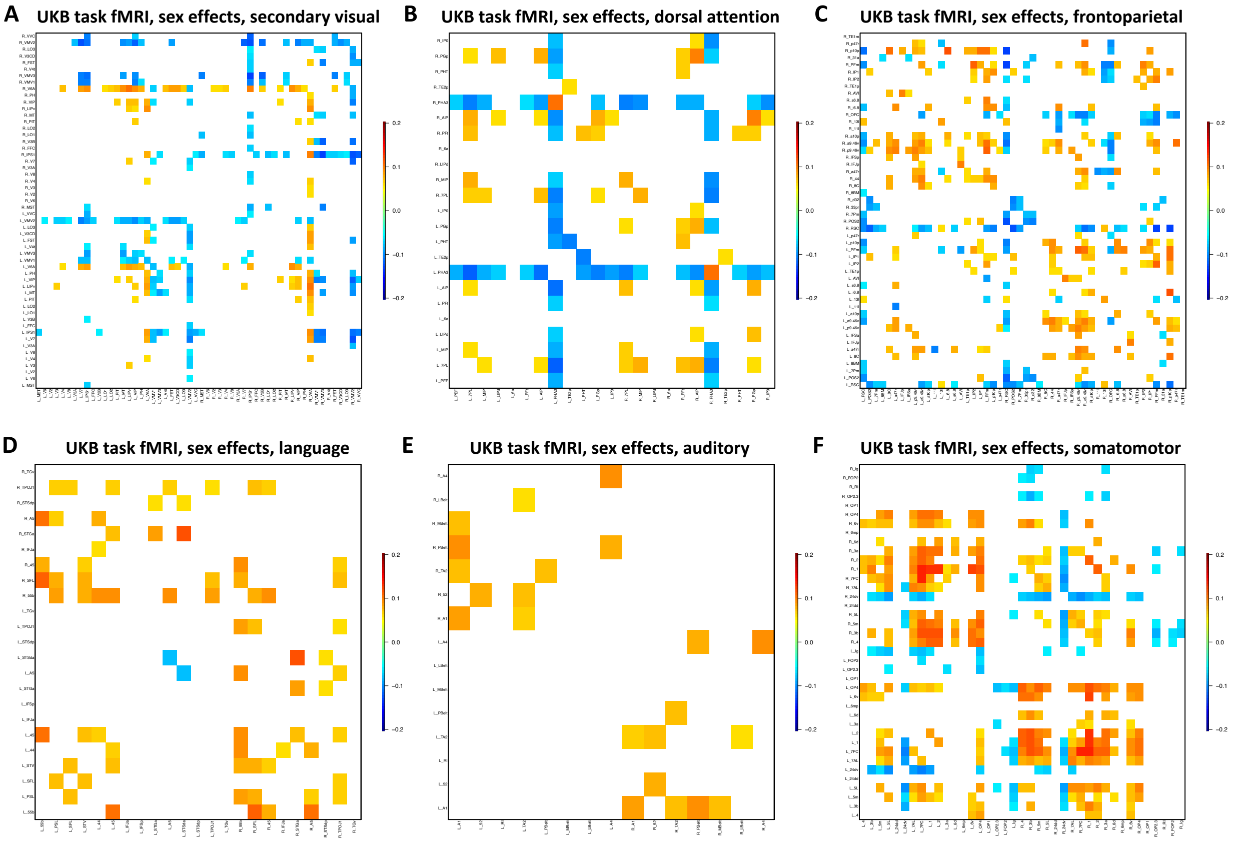


**Fig. S29 Sex effect patterns in selected functional networks of task fMRI.**

We illustrated the effects passing the Bonferroni significance level ($7.73 \times 10^{-7}$, 0.05/64,620) in the discovery dataset ($n$ = 28, 907) and also being significant at the nominal significance level (0.05) in the validation dataset ($n$ = 4, 884).

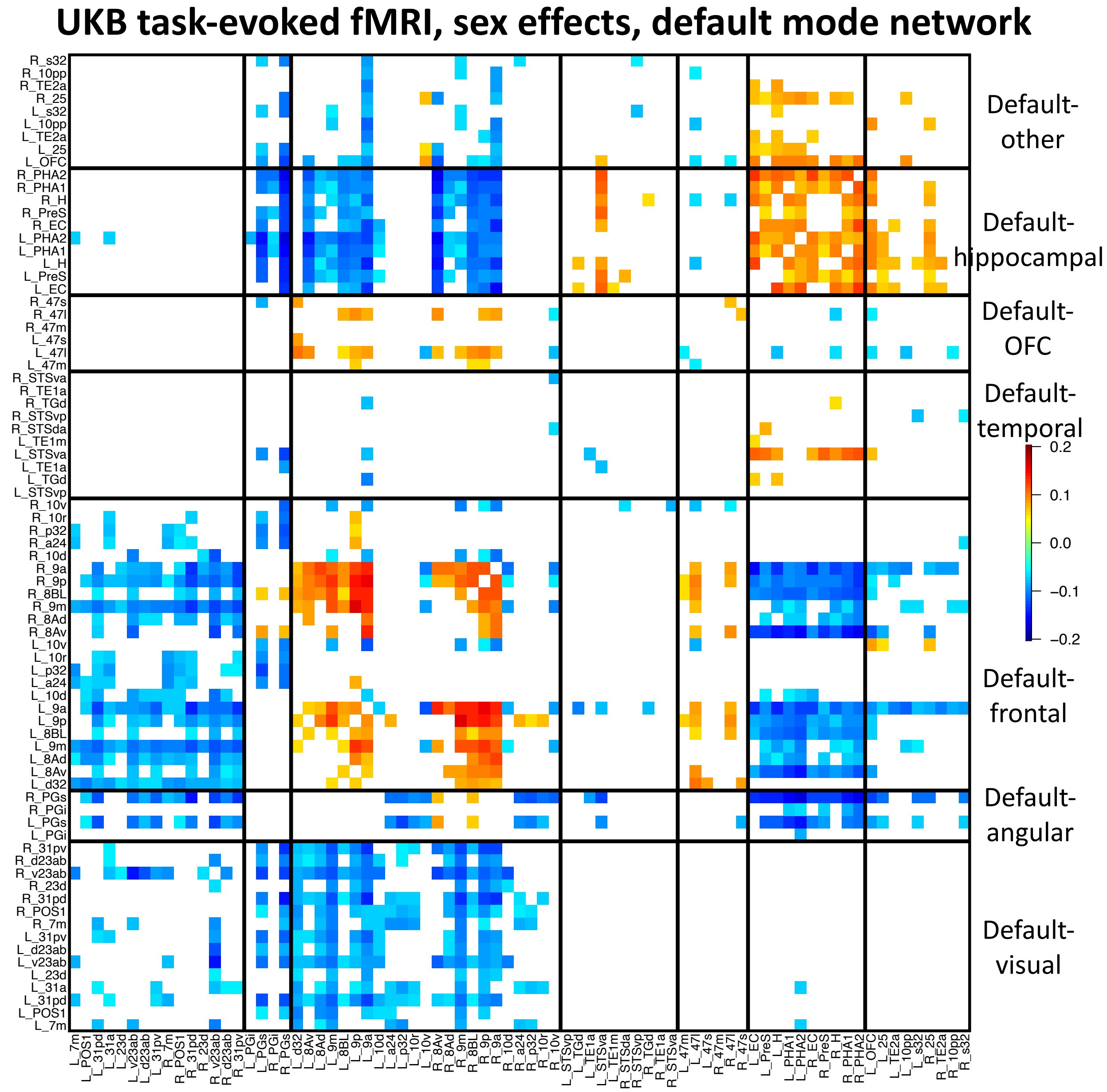


**Fig. S30 Sex effect pattern in the default mode network of task fMRI.**

We illustrated the effects passing the Bonferroni significance level ($7.73 \times 10^{-7}$, 0.05/64,620) in the discovery dataset ($n$ = 28, 907) and also being significant at the nominal significance level (0.05) in the validation dataset ($n$ = 4, 884).

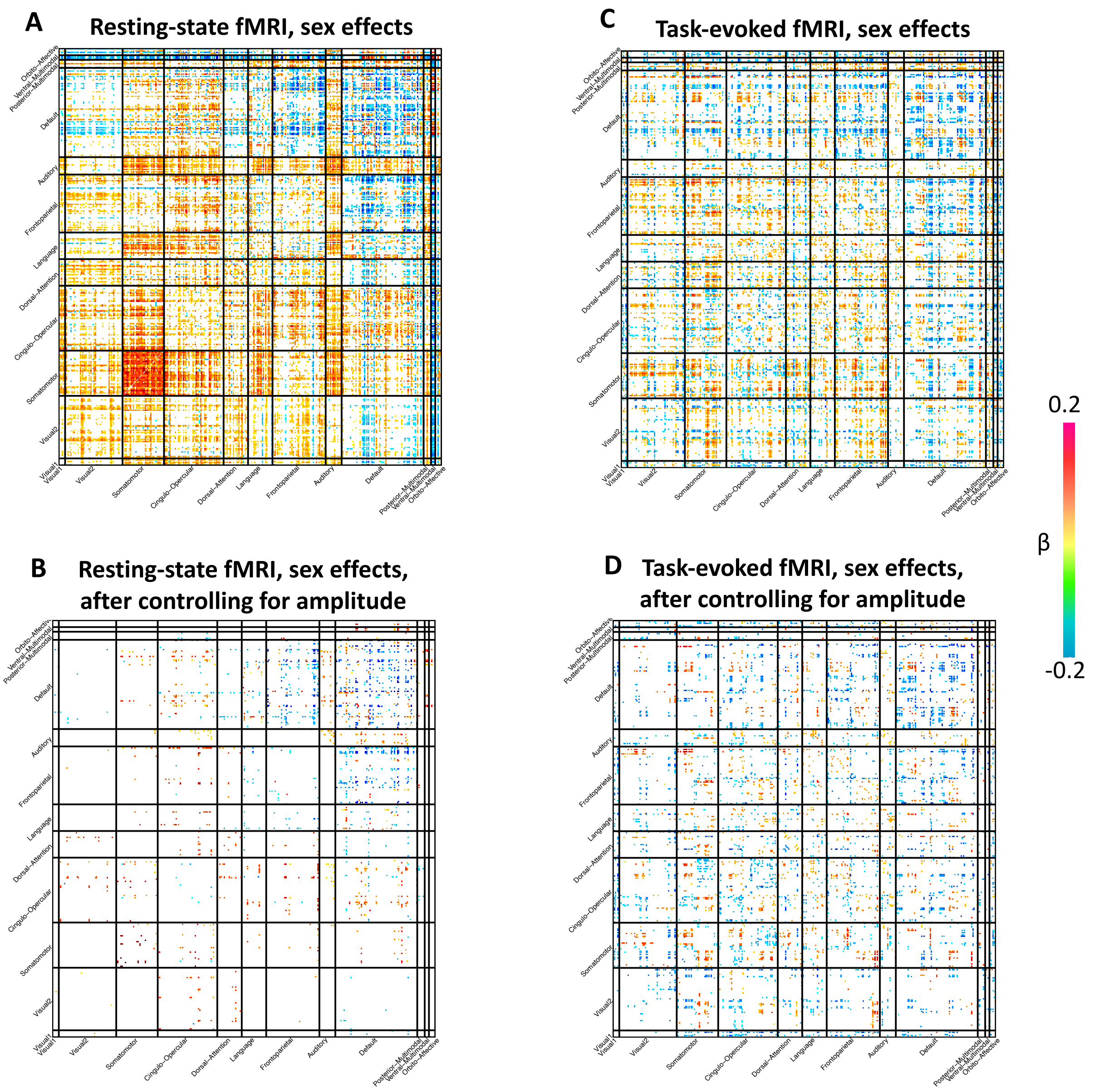


**Fig. S31 Sex effect patterns before and after controlling for amplitude traits.**

In A, we illustrated the sex effects in resting fMRI passing the Bonferroni significance level ($7.73 \times 10^{-7}$, 0.05/64,620) in the discovery dataset ($n$ = 33,795) and also being significant at the nominal significance level (0.05) in the validation dataset ($n$ = 5, 961). In B, we illustrated the remaining significant sex effects in resting fMRI after additionally controlling for amplitude traits. Similarly, the panel C illustrated the significant sex effects in task fMRI and D illustrated the remaining significant sex effects in task fMRI after additionally controlling for amplitude traits.

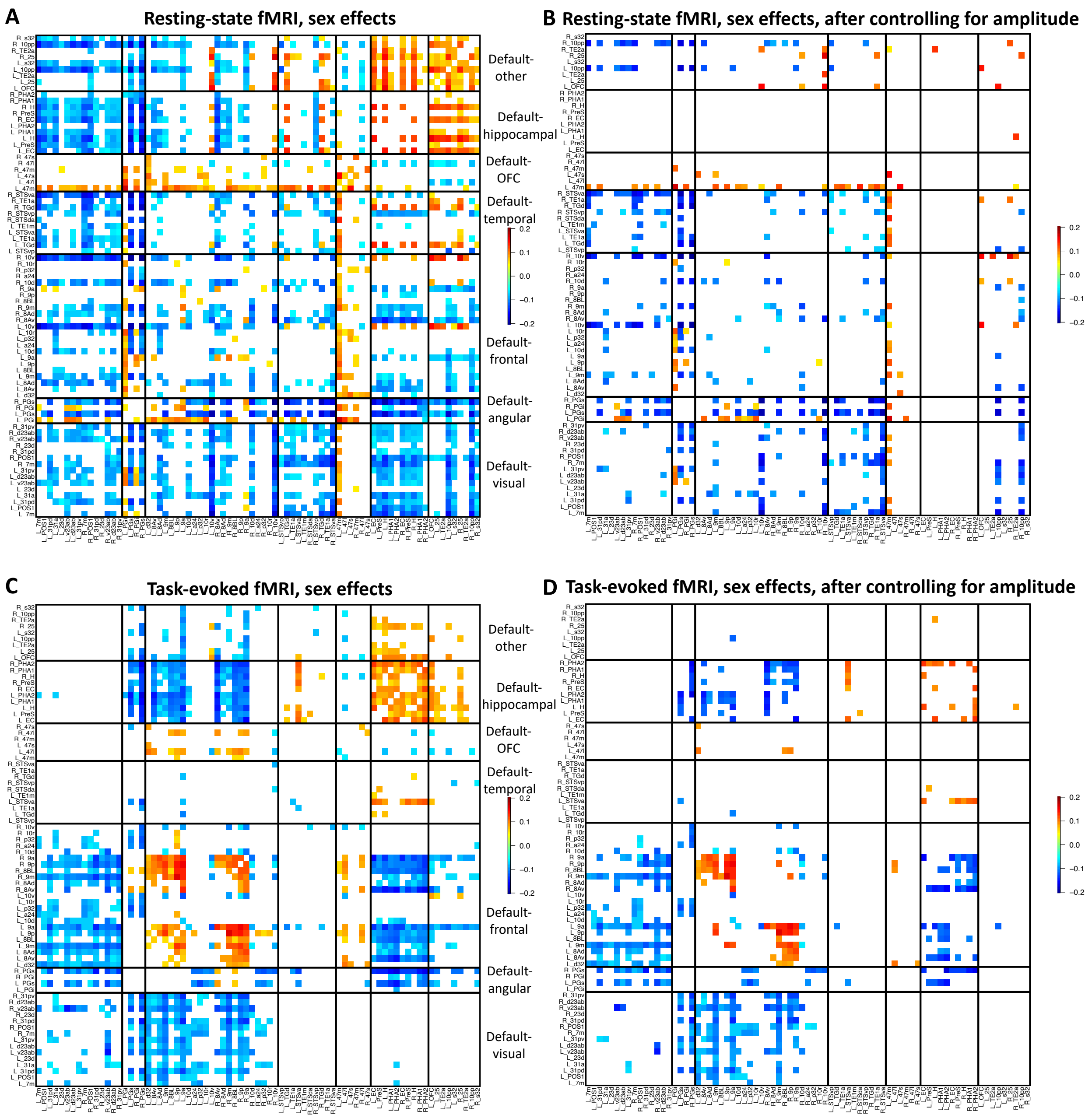


**Fig. S32 Sex effect pattern in the default mode network before and after controlling for amplitude traits.**

In A, we illustrated the sex effects in the default mode network of resting fMRI passing the Bonferroni significance level ($7.73 \times 10^{-7}$, 0.05/64,620) in the discovery dataset ($n$ = 33,795) and also being significant at the nominal significance level (0.05) in the validation dataset ($n$ = 5, 961). In B, we illustrated the remaining significant sex effects in the default mode network of resting fMRI after additionally controlling for amplitude traits. Similarly, the panel C illustrated the significant sex effects in the default mode network of task fMRI and D illustrated the remaining significant sex effects in the default mode network of task fMRI after additionally controlling for amplitude traits.

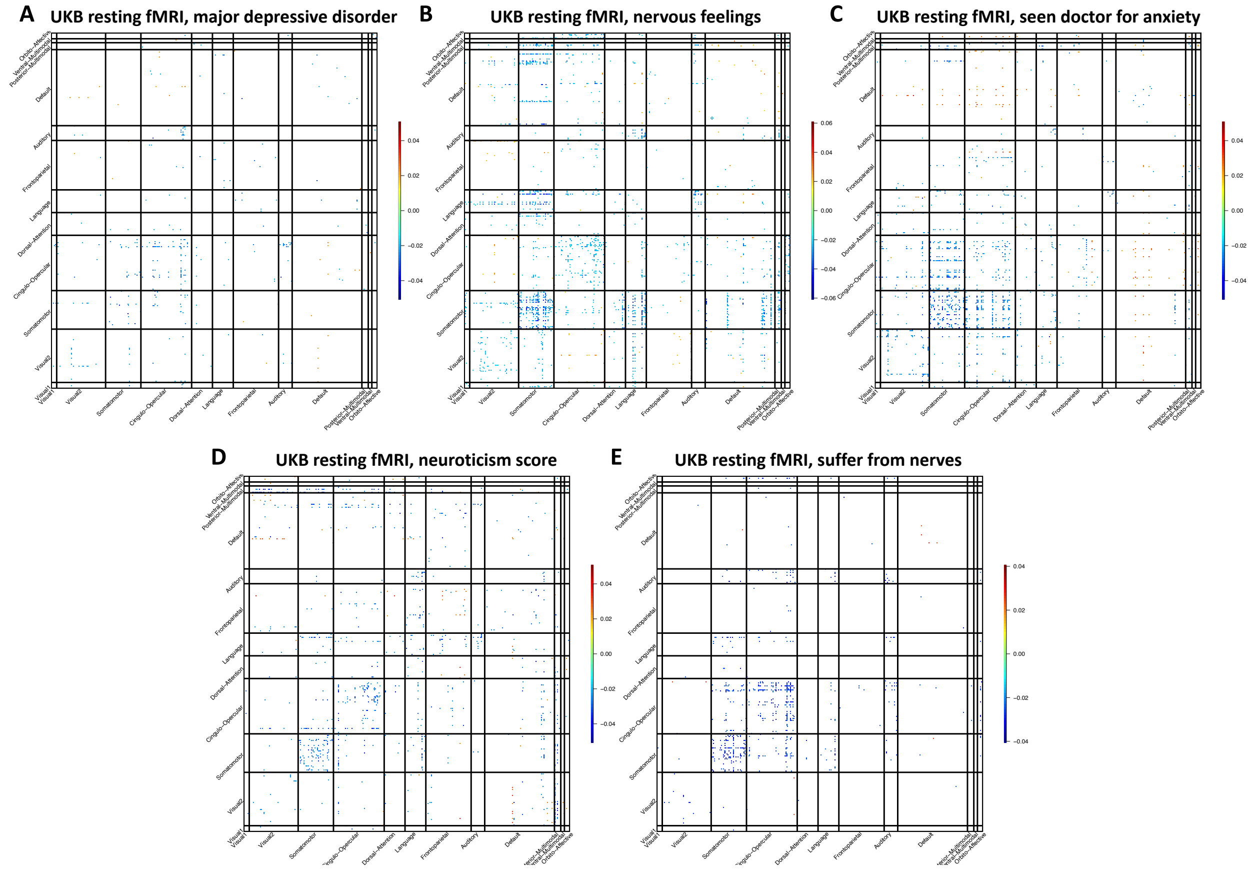


**Fig. S33 Associations between resting fMRI and selected mental health traits.**

We illustrated the correlation coefficients that were significant at FDR 5% level in the discovery dataset (*n* = 33,795) and were also significant at the nominal significance level (0.05) in the validation dataset (*n* = 5, 961). Major depressive disorder, ICD-10 code F329; Nervous feelings, *"Would you call yourself a nervous person?"* (Data field 1970); Seen doctor for anxiety, seen doctor for nerves anxiety tension or depression (Data field 2090); Neuroticism score, summary score of neuroticism, based on 12 neurotic behaviour domains (Data field 20127); and suffer from nerves, *"Do you suffer from 'nerves'?"* (Data field 2010).

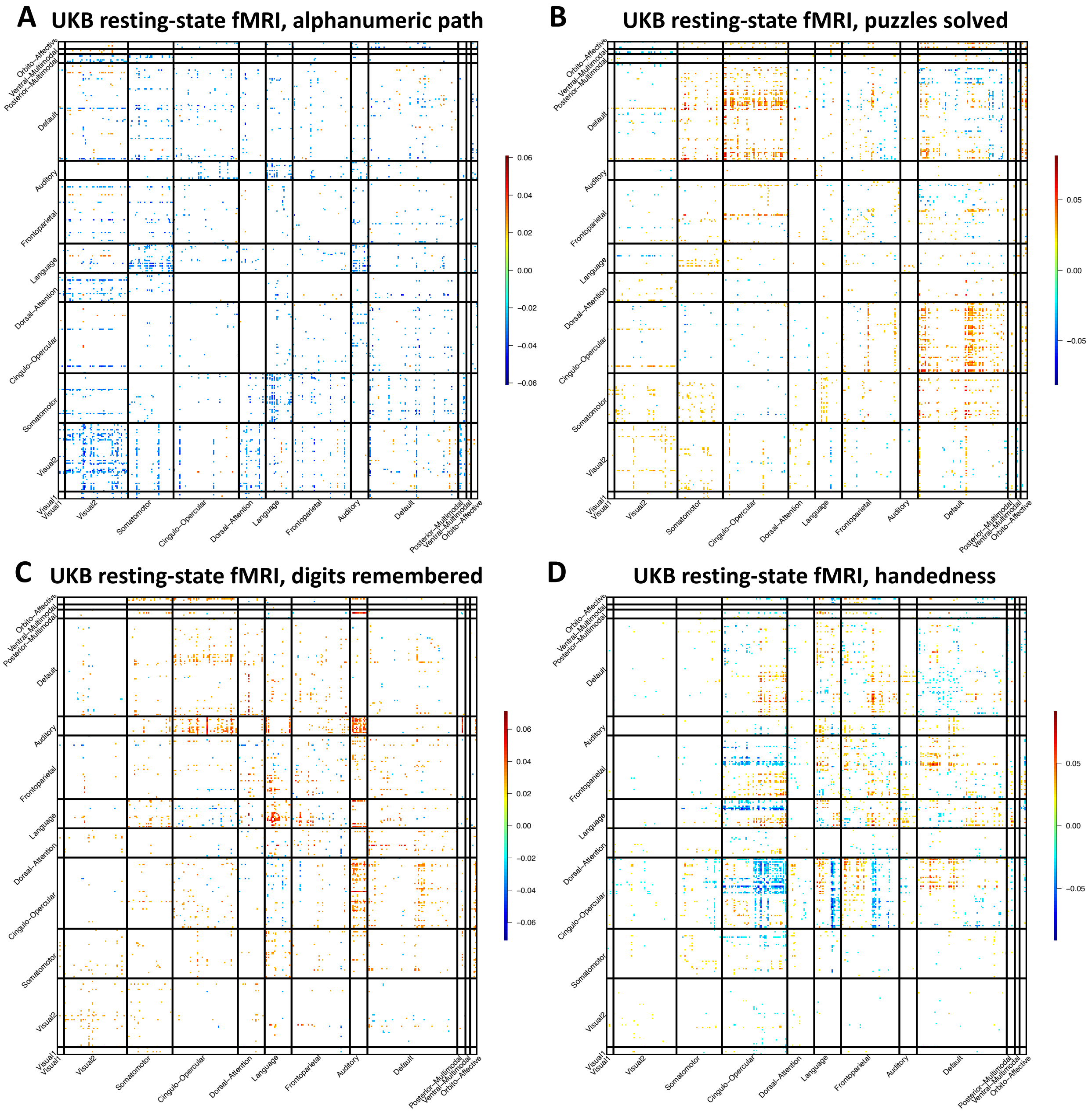


**Fig. S34 Associations between resting fMRI and selected brain-related complex traits.**

We illustrated the correlation coefficients that were significant at FDR 5% level in the discovery dataset ($n$ = 33,795) and were also significant at the nominal significance level (0.05) in the validation dataset ($n$ = 5, 961). Alphanumeric path, duration to complete alphanumeric path (Data field 6350); Puzzles solved, the number of puzzles correctly solved (Data field 6373); digits remembered, maximum digits remembered correctly (Data field 4282); and handedness, right or left-handed (chirality/laterality, 1=right-handed and 2=left-handed, Data field 1707)

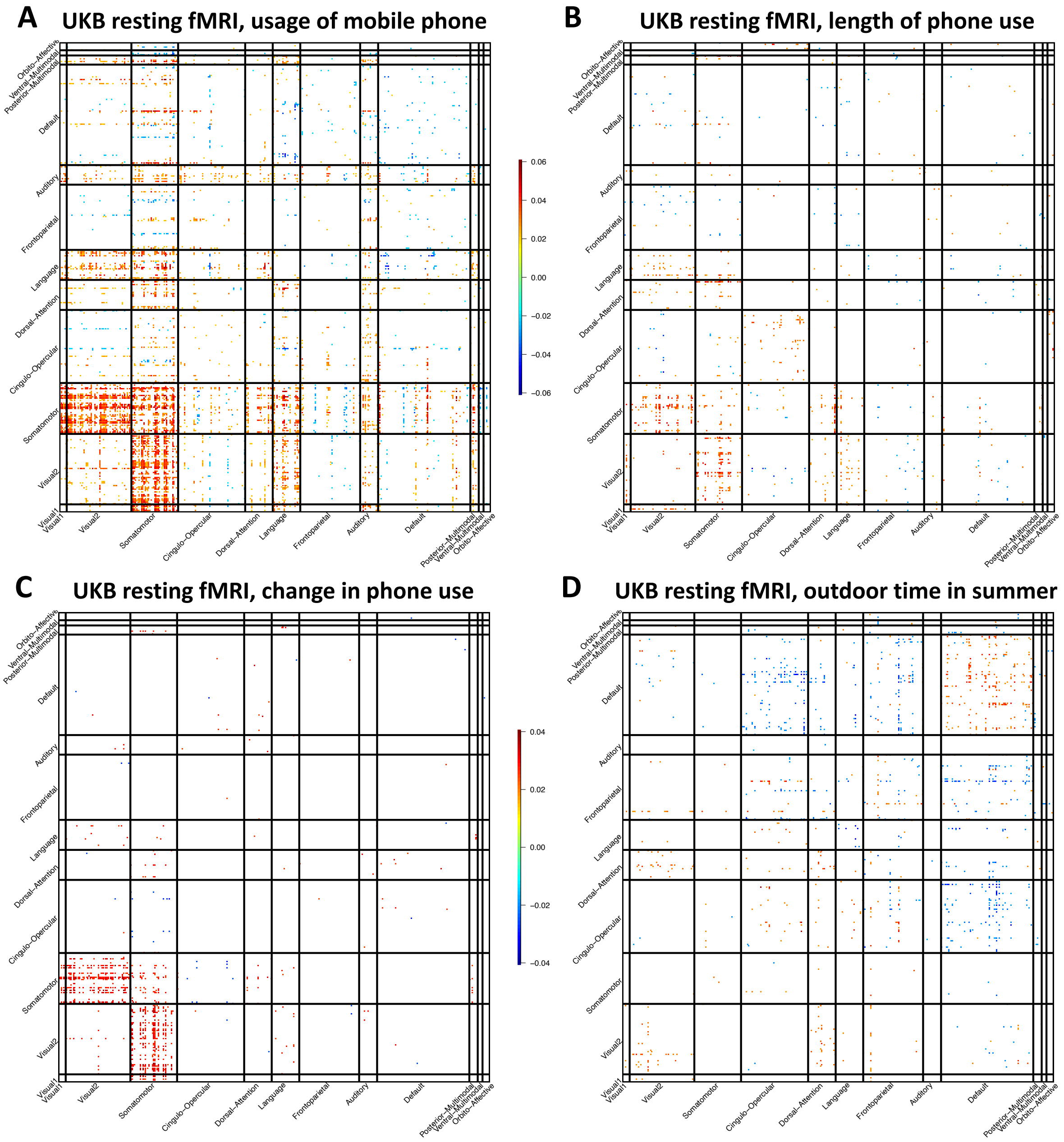


**Fig. S35 Associations between resting fMRI and selected electronic device use and physical activity traits.**

We illustrated the correlation coefficients that were significant at FDR 5% level in the discovery dataset ($n$ = 33,795) and were also significant at the nominal significance level (0.05) in the validation dataset ($n$ = 5, 961). Usage of mobile phone, weekly usage of mobile phone in last 3 months (Data field 1120); Length of phone use, length of mobile phone use (Data field 1110); and change in phone use, difference in mobile phone use compared to two years previously (Data field 1140).

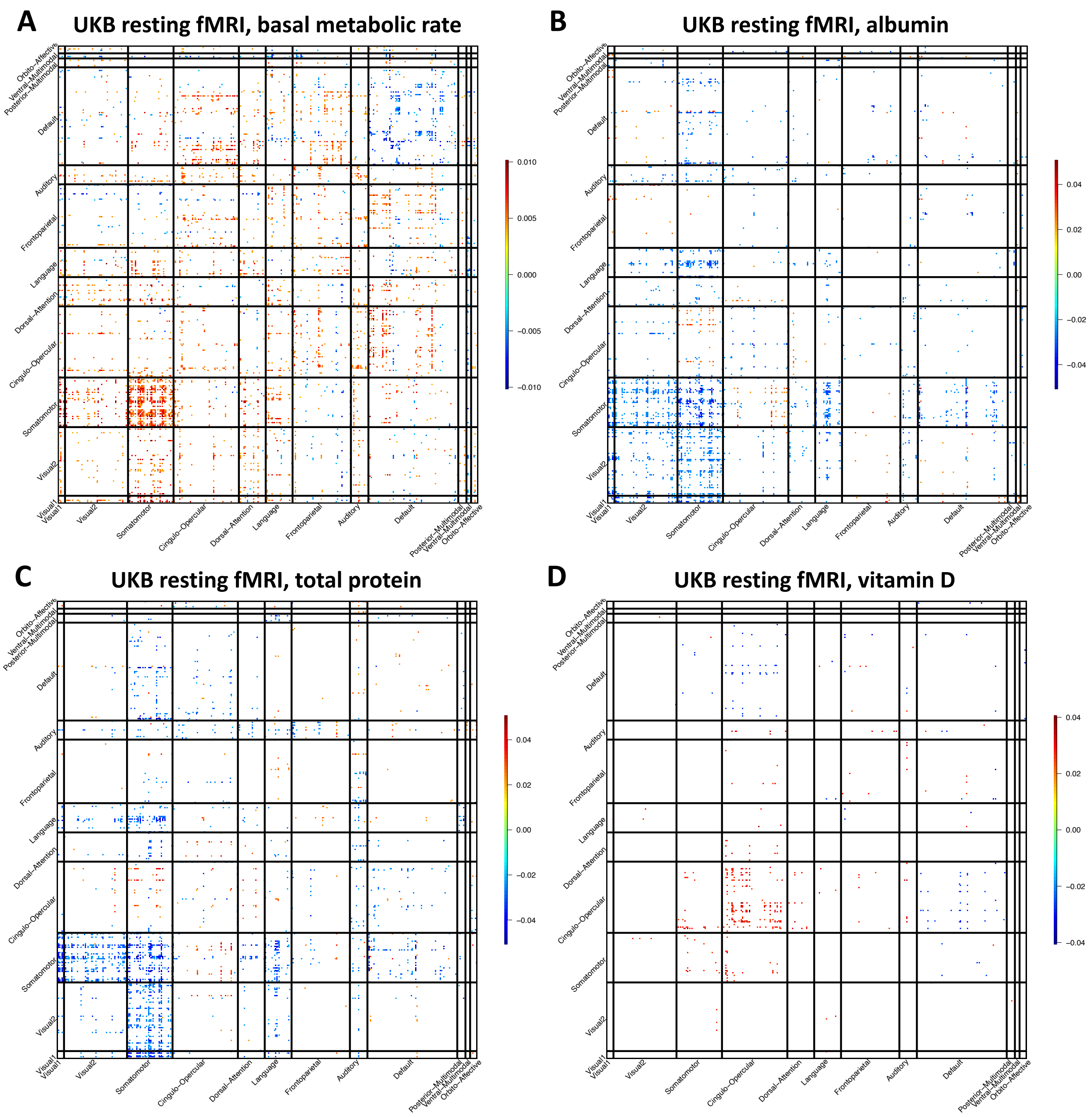


**Fig. S36 Associations between resting fMRI and selected biomarkers.**

We illustrated the correlation coefficients that were significant at FDR 5% level in the discovery dataset ($n$ = 33,795) and were also significant at the nominal significance level (0.05) in the validation dataset ($n$ = 5, 961). The information of the four biomarkers can be found at https://biobank.ndph.ox.ac.uk/showcase/label.cgi?id=18518.

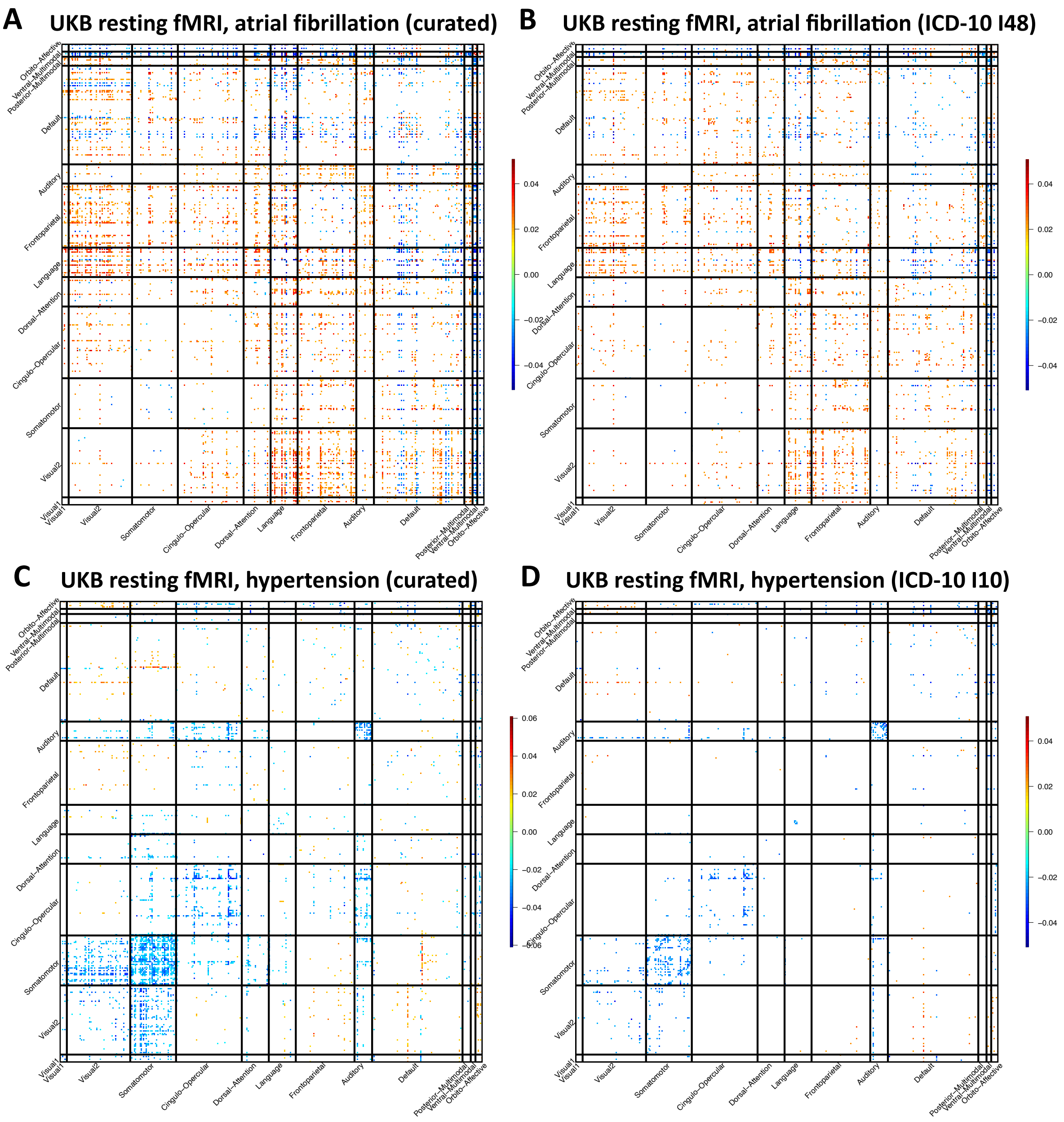


**Fig. S37 Associations between resting fMRI and selected diseases.**

We illustrated the correlation coefficients that were significant at FDR 5% level in the discovery dataset ($n$ = 33,795) and were also significant at the nominal significance level (0.05) in the validation dataset ($n$ = 5, 961). “Curated” indicates curated disease phenotype by combining multiple related ICD-10 codes.

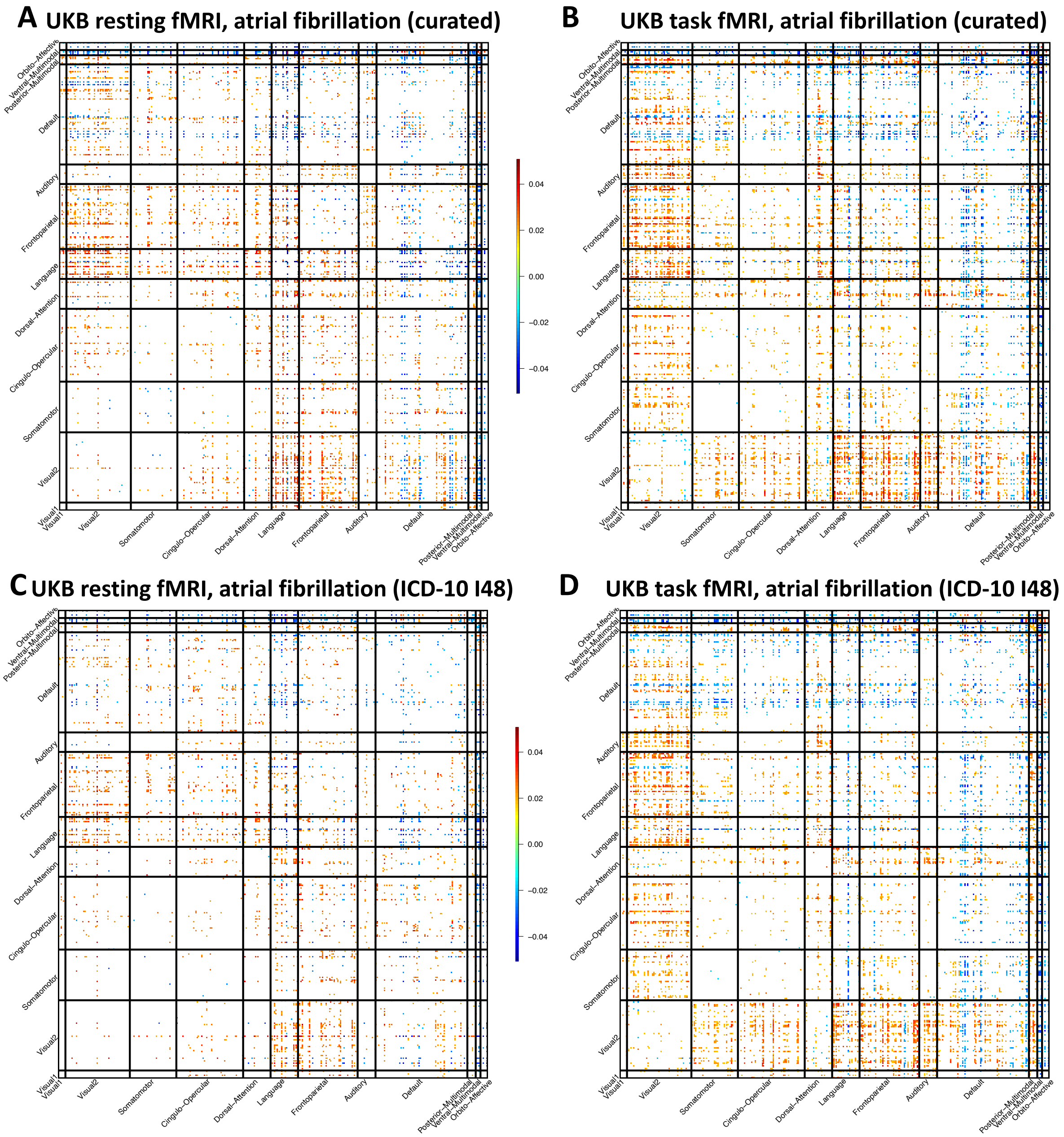


**Fig. S38 Associations between atrial fibrillation and resting and task fMRI.**

We illustrated the correlation coefficients that were significant at FDR 5% level in the discovery dataset (*n* = 33,795 for resting and 28, 907 for task) and were also significant at the nominal significance level (0.05) in the validation dataset (*n* = 5, 961 for resting and 4, 884 for task). “Curated” indicates curated disease phenotype by combining multiple related ICD-10 codes.

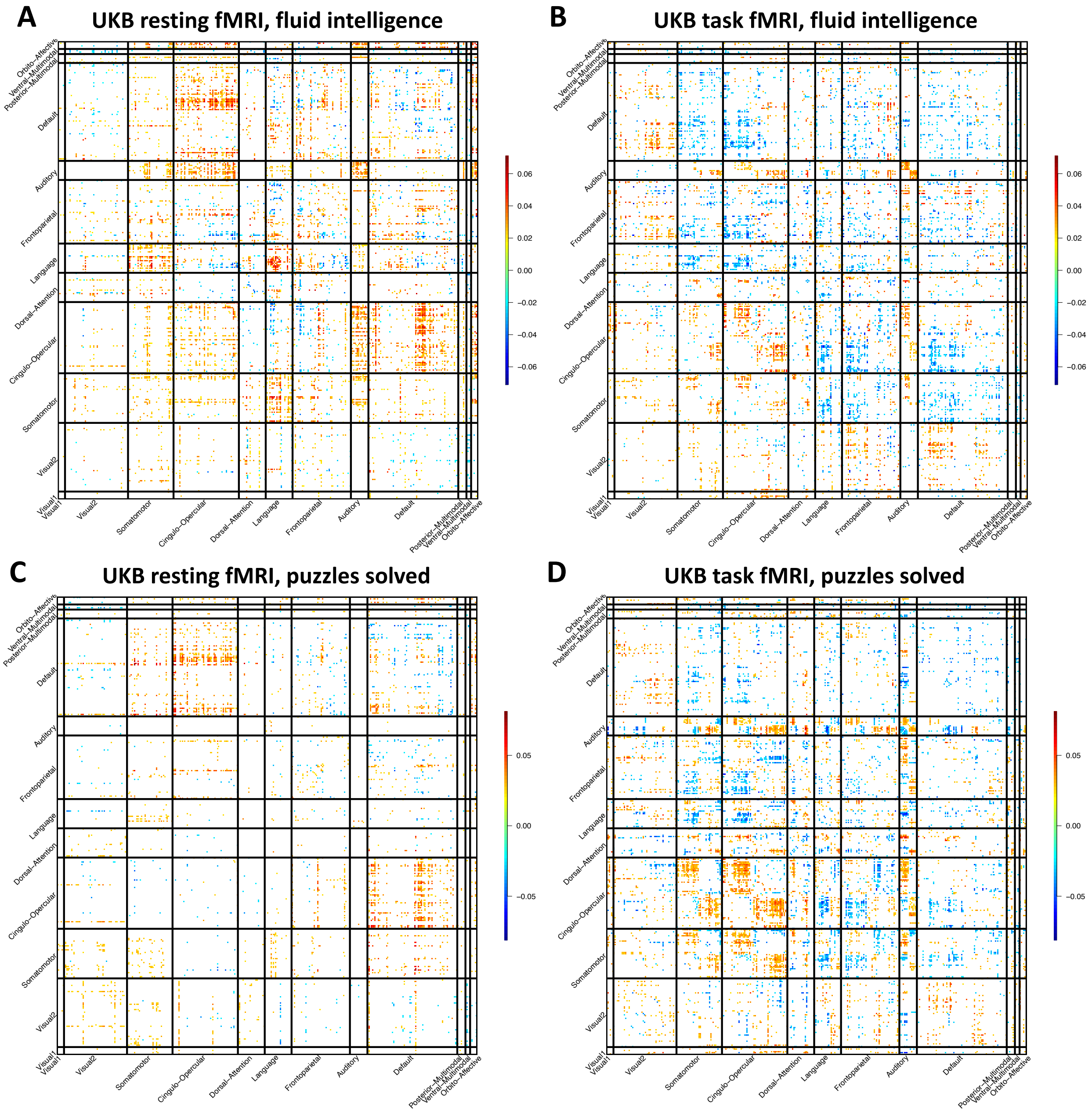


**Fig. S39 Associations between selected cognitive traits and resting and task fMRI.**

We illustrated the correlation coefficients that were significant at FDR 5% level in the discovery dataset (*n* = 33,795 for resting and 28, 907 for task) and were also significant at the nominal significance level (0.05) in the validation dataset (*n* = 5, 961 for resting and 4, 884 for task). Fluid intelligence, fluid intelligence score (Data field 20127); and puzzles solved, the number of puzzles correctly solved (Data field 6373).

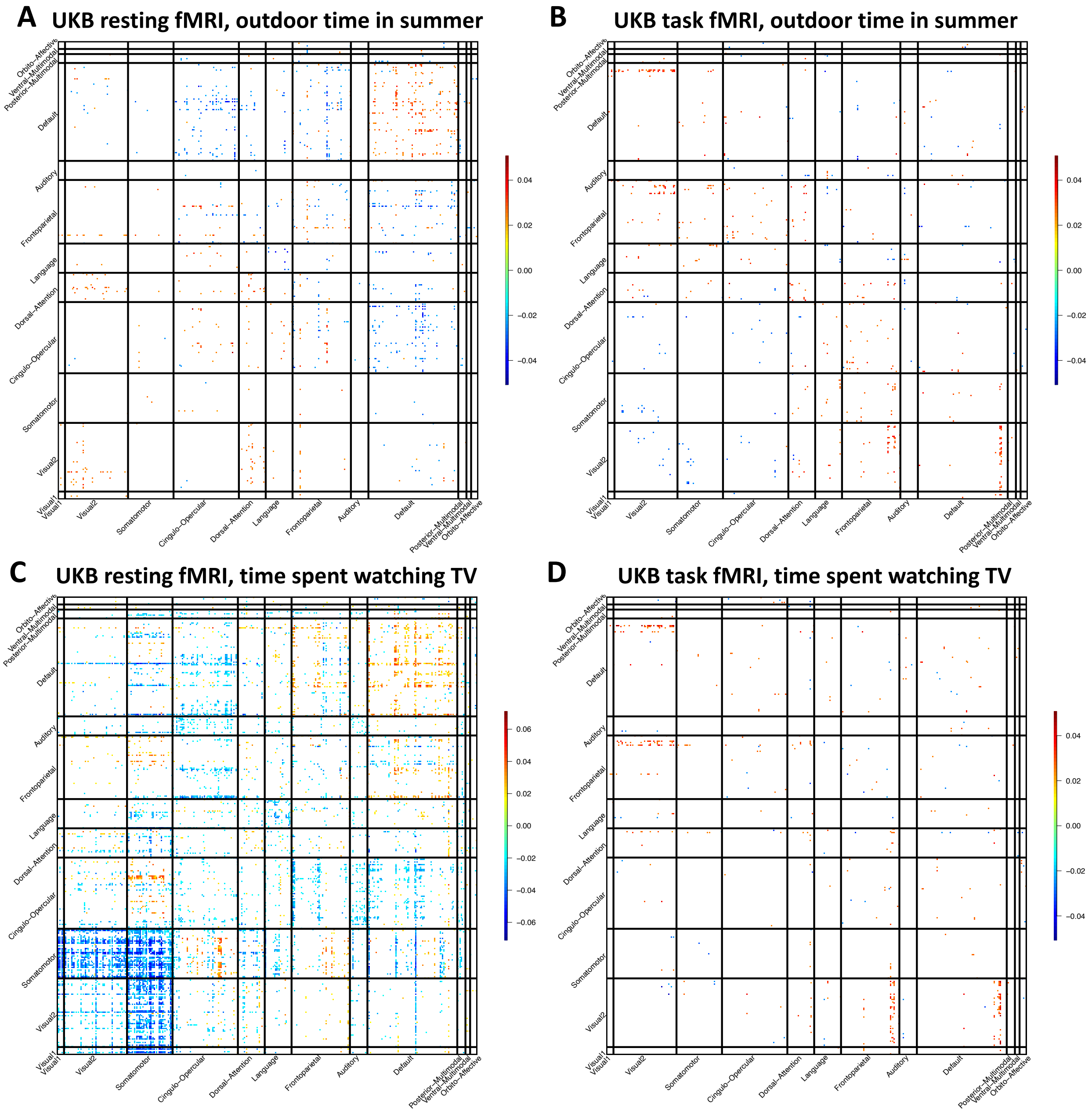


**Fig. S40 Associations between selected physical activity traits and resting and task fMRI.**

We illustrated the correlation coefficients that were significant at FDR 5% level in the discovery dataset ($n$ = 33,795 for resting and 28, 907 for task) and were also significant at the nominal significance level (0.05) in the validation dataset ($n$ = 5, 961 for resting and 4, 884 for task).

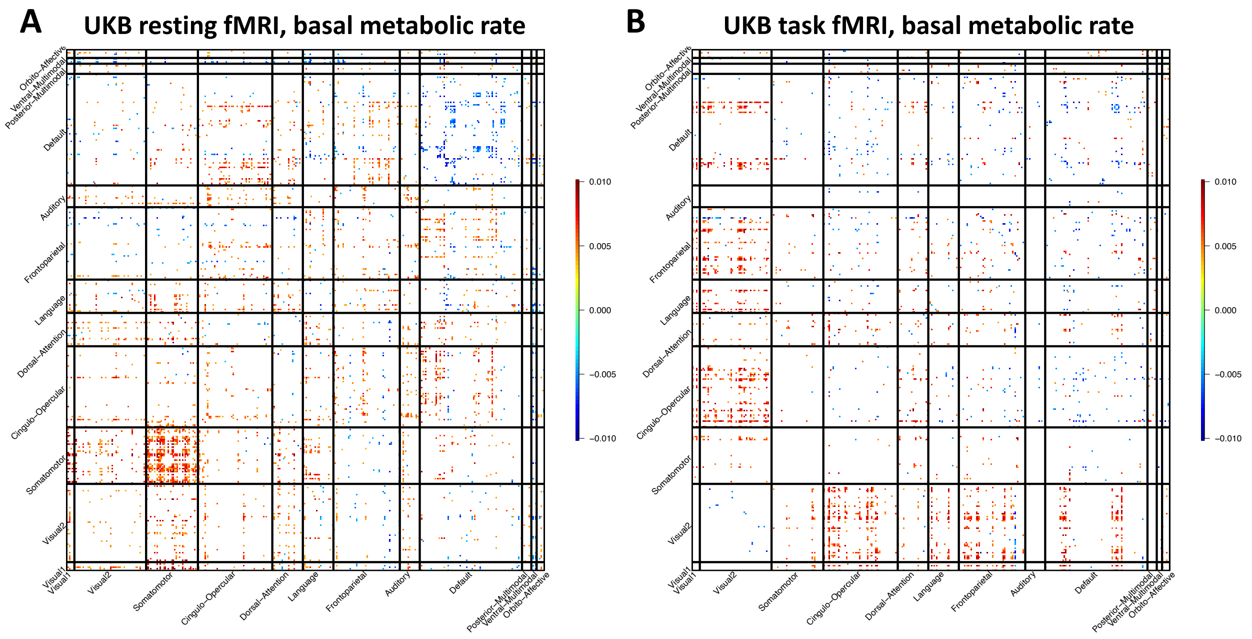


**Fig. S41 Associations between selected biomarker (basal metabolic rate) and resting and task fMRI.**

We illustrated the correlation coefficients that were significant at FDR 5% level in the discovery dataset ($n$ = 33,795 for resting and 28, 907 for task) and were also significant at the nominal significance level (0.05) in the validation dataset ($n$ = 5, 961 for resting and 4, 884 for task).

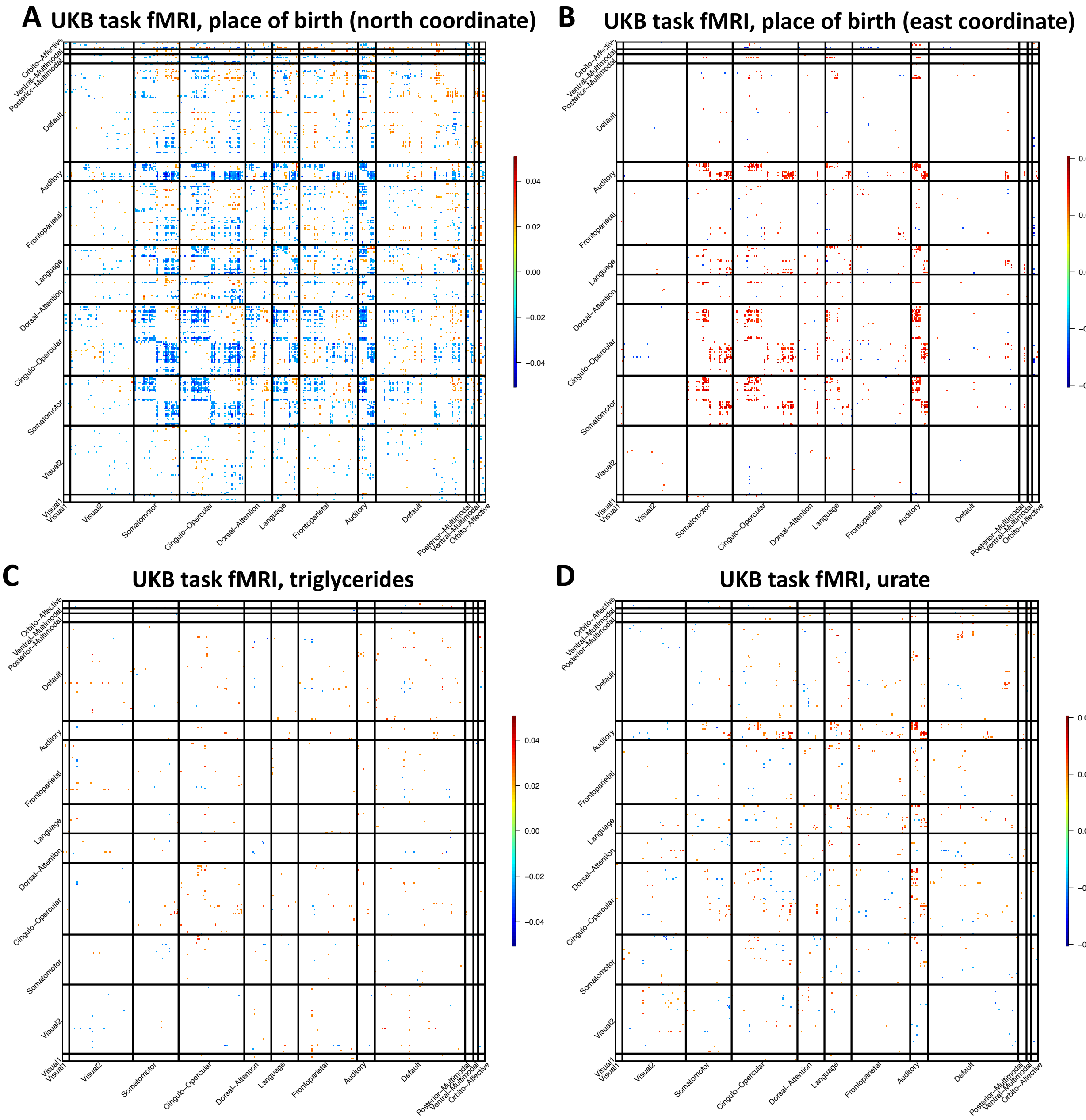


**Fig. S42 Associations between selected traits and task fMRI.**

We illustrated the correlation coefficients that were significant at FDR 5% level in the discovery dataset ($n$ = 28, 907) and were also significant at the nominal significance level (0.05) in the validation dataset ($n$ = 4, 884).

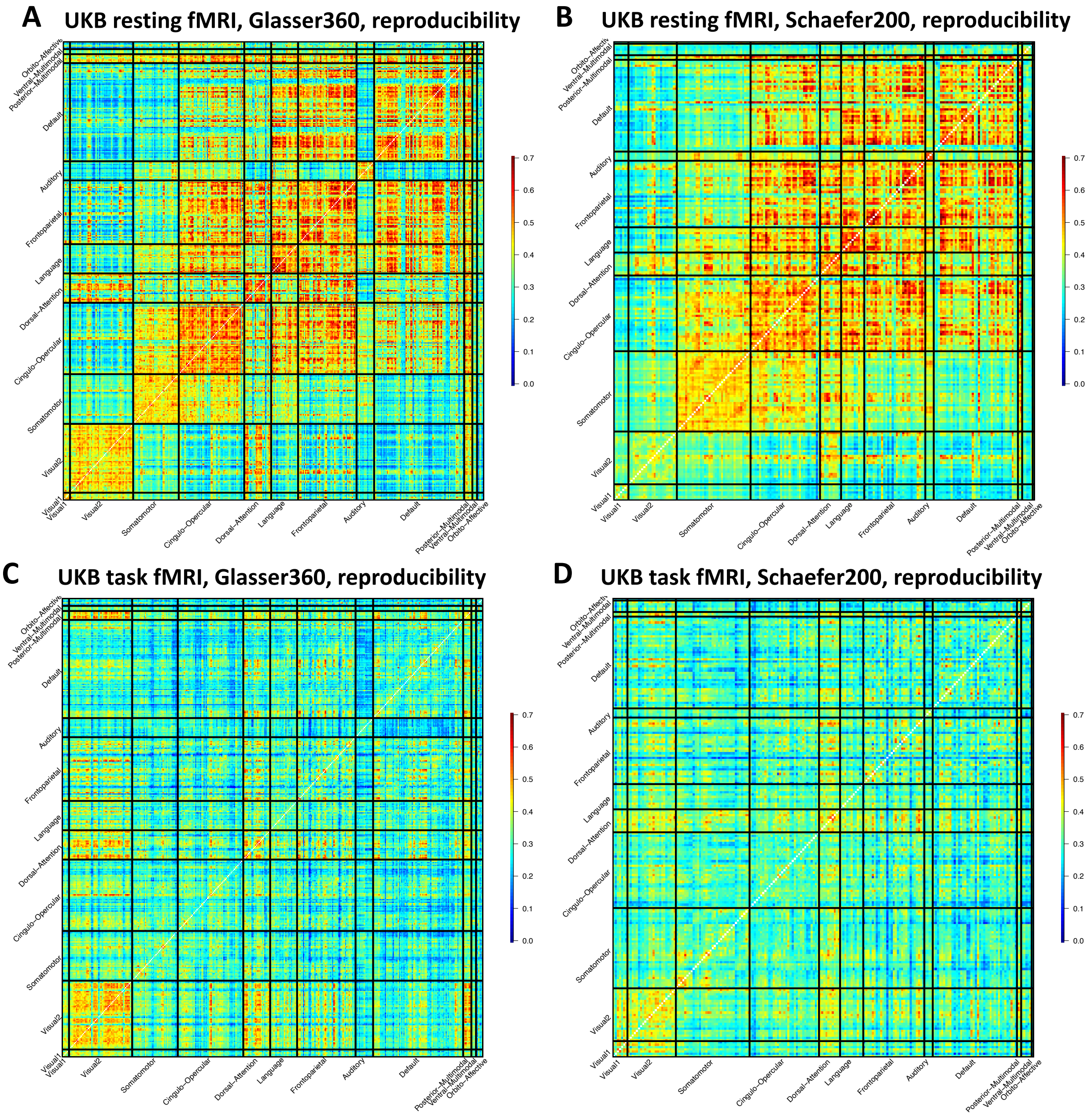


**Fig. S43 Comparation of the reproducibility spatial patterns in the Glasser360 and Schaefer200 atlases.**

The sample size of the UKB repeat imaging visit dataset was 2,771 for resting fMRI and 2,014 for task fMRI.

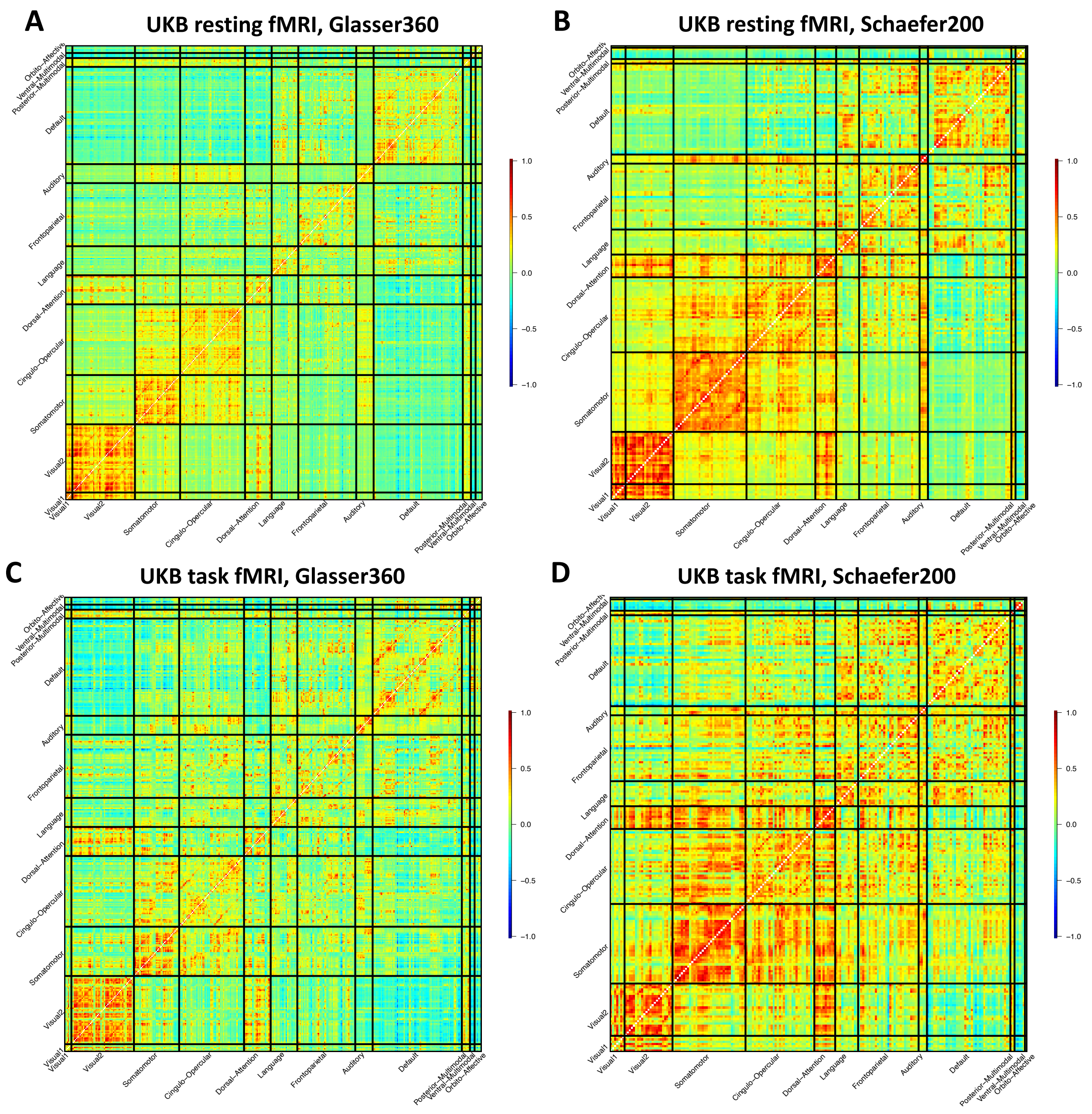


**Fig. S44 Comparation of the group mean spatial patterns in the Glasser360 and Schaefer200 atlases.**

The sample size was 37,794 subjects for resting fMRI and 32,144 subjects for task fMRI. We calculated the group average for each functional connectivity across all subjects.

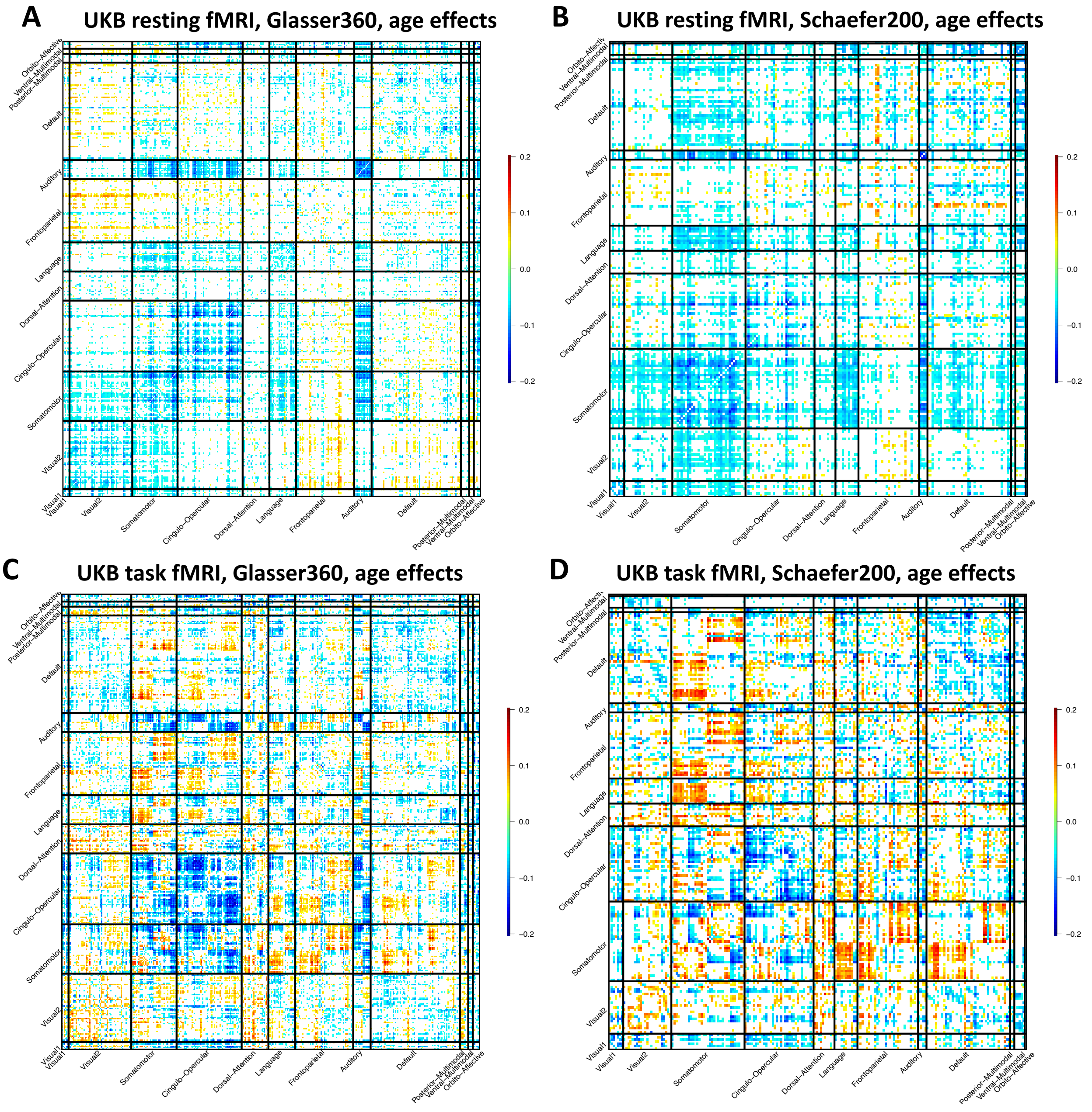


**Fig. S45 Comparation of the age effect patterns in the Glasser360 and Schaefer200 atlases.**

We illustrated the effects passing the Bonferroni significance level (64,620 tests for Glasser360 and 19,900 tests for Schaefer200) in the discovery dataset (*n* = 33,795 for resting and 28, 907 for task) and also being significant at the nominal significance level (0.05) in the validation dataset (*n* = 5, 961 for resting and 4, 884 for task).

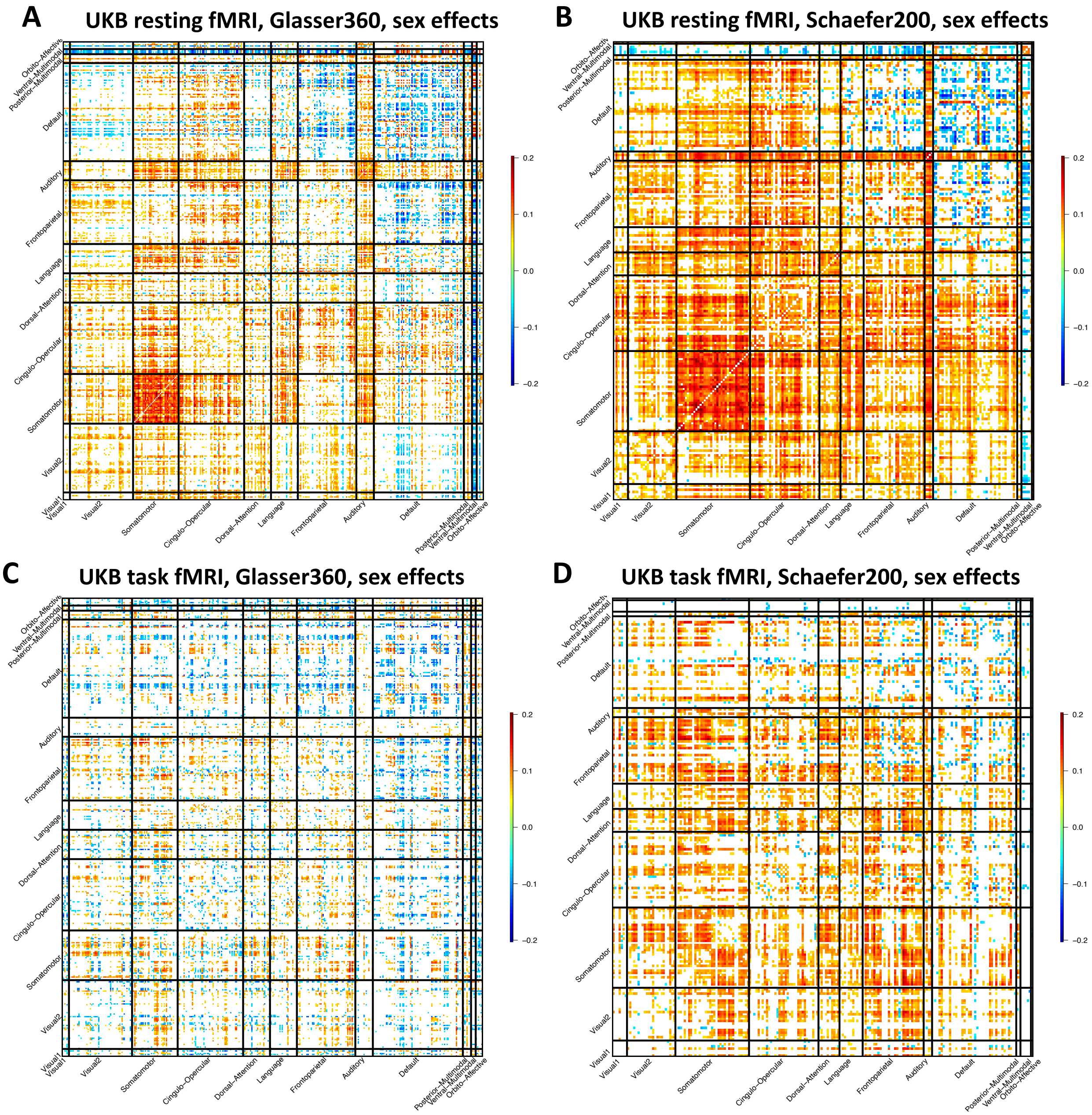


**Fig. S46 Comparation of the sex effect patterns in the Glasser360 and Schaefer200 atlases.**

We illustrated the effects passing the Bonferroni significance level (64,620 tests for Glasser360 and 19,900 tests for Schaefer200) in the discovery dataset (*n* = 33,795 for resting and 28, 907 for task) and also being significant at the nominal significance level (0.05) in the validation dataset (*n* = 5, 961 for resting and 4, 884 for task).

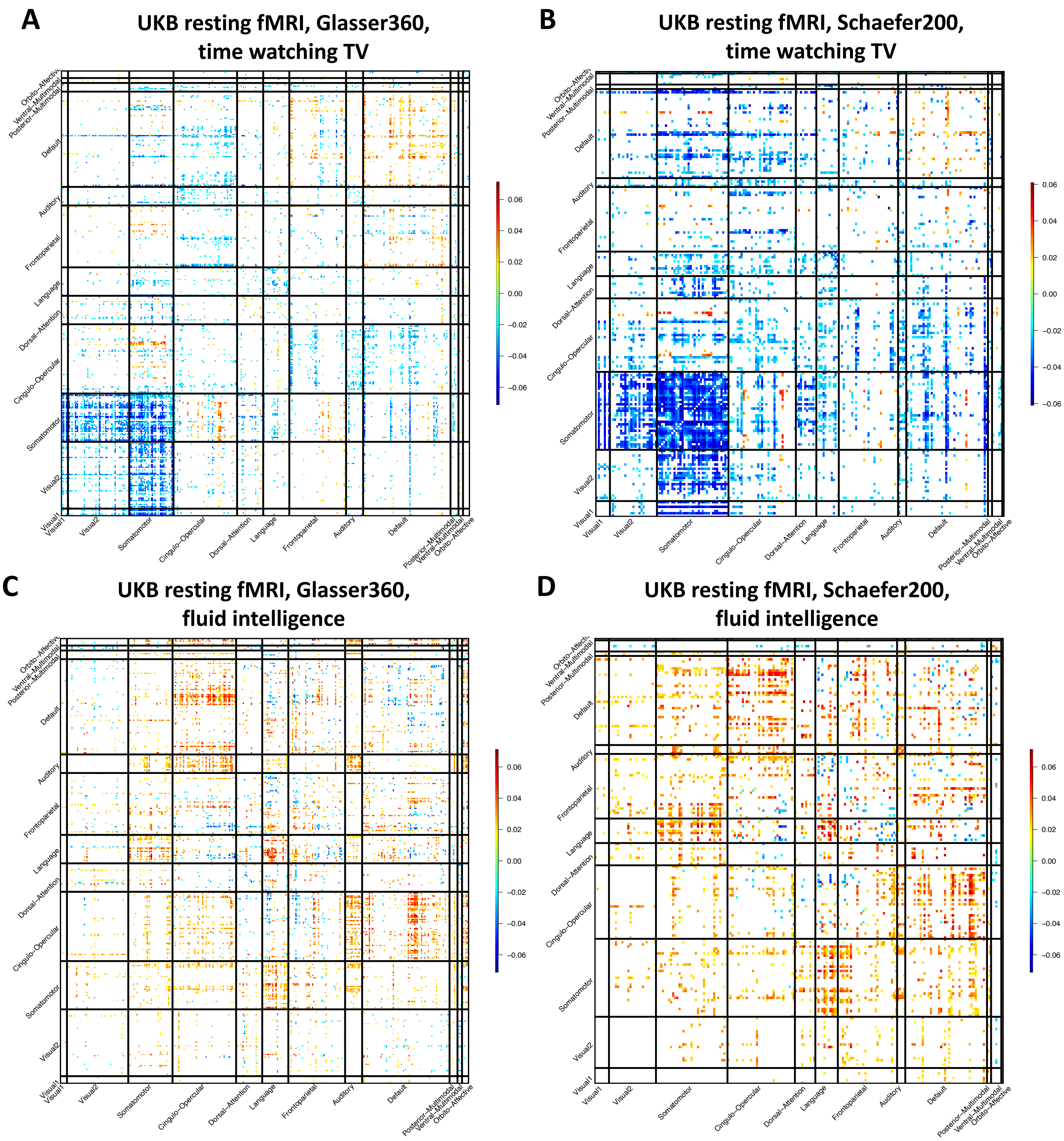


**Fig. S47 Comparation of association patterns in the Glasser360 and Schaefer200 atlases.**

We illustrated the correlation coefficients that were significant at FDR 5% level in the discovery dataset (n = 33,795) and were also significant at the nominal significance level (0.05) in the validation dataset (n = 5, 961). Time watching TV, time spent watching TV (Data field 1070); and fluid intelligence, fluid intelligence score (Data field 20127).

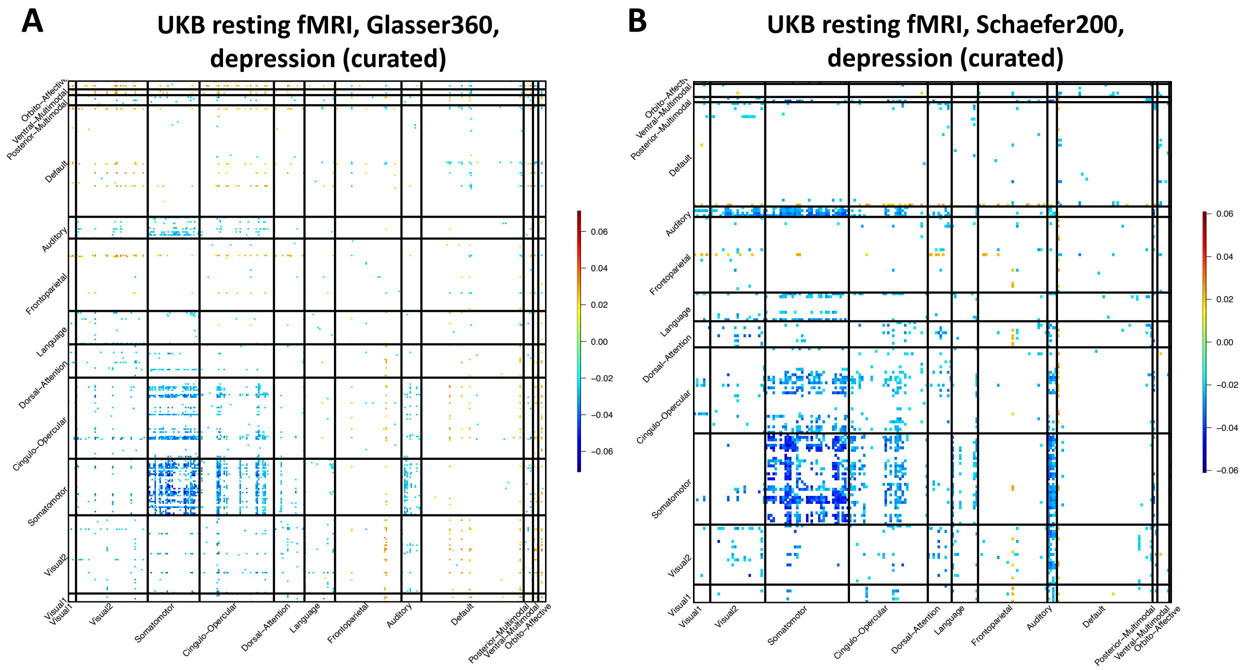


**Fig. S48 Comparation of association patterns in the Glasser360 and Schaefer200 atlases.**

We illustrated the correlation coefficients that were significant at FDR 5% level in the discovery dataset (n = 33,795) and were also significant at the nominal significance level (0.05) in the validation dataset (n = 5, 961). Time watching TV, time spent watching TV (Data field 1070); and fluid intelligence, fluid intelligence score (Data field 20127).

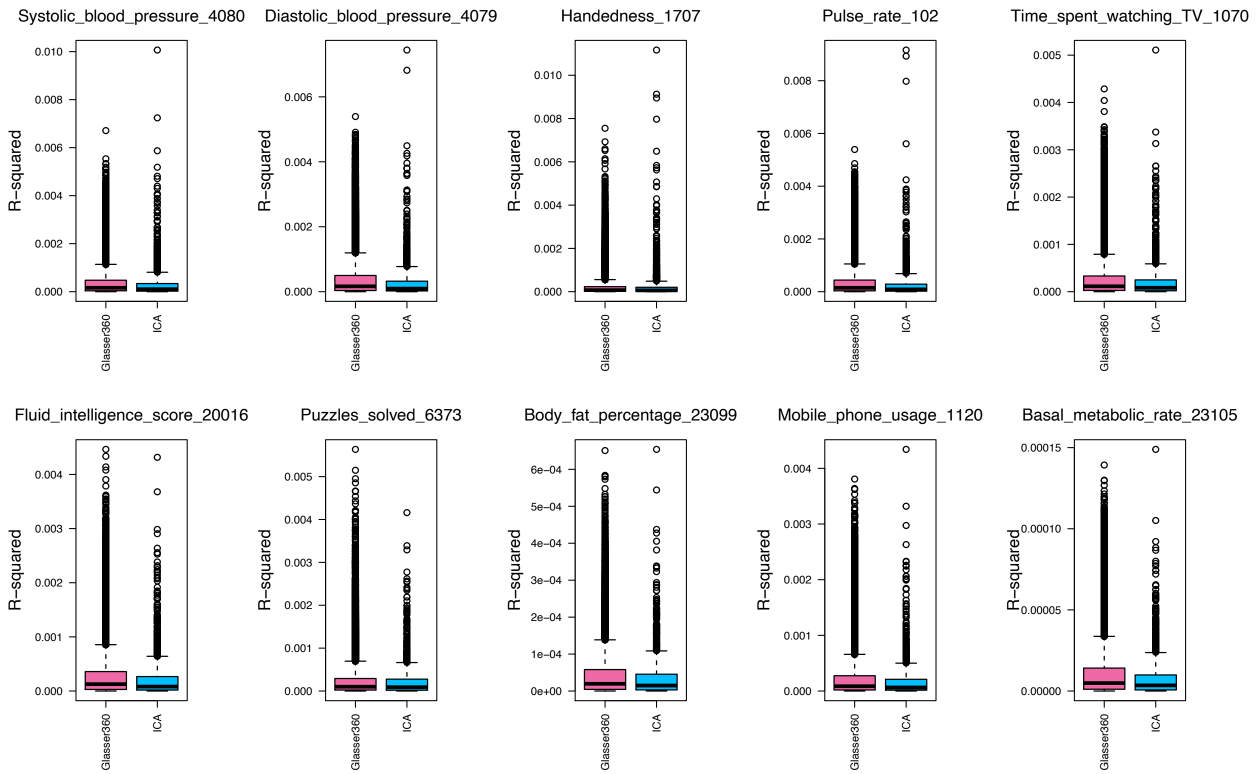


**Fig. S49 Comparation of selected trait associations with the Glasser360 atlas traits and whole brain ICA traits.**

We ranked all traits by their associations with the while brain ICA functional connectivity traits and plotted the top 10 ranked traits. In this figure, we illustrated the r-squared of the associations with the traits from the Glasser360 atlas traits (Glasser360) and whole brain ICA traits (ICA).

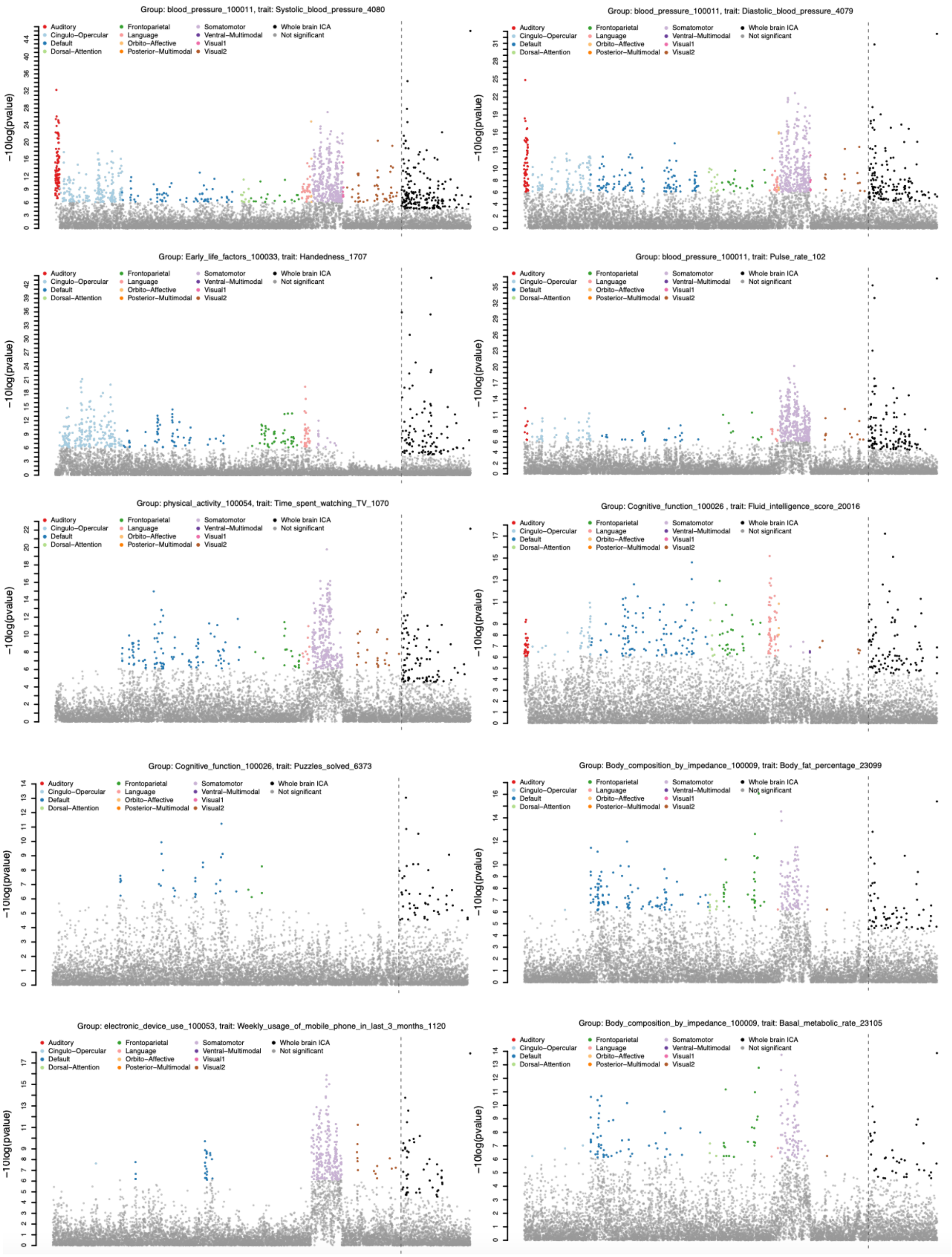


**Fig. S50 Comparation of selected trait associations with the Glasser360 atlas traits and whole brain ICA traits.**

We illustrated the -log10(pvalue) of the associations with the traits from the Glasser360 atlas traits and whole brain ICA traits. Bonferroni-significant traits are highlighted with colors. We ranked all traits by their associations with the while brain ICA functional connectivity traits and plotted the top 10 ranked traits.

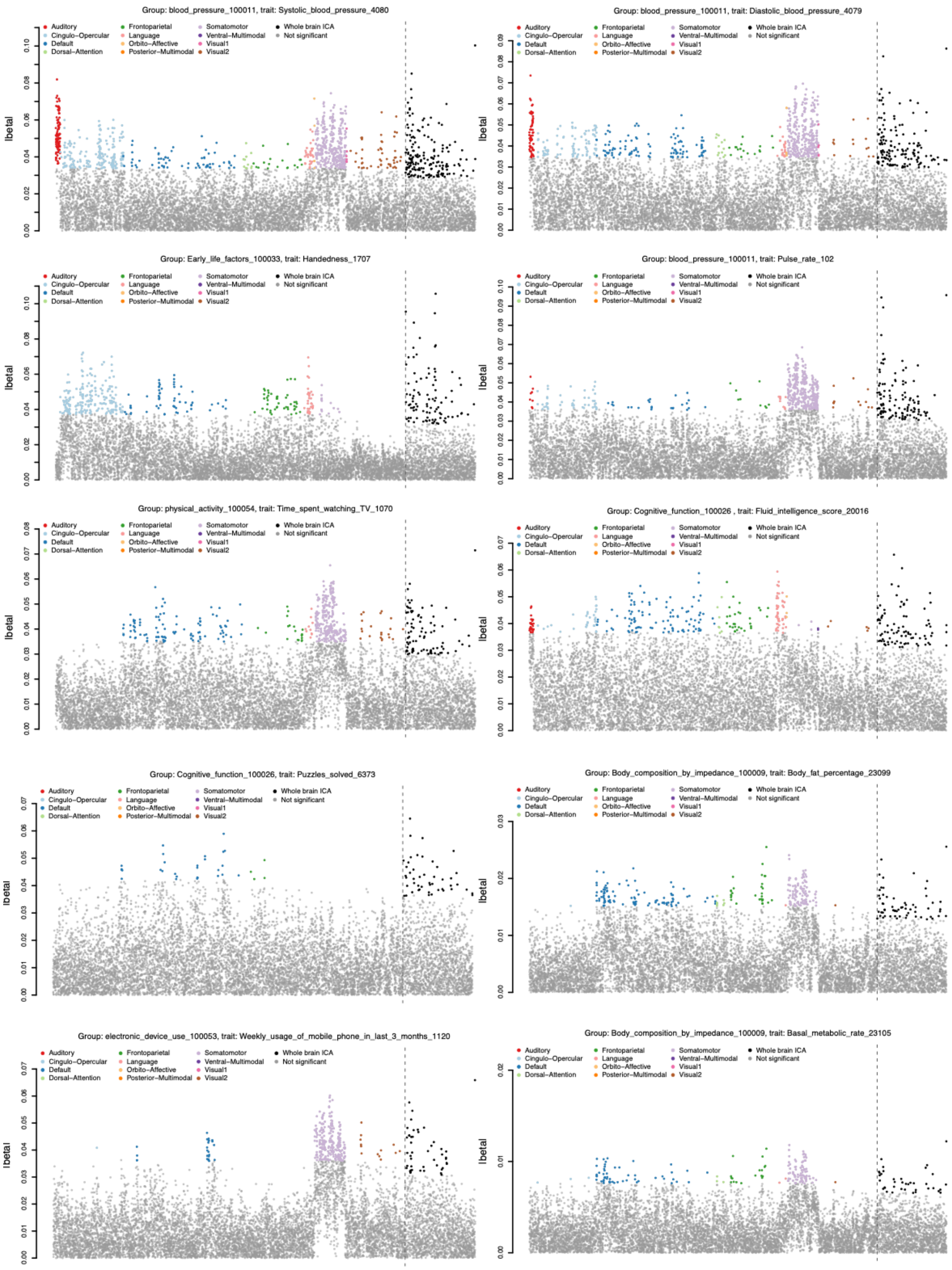


**Fig. S51 Comparation of selected trait associations with the Glasser360 atlas traits and whole brain ICA traits.**

We illustrated the absolute value of the regression coefficients of the associations with the traits from the Glasser360 atlas traits and whole brain ICA traits. Bonferroni-significant traits are highlighted with colors. We ranked all traits by their associations with the while brain ICA functional connectivity traits and plotted the top 10 ranked traits.
