## Supplementary material for "An atlas of trait associations with resting-state and task-evoked human brain functional architectures in the UK Biobank": supp_info

May 09, 2022

**This PDF file includes:**

Supplementary Note
Captions for Table S1 to S4

**Other Supplementary Information for this manuscript includes the following:**

Figs. S1 to S51 (available in a PDF file)
Tables S1 to S4 (.xlsx, available in a ZIP file)

### Supplementary Note

#### Image acquisition and preprocessing

This work made use of resting-state and task-evoked functional magnetic resonance imaging (fMRI) data from the UK Biobank (UKB) and HCP studies. The image acquisition and preprocessing procedures were detailed in the UK Biobank Brain Imaging Documentation (`https://biobank.ctsu.ox.ac.uk/crystal/crystal/docs/brain_mri.pdf`) for the UKB and in WU-Minn (2017); Smith et al. (2013); Van Essen et al. (2013) for HCP. Below we briefly introduced the image acquisition and preprocessing procedures used in the two studies.

**UKB fMRI acquisition** All UKB brain imaging datasets, including the resting-state fMRI (rsfMRI), the task-evoked fMRI (tfMRI), and the T1-weighted structural MRI (sMRI) data, were acquired from standard Siemens Skyra 3T scanners. The rsfMRI data were acquired at 490 time points and a duration of 6 minutes, each with a 2.4×2.4×2.4 mm spatial resolution at a dimension of 88×88×64. The gradient-echo echo-planar imaging (GE-EPI) was adopted with a multiband factor of 8, no iPAT, flip angle 52°, and fat saturation. The echo time (TE) and repetition time (TR) were 39 ms and 735 ms, respectively. As implemented in the CMRR multiband acquisition (Moeller et al., 2010), a separate "single-band reference scan" was also acquired. This had the same geometry (including EPI distortion) as the time series data, but had higher between-tissue contrast to noise, and was used as the reference scan in head motion correction and alignment to other modalities (Alfaro-Almagro et al., 2018). The tfMRI data adopted the Hariri faces/shapes "emotion" task which is the same as one of the HCP tasks. The data were acquired at 332 time points within a duration of 4 minutes. Other scanning parameters including the spatial resolution and dimension, multiband factor, TR and TE were the same as the UKB rsfMRIs. The T1-weighted sMRI scans were acquired at the isotropic resolution of 1 mm and a dimension of 208×256×256 matrix, with a TR of 2000 ms, an inversion time (TI) of 880 ms, a TE of 2 ms, an in-plane acceleration of 2, and a scan time of 5 min using straight sagittal orientation.

**UKB image preprocessing** The UKB rsfMRI and tfMRI data were preprocessed by the UK Biobank brain imaging team (Alfaro-Almagro et al., 2018). The full pipeline can be found in Section 3 of the UK Biobank Brain Imaging Documentation, referred to as UKB preprocessing pipeline in this note. Overall, the pipeline included three parts: image cleaning, image registration, and representative time series generation. The source codes have been shared by the UK Biobank team at `https://git.fmrib.ox.ac.uk/falmagro/UK_biobank_pipeline_v_1`.

For rsfMRI, the image cleaning workflow in the UKB preprocessing pipeline included the following steps: motion correction using MCFLIRT (Jenkinson et al., 2002); grand-mean intensity normalisation of the entire 4D dataset by a single multiplicative factor; highpass temporal filtering (Gaussian-weighted least-squares straight line fitting, with sigma of 50.0 s); EPI unwarping; GDC unwarping. Finally, structured artefacts were removed by the ICA+FIX processing (i.e., independent component analysis (ICA) followed by FMRIB's ICA-based X-noiseifier (Beckmann and Smith, 2004; Salimi-Khorshidi et al., 2014; Griffanti et al., 2014)). FIX was hand-trained on 40 UKB rsfMRI subjects by the UK Biobank imaging team. The image registration part had the following steps. First, they aligned the GDC unwarped rsfMRI data from the previous step with the high-resolution T1 MRI image. The EPI unwarping in the last step already included an alignment to the T1, though the unwarped data was written out in native (unwarped) fMRI space (and the transform to T1 space written out separately). This T1 alignment was carried out by FLIRT, with a final BBR cost function (Greve and Fischl, 2009). After the fMRI GDC unwarping, a final FLIRT realignment to T1 was applied, which toke into account any shifts resulting from the GDC unwarping. Second, they registered the T1 MR image for each individual to the standard MNI152 2×2×2 mm space. Third, they combined the two image warping together, conducted transformation from the GDC unwarped fMRI space to the MNI standard space, and registered the cleaned fMRI data from the previous step to the MNI standard space by applying the combined image warping. The above three steps were completed in the fMRI expert analysis tool (FEAT) from the software FSL.

For tfMRI, the same image cleaning and registration were applied as the above rfMRI, except that spatial smoothing using a Gaussian kernel of FWHM 5 mm was applied before the intensity normalisation, and no ICA+FIX artefact removal was run. Task-induced activation modelling was carried out using FEAT, and time-series statistical analysis was carried out using FILM with local autocorrelation correction (Woolrich et al., 2001). The timing of the two blocks including faces and shapes were recorded. The shape, face, and shape-face activation contrasts were major interests of the UK Biobank brain imaging team. These activation maps have been generated through the group-level fixed effect z-statistic with threshoded at $Z > 120$.

**HCP fMRI acquisition** We used the HCP young adult data which recruited $1,200$ healthy adults in the age range of 22 to 35 years old. Each participant underwent the MRI screening in two consecutive days to take their rsfMRI, tfMRI and the T1 and T2 weighted sMRI scans. The rsfMRI for each participant included 4 runs (15 minutes each) from two image sessions where each session had two phase-encoding directions (LR and RL). In the present study, to evaluate the reproducibility of the HCP rsfMRI functional connectivities, the RL runs at day 1 and day 2 were employed as the test and re-test datasets, respectively. The HCP rsfMRI data were acquired using the gradient-echo echo-planar imaging (GE-EPI) on a customized Siemens 3T "Connectome Skyra", which included a multi-band acceleration factor of 8, a 2-mm isotropic spatial resolution of voxels, a TR of 700 ms and TE of 33.1 ms, and a flip angle 52°. The receiver bandwidth was 2290 Hz/Px and the echo spacing was 0.58 ms. The HCP tfMRI data were acquired with the same EPI pulse sequence parameters as rsfMRI, except for the run duration information. There were seven tfMRI paradigms with 14 runs in opposing (L/R and R/L) phase-encoding directions, three of which (working memory, reward processing and motor processing) were included in one 30-minutes imaging session, and four of which (language, social cognition, relational processing and emotion processing) were taken in another 30 minutes imaging session. To match the UKB tfMRI data, we focused on the emotion processing task only. Each run of the task includes 176 frames which lasted for 136 seconds. Specifically, each run of the task included 3 face blocks, 3 shape blocks, and 8 seconds of fixation at the end of each run. Each of the block was composed of 6 trials and a 3000 ms task cue ("shape" or "face"), and each trial included a duration with the stimulus presented for 2000 ms and a 1000 ms intertrial interval (ITI). The two runs L/R and R/L emotion tfMRI data were employed in the present study as the test and re-test tfMRI datasets to evaluate the reproducibility of the connectivity. For sMRI, the T1-weighted images were acquired at 0.7 $mm^3$ isotropic resolution with TI/TR/TE = 1000/2400/2.14 ms and the T2-weighted images were acquired at 0.7 $mm^3$ isotropic resolution with TR/TE = 3200/565 ms. The bandwidth for the T1 and T2 MRI were 210 and 744 Hz/Px, respectively.

**HCP fMRI preprocessing** We downloaded the FIX-denoised rsfMRI data which underwent the standard HCP fMRIVolume pipeline and the additional rsfMRI processing pipeline, described in (WU-Minn, 2017) and (Smith et al., 2013), respectively. We also downloaded the minimally preprocessed tfMRI data which underwent the standard HCP fMRIVolume pipeline. The fMRIvolume pipeline removed spatial distortions from the gradient distortion correction, realigned volumes to for motion correction, corrected for the EPI distortion, and registered the EPI to the processed sMRIs through the brain-boundary-based registration. Then the fMRIvolume pipeline performed the intensity normalization (to the global mean) and bias field removal, and masked the data with the final brain mask. The sMRI data were processed by the gradient distortion correction, coregistration of the T1w and T2w sMRIs, bias field correction, and registration of the subject's native structural volume space to MNI152 $2 \times 2 \times 2$ mm space. The additional rsfMRI processing pipeline denoised the minimal processed data based on the fMRIVolume pipeline through the FIX independent components analysis (FIX-ICA) processment, and transformed to the MNI152 standard space through the warp field of the structural image registration. For tfMRI, spatial smoothing was applied to the minimally processed tfMRI images using an unconstrained 3D Gaussian kernel with full width at half maximum (FWHM) of 4 mm. For both the processed rsfMRI and tfMRI data, we performed an additional high-pass temporal filtering step with the sigma of 50 s to ensure the consistency with the UKB dataset.

**The Glasser360 and Schaefer200 atlases** The HCP-MMP 1.0 atlas or the Glasser 360 atlas proposed by Glasser et al. (2016) is a detailed group-based parcellation constructed using both high-resolution functional and anatomical MRI data from subjects of the HCP database. This atlas includes 360 cortical regions with 180-area per hemisphere. We used the HCP-MMP 1.0 volumetric parcellation `https://identifiers.org/neurovault.collection:1549`, which was converted from the original HCP-MMP 1.0 surface-based atlas to volumetric atlas by a series of conversion and transformation steps using FreeSurfer (Andreas, 2016). After the preprocessing steps in the above sections, this atlas was mapped onto both the UKB and HCP registered and cleaned rsfMRI and tfMRI datasets to extract the 360 regional time series. The amplitude traits and the full connectivity matrices were extracted from the time series standard deviation and the Gaussianised temporal correlation between every node time series pairs, respectively. Afterwards, we incorporated the network defined in Ji et al. (2019) which split the 360 regions into 12 networks. Similarly, we also analyzed the data using the Schaefer200 atlas (Schaefer et al., 2018), which partitioned the brain into 200 regions.

#### Legends for Tables S1 to S4 (All tables can be found in a zip file).

**Table S1: The 360 functional areas defined in the Glasser360 atlas.** More details can be found in Glasser et al. 2016 (`https://doi.org/10.1038/nature18933`).

**Table S2: The 200 functional areas defined in the Schaefer200 atlas.** More details can be found in Schaefer et al. 2018 (`https://doi.org/10.1093/cercor/bhx179`).

**Table S3: Summary of association results between functional connectivity measures and 647 traits in the UK Biobank.** The full set of results can be found here `http://165.227.92.206/traitList.html`.

**Table S4: Summary of intelligence prediction results using multiple data types.**
